# PedDx: An End-to-End Deep Learning Framework for Pediatric Posterior Fossa Tumor Diagnosis with Agentic AI–Enabled Independent External Evaluation

**DOI:** 10.64898/2026.10.27.26366616

**Authors:** Anahita Fathi Kazerooni, Fadel Batal, Bhavyasri Vunnava, Eswar Damaraju, Amin Tavallaii, Deep Gandhi, Anurag Gottipati, Pengjia Zhu, Dimosthenis Chrysochoou, Sanaz Varshochi, Sahand Adib Moradi, Ariana M. Familiar, Andrew Chiang, Yog Vaghani, Ghasem Azemi, Mikaela Doig, Minh-Son To, Amanda Luck, Meen Chul Kim, Neda Khalili, Nastaran Khalili, Paarth Jain, Celine Lee, Jordan R. Hansford, Antonio Di Ieva, Phillip B. Storm, Adam C. Resnick, Kartik Reddy, Arastoo Vossough, Ali Nabavizadeh

## Abstract

Reliable clinical translation of diagnostic artificial intelligence (AI) requires evaluation beyond classification accuracy. We developed PedDx, a segmentation-free framework for pediatric posterior fossa tumor diagnosis using T2-weighted MRI, pretrained 3DINO representations, calibrated predictions, and confidence-based abstention. Evaluation included 492 discovery, 114 replication, 129 external validation, and 40 independently tested cases, alongside 236 out-of-distribution (OOD) cases and controlled imaging perturbations. Classification accuracy was 79.8% in replication and 81.4% in external validation, increasing to 90.5% and 88.7%, respectively, among non-abstained cases. OOD analyses revealed performance degradation and confidently incorrect predictions, while perturbation analyses identified acquisition and alignment vulnerabilities. An external institution independently executed the complete agentic workflow, successfully generating predictions and structured quality-control reports for all 40 cases, with classification accuracy of 92.5%, increasing to 97.3% among non-abstained cases. PedDx demonstrates how uncertainty assessment, failure-mode characterization, external validation, and independently reproducible, traceable reporting can collectively inform the reliability and limitations of diagnostic AI systems.

## Introduction

Pediatric posterior fossa tumors comprise biologically and clinically distinct neoplasms, including medulloblastoma, ependymoma, atypical teratoid/rhabdoid tumor (ATRT), pediatric low-grade glioma (LGG), and diffuse midline glioma, including diffuse intrinsic pontine glioma (DMG/DIPG) ^1,2^. Accurate preoperative differential diagnosis can inform the goals and extent of surgery, the need for additional staging, and counseling about anticipated treatment and prognosis. These distinctions are particularly consequential in the posterior fossa, where the potential benefits of tumor resection must be weighed against the risk of injury to critical brainstem and cerebellar structures ^3–5^. MRI-based assessment can support these decisions before definitive diagnostic characterization, but overlapping imaging appearances make differentiation challenging even for experienced neuroradiologists ^6,7^. This challenge has motivated imaging-based artificial intelligence (AI) approaches to assist pediatric neuro-oncologic diagnosis ^8–10^.

Prior imaging-based AI studies have addressed radiomics-based classification ^8,9,11,12^ and deep learning for histological classification and molecular subtyping ^13,14^. These studies provide a foundation for automated classification, but diagnostic discrimination alone does not establish how a model will behave across the broader range of examinations encountered during external use. A multiclass classifier must be evaluated for class-specific errors, the reliability of its confidence estimates, and its ability to defer unconfident predictions ^15–17^. This is particularly important when aggregate performance can obscure limited recognition of less frequent tumor types.

Evaluation must also address inputs outside the intended diagnostic population, referred to as out-of-distribution (OOD) data. A model developed for treatment-naïve posterior fossa tumors may encounter postoperative changes, suboptimal imaging, tumors outside the posterior fossa, or rare histologies absent from its diagnostic categories. Such distribution shifts can compromise predictive performance without a corresponding reduction in confidence, potentially producing confident but incorrect predictions ^15,18–20^. Probability calibration and confidence-based abstention, complemented by systematic OOD and failure-mode analyses help a more comprehensive validation of a model. Evaluation across clinically relevant distribution shifts can identify vulnerabilities associated with unfamiliar anatomy, treatment effects, or unsupported histologies, while controlled imaging perturbations can characterize sensitivity to acquisition artifacts and preprocessing errors ^20,21^. These analyses help establish the conditions under which model predictions may become unreliable and whether uncertainty measures adequately identify such failures.

Beyond predictive performance, generalizability also depends on whether the complete AI workflow can be reliably executed across institutions. Conventional external validation assesses the performance of a fixed classifier on independently acquired data, whereas independent execution by an external institution additionally evaluates the reproducibility of preprocessing, quality control (QC), inference, and reporting. These considerations align with the AI-RAPNO recommendations for robust validation and clinical translation in pediatric neuro-oncology ^22,23^ and the broader FUTURE-AI consensus guidelines emphasizing generalizability, robustness, traceability, and continuous quality control in medical AI systems ^24^. Transparent documentation of processing steps, QC findings, model outputs, and potential failures can further facilitate troubleshooting and independent evaluation. These principles motivate evaluating diagnostic AI as an end-to-end system rather than solely through classification performance.

Here, we present PedDx, a confidence-based framework designed to classify five pediatric posterior fossa tumor categories from T2-weighted MRI, using a frozen three-dimensional vision transformer pretrained by self-supervision (3DINO ^25^) and a lightweight classification head. The classifier uses atlas-defined posterior fossa sampling without requiring tumor segmentation and incorporates probability calibration and confidence-based abstention as a safeguard against presenting uncertain outputs as diagnostic assignments, accepting that some correct predictions will also be withheld. To support reproducible deployment and transparent reporting, the classifier is integrated into an agentic workflow that orchestrates preprocessing, quality control, inference, and structured report generation. Language-model agents record processing and QC evidence to document potential issues and facilitate troubleshooting, while diagnostic probabilities and abstention decisions remain exclusively determined by the fixed imaging classifier.

We evaluate PedDx at both the classifier and workflow levels. We assess diagnostic performance, calibration, and abstention in discovery, replication, and external cohorts; characterize failure modes using clinical and biological distribution shifts and controlled imaging perturbations; and determine whether additional OOD scores improve identification of shifted inputs. Finally, an external team independently evaluates the complete workflow. These design and evaluation choices address complementary requirements for clinical translation: confidence-based deferral, independent end-to-end execution, and characterization of distribution shift and failure modes.

## Results

### Study overview

PedDx was evaluated for diagnostic performance within its intended (in-distribution) population, behavior under distribution shift, and generalizability of both the classifier and the complete workflow (Fig. 1). Model development used 492 treatment-naïve cases, along with internal replication in 114 cases on CBTN data and external classifier evaluation in 129 cases from Children’s Healthcare of Atlanta (CHOA). An external team at Macquarie University (Sydney, Australia) independently evaluated the deployed framework on 40 cases from the Women’s and Children’s Hospital (WCH; Adelaide, Australia). Additional analyses examined 236 clinical or biological OOD cases (clinical OOD, post-operative or post-treatment imaging, artifacts, and tumors outside the posterior fossa; biological OOD, rare posterior fossa tumor histologies not represented during model training) and controlled perturbations of 50 replication examinations. Cohort characteristics are summarized in Table 1 and Supplementary Note 1.

**Fig. 1.**
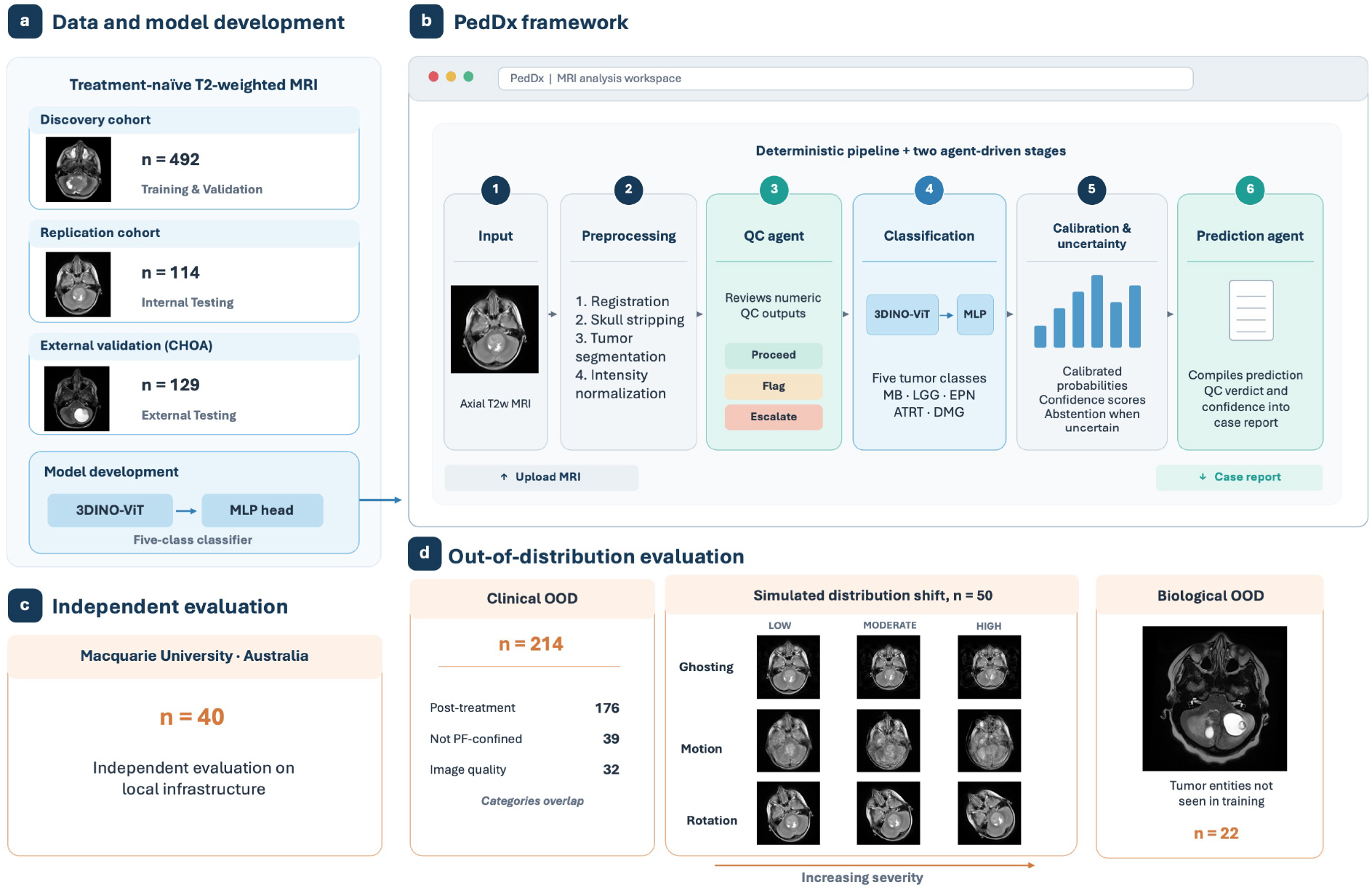
Study design and the PedDx framework. **a**, Data and model development. Treatment-naive axial T2-weighted MRI from the discovery cohort (n = 492) was used to train the five-class classifier, a frozen 3DINO vision transformer (3DINO-ViT) with a multilayer perceptron (MLP) head; the replication cohort (n = 114) was used for internal validation, and a five-class cohort from Children’s Healthcare of Atlanta (CHOA; n = 129; 102 medulloblastomas) for external validation. **b**, The PedDx framework as deployed in the web interface. An uploaded axial T2-weighted volume (1) is preprocessed by registration, skull stripping, tumor segmentation and intensity normalization (2) and assessed by the quality-control agent, which reasons over numeric quality-control outputs and assigns PROCEED, FLAG or ESCALATE (3). The frozen classifier assigns one of five classes (4), after which calibrated probabilities, a confidence score and an abstention rule are applied (5). The output (6) combines the prediction, class probabilities, confidence, quality-control verdict and a case report. **c**, Independent evaluation of the complete framework by the team at Macquarie University, Australia on data from Women’s and Children’s Hospital (WCH; Adelaide, Australia) (n = 40), run on local infrastructure. **d**, Out-of-distribution evaluation: clinical shift (post-treatment examinations, n = 176), simulated distribution shift (ghosting, rotation and motion at graded severity, applied to replication data, n = 50) and biological shift (tumor entities not seen in training, n = 22). ATRT, atypical teratoid/rhabdoid tumor; DMG, diffuse midline glioma; EPN, ependymoma; LGG, low-grade glioma; MB, medulloblastoma; OOD, out-of-distribution; QC, quality control.

**Table 1.** Cohort characteristics.

| Characteristic | Discovery (n = 492) | Replication (n = 114) | Children’s Healthcare of Atlanta (n = 129) | Women’s and Children’s Hospital (WCH; Adelaide, Australia) (n = 40) |
|---|---|---|---|---|
| Age, median (IQR), years | 6.9 (4.0–11.1) | 6.2 (3.6–10.6) | – | – |
| **Organization, n (%)** |  |  |  |  |
| The Children’s Hospital of Philadelphia | 370 (75.2) | 87 (76.3) | – | – |
| Lurie Children’s Hospital | 79 (16.1) | 21 (18.4) | – | – |
| Seattle Children’s Hospital | 13 (2.6) | 0 (0.0) | – | – |
| Orlando Health Arnold Palmer Hospital | 11 (2.2) | 1 (0.9) | – | – |
| Children’s National Medical Center | 9 (1.8) | 0 (0.0) | – | – |
| University of Pittsburgh | 3 (0.6) | 0 (0.0) | – | – |
| Weill Cornell Medical College | 3 (0.6) | 0 (0.0) | – | – |
| Dayton Children’s Hospital | 1 (0.2) | 3 (2.6) | – | – |
| Joseph M. Sanzari Children’s Hospital | 1 (0.2) | 1 (0.9) | – | – |
| UCSF Benioff Children’s Hospital | 1 (0.2) | 1 (0.9) | – | – |
| Wake Forest Baptist Medical Center | 1 (0.2) | 0 (0.0) | – | – |
| **Sex, n (%)** |  |  |  |  |
| Male | 275 (55.9) | 67 (58.8) | – | – |
| Female | 217 (44.1) | 47 (41.2) | – | – |
| **Histopathologic diagnosis, n (%)** |  |  |  |  |
| Medulloblastoma | 190 (38.6) | 48 (42.1) | 102 (79.1) | 20 (50.0) |
| Diffuse midline glioma / DIPG | 82 (16.7) | 16 (14.0) | 5 (3.9) | 9 (22.5) |
| Atypical teratoid/rhabdoid tumor | 25 (5.1) | 6 (5.3) | 5 (3.9) | 0 (0.0) |
| Low-grade glioma | 158 (32.1) | 36 (31.6) | 13 (10.1) | 11 (27.5) |
| Ependymoma | 37 (7.5) | 8 (7.0) | 4 (3.1) | 0 (0.0) |

Values are median (IQR) or n (%), unless otherwise indicated. –, not available or not applicable. DIPG, diffuse intrinsic pontine glioma; IQR, interquartile range.

### Diagnostic performance and uncertainty in the intended (in-distribution) population

The model combined a frozen 3DINO backbone with posterior fossa multi-crop sampling and a lightweight multiclass head (PedDx v2). Macro-AUC-ROC was 0.901 ± 0.023 in discovery and 0.949 in replication; corresponding five-class accuracies were 74.2% ± 2.5% and 79.8% (Table 2; Fig. 2). Performance was strongest for DMG/DIPG, LGG, and medulloblastoma, with replication AUC-ROCs of 0.998, 0.976, and 0.968, respectively. However, five-class balanced accuracy was substantially lower (0.587 ± 0.065 and 0.647), reflecting limited diagnostic performance for ATRT and ependymoma. Performance metrics on ependymoma indicate the distinction between discrimination and classification: its replication AUC-ROC was 0.934, but sensitivity was 0.125. Complete class-wise metrics are provided in Table 2.

**Fig. 2.**
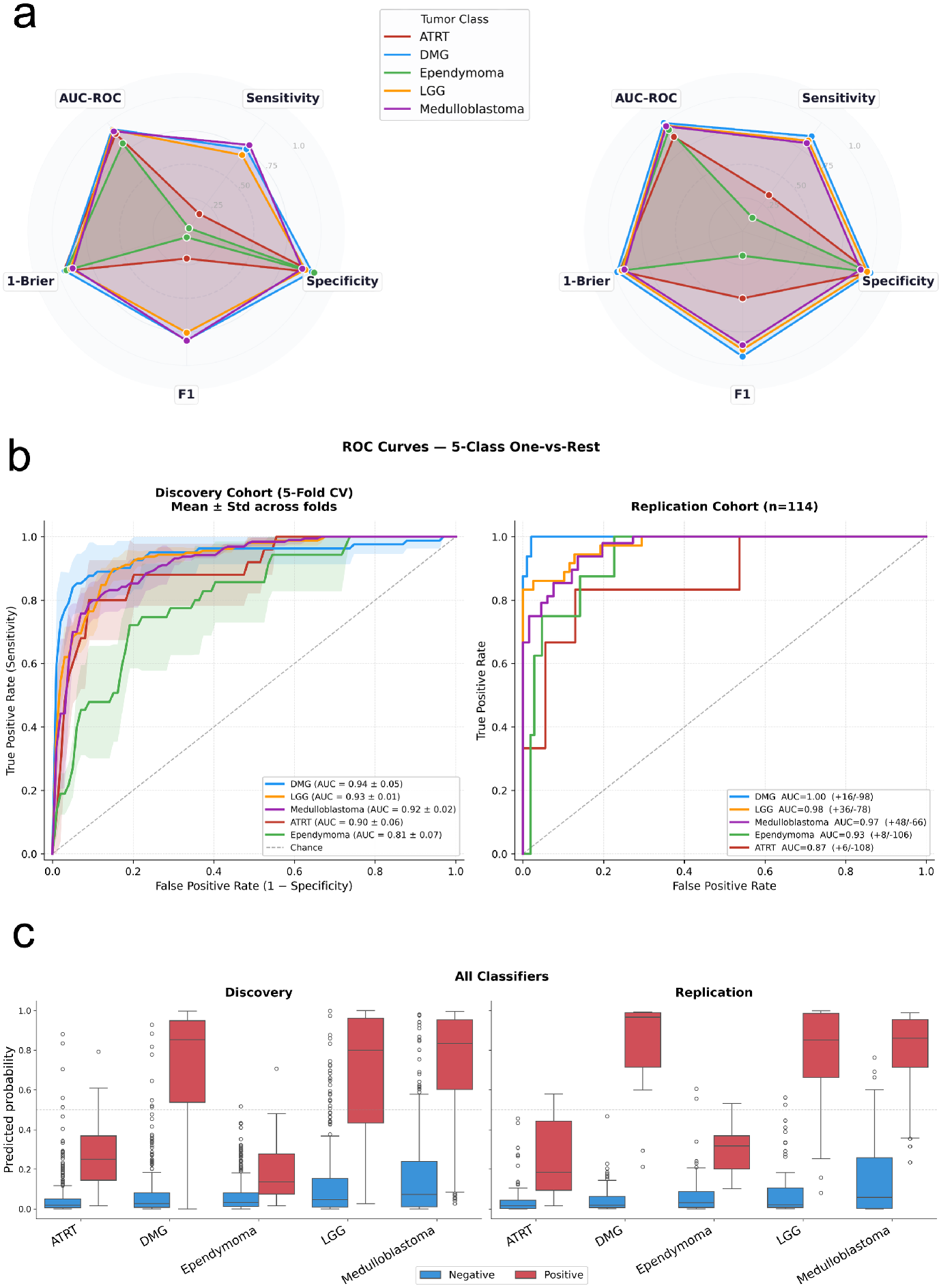
Diagnostic performance of the PedDx classifier across discovery and replication cohorts. **a**, Radar plots comparing AUC-ROC, sensitivity, specificity, F1-score, and Brier score (plotted as 1−Brier) for each tumor class in the discovery (left) and replication (right) cohorts. b, Receiver operating characteristic (ROC) curves for each of the five posterior fossa tumor classes in the discovery and replication cohorts, with area under the curve (AUC) reported per class; in the replication panel, (+n/−n) gives the number of positive and negative cases for that class. **c**, Calibrated predicted probability distributions after temperature scaling per class, shown separately for the discovery (left) and replication (right) cohorts; red indicates true-positive cases and blue indicates true-negative cases. All panels use calibrated probabilities (T = 1.30).

**Table 2.** Per-class performance on the discovery cohort (n = 492), mean ± s.d. across five folds, and on the replication cohort (n = 114). Balanced accuracy, F1, sensitivity and specificity are computed at a 0.5 threshold on each class’s calibrated probability; balanced accuracy is the mean of sensitivity and specificity for that class. The macro average is the unweighted mean of the five per-class values.

| Class | Cohort | Balanced accuracy | F1 | AUC-ROC | Sensitivity | Specificity |
|---|---|---|---|---|---|---|
| **ATRT** | Discovery | 0.575 ± 0.110 | 0.203 ± 0.282 | 0.898 ± 0.062 | 0.160 ± 0.219 | 0.989 ± 0.011 |
|  | Replication | 0.667 | 0.500 | 0.870 | 0.333 | 1.000 |
| **DMG/DIPG** | Discovery | 0.869 ± 0.074 | 0.812 ± 0.112 | 0.942 ± 0.059 | 0.757 ± 0.140 | 0.981 ± 0.014 |
|  | Replication | 0.938 | 0.933 | 0.998 | 0.875 | 1.000 |
| **Ependymoma** | Discovery | 0.513 ± 0.029 | 0.044 ± 0.099 | 0.810 ± 0.081 | 0.029 ± 0.064 | 0.998 ± 0.005 |
|  | Replication | 0.553 | 0.182 | 0.934 | 0.125 | 0.981 |
| **LGG** | Discovery | 0.815 ± 0.036 | 0.756 ± 0.046 | 0.933 ± 0.007 | 0.702 ± 0.081 | 0.928 ± 0.019 |
|  | Replication | 0.904 | 0.882 | 0.976 | 0.833 | 0.974 |
| **Medulloblastoma** | Discovery | 0.849 ± 0.031 | 0.815 ± 0.036 | 0.921 ± 0.020 | 0.795 ± 0.071 | 0.904 ± 0.027 |
|  | Replication | 0.868 | 0.848 | 0.968 | 0.812 | 0.924 |
| **Macro average** | Discovery | 0.724 ± 0.026 | 0.526 ± 0.054 | 0.901 ± 0.023 | 0.489 ± 0.050 | 0.960 ± 0.008 |
|  | Replication | 0.786 | 0.669 | 0.949 | 0.596 | 0.976 |

ATRT, atypical teratoid/rhabdoid tumor; AUC-ROC, area under the receiver operating characteristic curve; DMG/DIPG, diffuse midline glioma/diffuse intrinsic pontine glioma; LGG, low-grade glioma.

Development analyses supported the selected configuration. Multi-crop sampling increased macro-AUC-ROC by 0.023 relative to a single-centered crop, while alternative multi-view counts and offset magnitudes did not differ significantly (Supplementary Note 2). Against the fine-tuned ResNet-34 benchmark (PedDx v1), the fold-averaged macro-AUC-ROC difference was 0.013 (95% CI, −0.010 to 0.036; q = 0.33), and no replication difference remained significant after multiplicity correction. PedDx v2 used approximately 1,000-fold fewer trainable parameters and showed less variation in score scale across fold models. Detailed benchmark metrics and the distinction between fold-averaged and pooled comparisons are reported in Supplementary Note 3 and Supplementary Tables 3–10.

Temperature scaling (T = 1.30) reduced expected calibration error from 0.066 to 0.039 in discovery and from 0.092 to 0.086 in replication (Supplementary Note 4). At the fixed confidence threshold of 0.50, PedDx v2 abstained on 19 of 114 replication cases (16.7%), retaining predictions for 95 (83.3%). Abstention intercepted 14 of 23 errors, increasing accuracy among retained cases to 90.5% (86/95), compared with 79.8% without abstention (Fig. 3a). All 62 high-confidence predictions (≥0.80) were correct, whereas 24 of 33 moderate-confidence predictions were correct (72.7%). The nine remaining errors show that abstention reduced, but did not eliminate, misclassification. Representative correct and incorrect predictions with similar moderate confidence are shown in Fig. 3b.

**Fig. 3.**
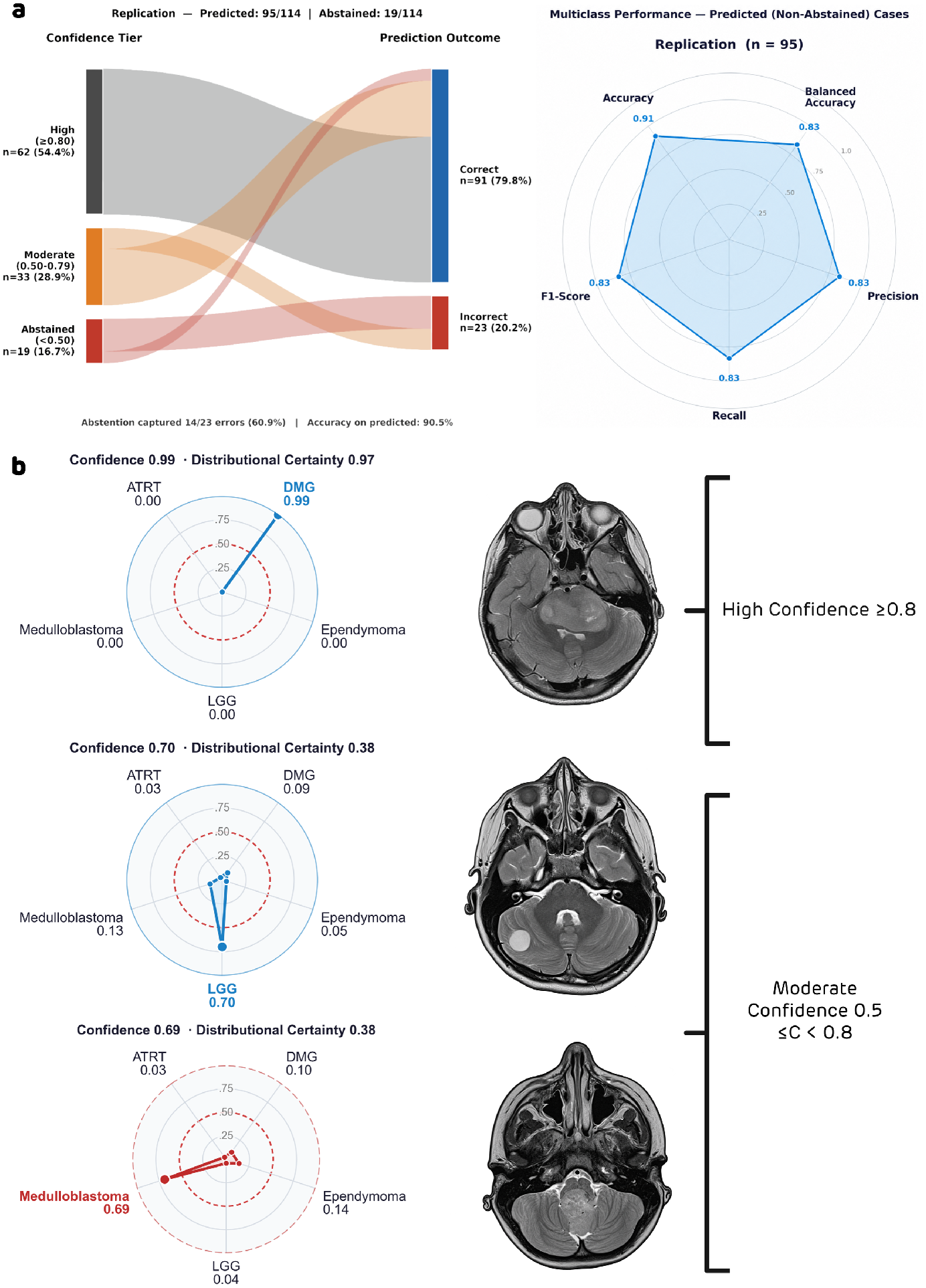
Confidence-tier stratification, multiclass performance, and representative case probability distributions in the replication cohort. **a**, Sankey diagram (left) illustrating the flow of replication cohort cases (n = 114) from calibrated confidence tiers, high (≥0.80, n = 62), moderate (0.50–0.79, n = 33), and abstained (< 0.50, n = 19), to final classification outcomes. The abstention rule captured 14 of 23 total cohort errors (60.9%), increasing diagnostic accuracy among non-abstained predicted cases (n = 95) from 79.8% to 90.5%. Radar plot (right) summarizing multiclass evaluation metrics calculated on non-abstained predicted cases. **b**, Five-class calibrated probability distribution radar plots and corresponding axial T2w scans for representative cases across confidence tiers. Top panel: High-confidence correct prediction of diffuse midline glioma (confidence = 0.99, distributional certainty = 0.97). Middle panel: Moderate-confidence correct prediction of low-grade glioma (confidence = 0.70, distributional certainty = 0.38). Bottom panel: Moderate-confidence misclassified prediction of medulloblastoma (confidence = 0.69; distributional certainty = 0.38). All probabilities reflect post-temperature scaling (T = 1.30) outputs. DMG, diffuse midline glioma; LGG, low-grade glioma; ATRT, atypical teratoid rhabdoid tumor.

### External evaluation and class-specific limitations

The fixed classifier was evaluated on the five-class CHOA cohort without updating model weights, calibration, or the abstention threshold. Accuracy without abstention was 81.4% (105/129; 95% CI, 74.4–87.6%). Abstention excluded 23 cases (17.8%) and intercepted 12 of 24 errors; accuracy among the 106 retained cases was 88.7% (94/106; 95% CI, 82.1–94.3%) (Fig. 4a). High-confidence predictions were correct in 61 of 65 cases (93.8%), indicating that the confidence–accuracy relationship extended to this external cohort, although high confidence did not guarantee correctness.

**Fig. 4.**
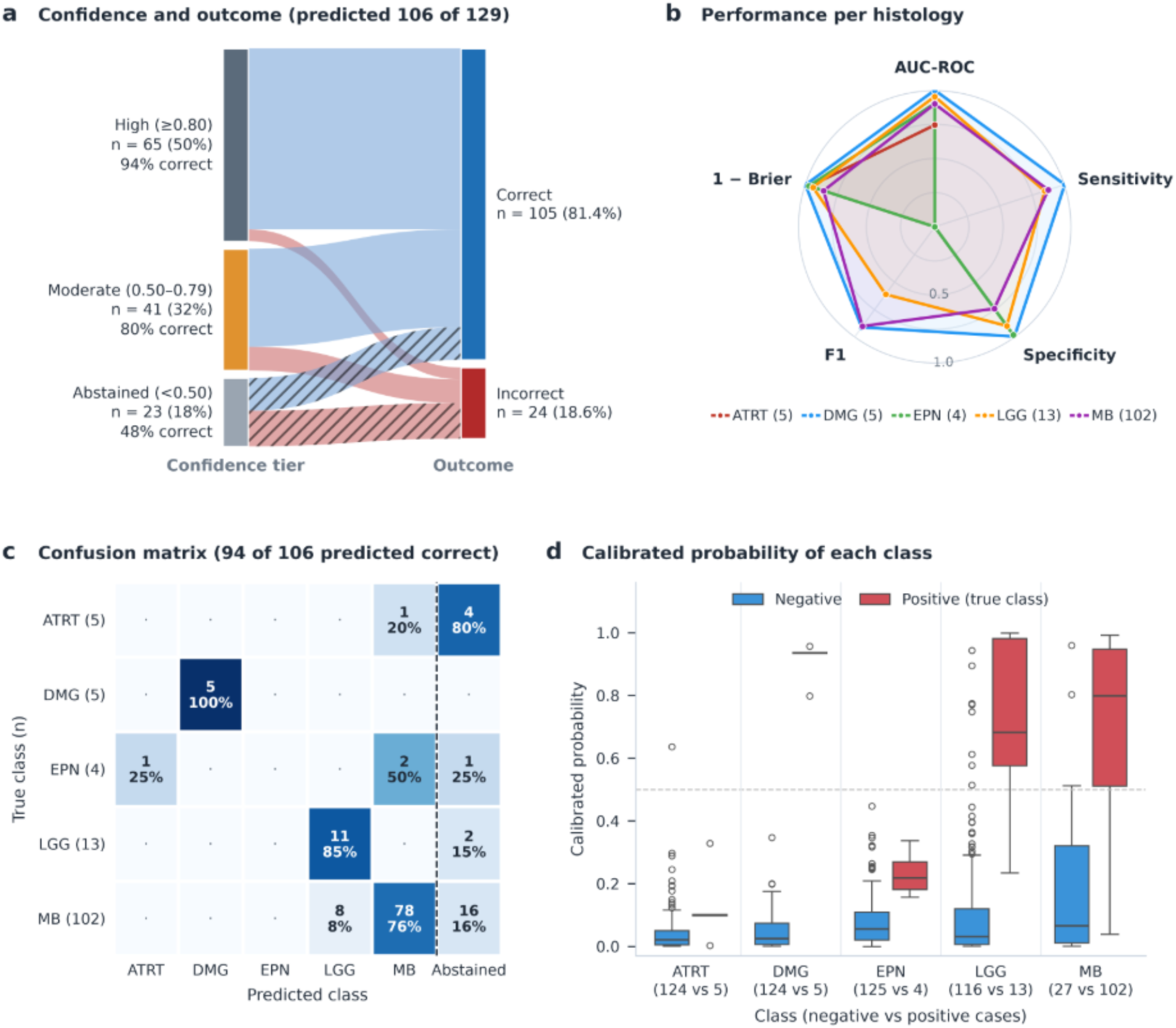
External evaluation on the five-class CHOA cohort (n = 129). a, Flow of cases from calibrated confidence tier, high (≥0.80, n = 65), moderate (0.50–0.79, n = 41) and abstained (<0.50, n = 23), to outcome without abstention; abstained cases are shown as hatched flows. b, Performance per histology (AUC-ROC, sensitivity, specificity, F1 and 1 − Brier score) at the most probable class, on all 129 cases. c, Confusion matrix with abstention at a calibrated probability of 0.50; abstained cases are in the final column; cells give the count and the percentage of the row. d, Calibrated probability of each class for positive (true class) and negative cases; the dashed line marks 0.50. CHOA, Children’s Healthcare of Atlanta; ATRT, atypical teratoid/rhabdoid tumor; DMG/DIPG, diffuse midline glioma/diffuse intrinsic pontine glioma; EPN, ependymoma; LGG, low-grade glioma; MB, medulloblastoma.

External performance remained class-dependent. Without abstention, PedDx correctly classified all five DMG/DIPG cases, 11 of 13 LGGs, and 89 of 102 medulloblastomas, but none of the five ATRT or four ependymoma cases (Table 3; Fig. 4b,c). Five of these nine rare-class cases (ependymoma and ATRT) led to abstention. Among retained cases, the 12 errors comprised eight medulloblastomas labeled LGG and four ATRT or ependymoma cases assigned another histology. Thus, the external cohort reproduced the internal pattern of stronger performance for the three common classes and poor diagnosis of the two rarer classes. Because medulloblastoma accounted for 102 of 129 cases, overall accuracy should be interpreted alongside class-wise performance and the small samples for other diagnoses.

**Table 3.** Per-class performance on the external CHOA cohort (n = 129). One-versus-rest metrics from the most probable class without abstention, with AUC-ROC from the calibrated probability of each class. Sensitivity after abstention is computed over cases on which the model did not abstain (maximum calibrated probability ≥ 0.50). Median probability is the median calibrated probability of the class among cases of that class (positive) and all other cases (negative).

| **Histology** | **n** | **AUC-ROC** | **Sensitivity** | **Specificity** | **PPV** | **Balanced accuracy** | **Abstained, n** | **Sensitivity after abstention** | **Median probability, positive / negative** |
|---|---|---|---|---|---|---|---|---|---|
| ATRT | 5 | 0.745 | 0.00 | 0.99 | 0.00 | 0.50 | 4 | 0.00 | 0.100 / 0.021 |
| DMG | 5 | 1.000 | 1.00 | 0.99 | 0.83 | 1.00 | 0 | 1.00 | 0.936 / 0.024 |
| Ependymoma | 4 | 0.904 | 0.00 | 0.98 | 0.00 | 0.49 | 1 | 0.00 | 0.218 / 0.055 |
| LGG | 13 | 0.954 | 0.85 | 0.90 | 0.48 | 0.87 | 2 | 1.00 | 0.682 / 0.031 |
| Medulloblastoma | 102 | 0.900 | 0.87 | 0.74 | 0.93 | 0.81 | 16 | 0.91 | 0.799 / 0.065 |

CHOA, Children’s Healthcare of Atlanta; ATRT, atypical teratoid/rhabdoid tumor; AUC-ROC, area under the receiver operating characteristic curve; DMG, diffuse midline glioma; PPV, positive predictive value; LGG, low-grade glioma.

To distinguish institutional shift from differences in histologic composition, we also compared CHOA and discovery medulloblastomas using the OOD scores. Free energy provided little separation after class matching (AUC-ROC, 0.521; 95% CI, 0.452–0.591), indicating limited detectable distribution shift by this score within the matched class (Supplementary Tables 18–19). Class-probability distributions are shown in Fig. 4d.

### Failure modes under clinical and biological distribution shift: out-of-distribution (OOD) analysis

Clinical shift markedly reduced accuracy, with a smaller reduction in confidence. Accuracy without abstention fell from 79.8% in replication to 38.1% in clinical OODs with post-treatment imaging, 28.1% in clinical OODs with inadequate image quality, and 17.9% for tumors outside the posterior fossa. Median confidence decreased from 0.802– 0.830 in the internal in-distribution cohorts to 0.535–0.608 across clinical-shift strata, while abstention increased to 35.9–37.5% (Supplementary Table 12). Unlike its effect in the replication set, abstention provided little recovery of accuracy in OOD cohort: retained-case accuracy was 39.1% for post-treatment cases, 30.0% for cases with inadequate image quality, and 24.0% for tumors outside the posterior fossa. Confidence-based selection therefore did not preserve diagnostic performance outside the intended population.

We assessed whether additional scores could identify these shifted inputs. Among 20 post-hoc scores, free energy separated the clinical-shift strata from discovery (in-distribution) cohort with AUC-ROCs of 0.728–0.774. It modestly improved on maximum softmax probability for post-treatment and anatomically atypical cases (AUC-ROC differences, 0.030 and 0.043; adjusted P = 0.004 and 0.0065, respectively), but feature-distance scores did not outperform the softmax baseline across clinical strata. None of the scores significantly separated the independent replication control from discovery after multiplicity correction (Supplementary Tables 14–17). Class matching reduced detection of anatomically atypical tumors from 0.746 to 0.663, showing that some pooled separation reflected differences in histologic composition; post-treatment detection was largely unchanged (Supplementary Tables 18–19).

Biological OOD cases exposed a different limitation. Fifteen of 22 tumors outside the five-class labels (68.2%) exceeded the confidence threshold and received a modeled diagnosis. Only two scores—crop mutual information and crop argmax instability— significantly separated these cases from discovery after correction, each with an AUC-ROC of 0.716. At a threshold set to the discovery reference’s 95th percentile, however, they detected only 9.1% and 0% of biological OOD cases, respectively (Supplementary Tables 20–21). Consequently, moderate ranking-based separation did not translate into reliable identification at this operating point, and the routine abstention rule failed to reject most unsupported histologies.

Exploratory feature-space analyses provided context for the limited utility of distance-based detection. Frozen embeddings showed weak histologic clustering despite supporting classification, and biological OOD cases were not unusually distant from the discovery center (median 40th percentile of discovery distances). These observations suggest that proximity in the frozen feature space was insufficient to establish whether a tumor belonged to the supported diagnostic classes (Supplementary Note 7; Supplementary Fig. 8; Supplementary Tables 22–25).

### Controlled perturbations identify acquisition and alignment vulnerabilities

Paired perturbations of 50 replication cases tested sensitivity to specific degradations while holding patient and histology constant. Accuracy without abstention declined from 80.0% on unperturbed images to 68.0% at the highest ghosting severity and 64.0% at the highest motion severity (Fig. 5a,b). Severity trends were significant for both artifacts (P = 0.013 and P = 0.006, respectively). Motion increased abstention from 18.0% to 36.0%, whereas ghosting did not produce a consistent increase, indicating different uncertainty responses to the two artifact types (Fig. 5e).

**Fig. 5.**
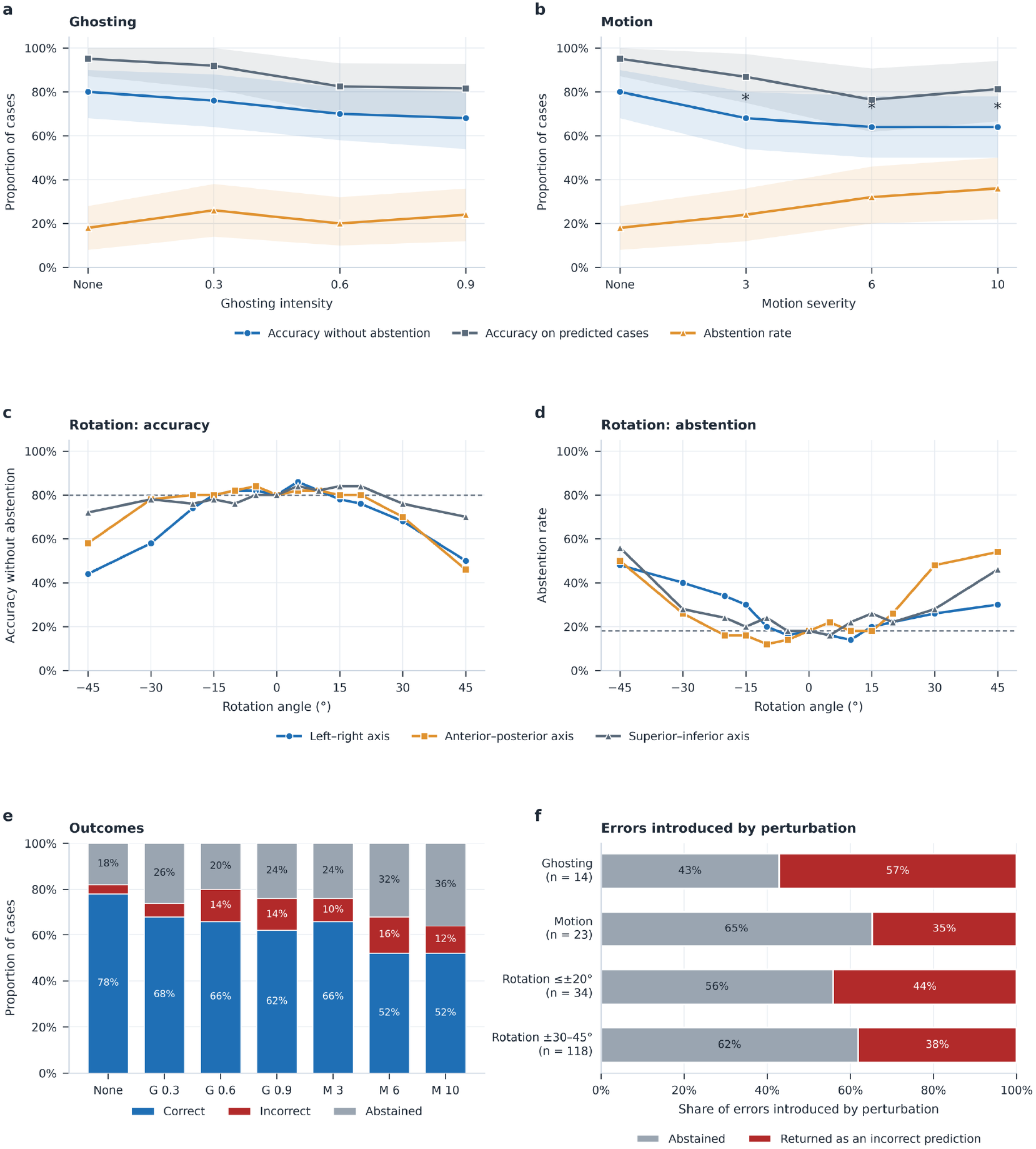
Sensitivity of the PedDx classifier to simulated imaging perturbations in the replication cohort (n = 50). a, Accuracy without abstention, accuracy on predicted cases and abstention rate by ghosting intensity. b, The same by motion severity; asterisks mark levels at which correctness differed from the unperturbed reference (exact McNemar test, Holm-adjusted P < 0.05). Shaded bands are subject-level bootstrap 95% confidence intervals (2,000 resamples). c, Accuracy without abstention under rigid rotation about the left–right, anterior–posterior and superior–inferior axes, applied to the preprocessed volume; the dashed line marks the unperturbed reference. d, Abstention rate under rotation. e, Correct, incorrect and abstained cases without perturbation and at each ghosting (G) and motion (M) level. f, Errors introduced by perturbation, defined as incorrect perturbed results in subjects whose unperturbed prediction was correct, divided into those that led to abstention and those returned as an incorrect prediction; n is the number of such errors pooled across levels, and rotation is pooled across axes and directions. Ghosting and motion images had negative intensities set to zero after skull stripping, before normalization. A case was abstained when its maximum calibrated probability was below 0.50.

Rotations applied after preprocessing evaluated sensitivity to residual misalignment of the atlas-defined sampling region. Aggregate accuracy remained between 74.0% and 86.0% for rotations up to ±20°, with no significant difference from the reference after correction, although individual predictions could change. Larger rotations produced axis-dependent degradation: rotations about the left-right axis reduced accuracy to 44.0– 50.0% at ±45°, while rotations about the superior-inferior axis primarily increased abstention (Fig. 5c,d).

Across perturbation conditions, abstention intercepted 113 of 189 newly incorrect results (59.8%) arising from cases correctly classified at baseline; the remaining 76 were returned as incorrect predictions (Fig. 5f). These counts represent subject–condition results, not unique patients. Confidence became less informative about errors as degradation increased: error-detection AUC-ROC fell from 0.97 without perturbation to 0.75 under 30–45° rotations. The controlled analyses therefore identified both sensitivity to degraded inputs and incomplete protection from confidence-based abstention. Full severity-specific results are provided in Supplementary Note 8 and Supplementary Tables 26–27.

### Independent evaluation of the complete workflow

Macquarie University independently executed the deployed framework on 40 examinations from patients diagnosed at Women’s and Children’s Hospital (WCH; Adelaide, Australia), without involvement from the development team. All cases completed processing and produced a report and QC record (Supplementary Note 9). The classifier returned 37 diagnoses and three abstentions; accuracy was 92.5% without abstention (37/40; 95% CI, 80.1-97.4%) to 97.3% among retained cases (36/37; 95% CI, 86.2-99.5%; Fig. 6a). The remaining error was a DMG/DIPG assigned medulloblastoma with confidence of 0.84. This cohort contained medulloblastoma, LGG, and DMG/DIPG only, so it did not evaluate the framework’s diagnostic performance for ATRT or ependymoma. Per-class results are provided in Fig. 6b,c and Supplementary Table 28.

**Fig. 6.**
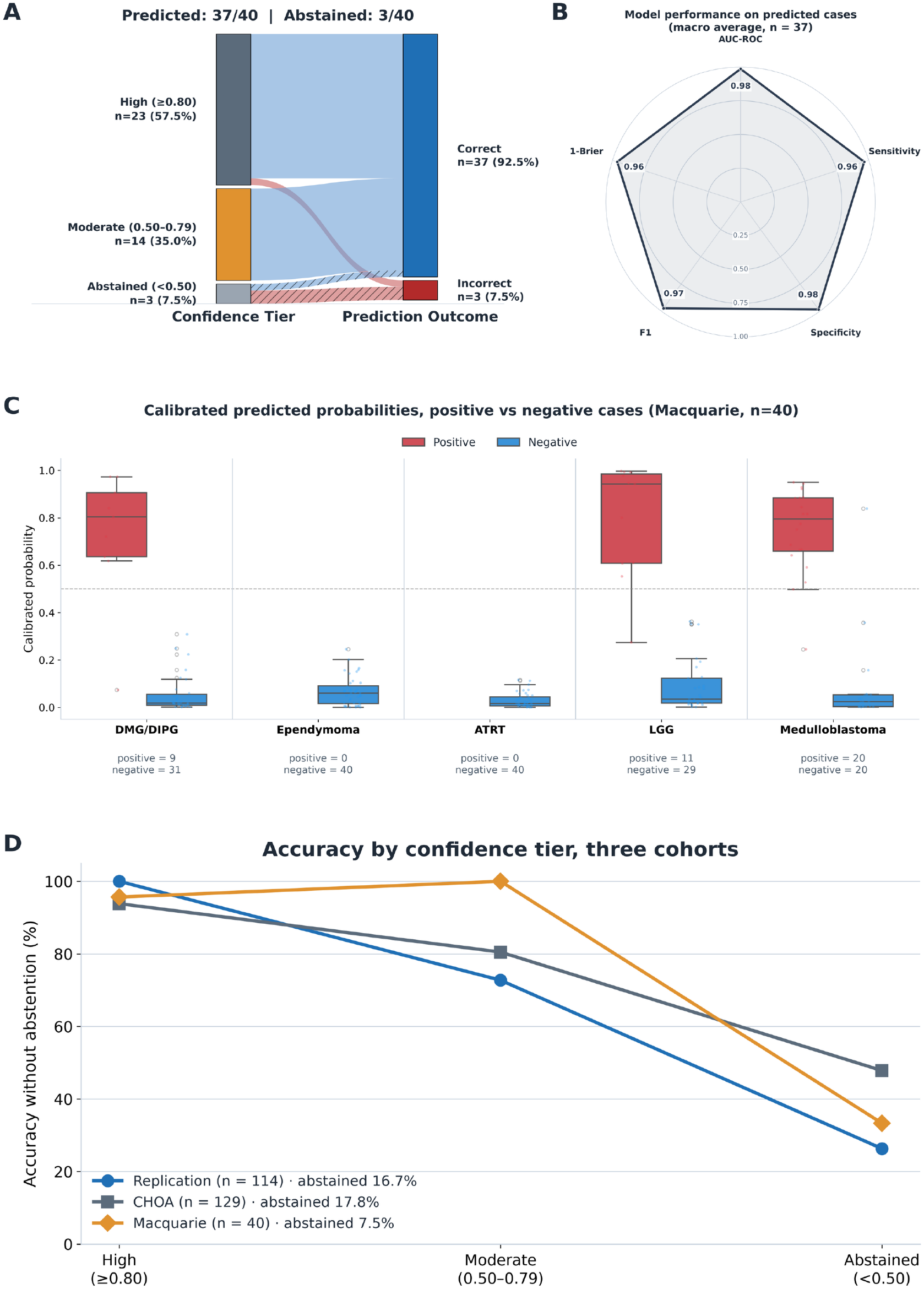
Independent evaluation of the framework by the team at Macquarie University on data from WCH (n = 40). a, Flow of cases from calibrated confidence tier, high (≥0.80, n = 23), moderate (0.50–0.79, n = 14) and abstained (<0.50, n = 3), to outcome; abstained cases are shown as hatched flows and scored without abstention. b, Macro-averaged one-vs-rest performance on the 37 predicted cases (AUC-ROC, sensitivity, specificity, F1 and 1 − Brier score), averaged over DMG/DIPG, LGG and medulloblastoma, the three histologies in this cohort. c, Per-class calibrated probabilities for positive (true class) and negative cases across all 40 subjects; the dashed line marks 0.50; the cohort contained no ATRT or ependymoma. d, Accuracy without abstention by confidence tier for the replication (n = 114), CHOA (n = 129) and Macquarie (n = 40) cohorts at the same fixed thresholds; legend values give the proportion of abstained cases. CHOA, Children’s Healthcare of Atlanta; DMG/DIPG, diffuse midline glioma/diffuse intrinsic pontine glioma; LGG, low-grade glioma.

Across replication, CHOA, and WCH, the same confidence thresholds yielded high-confidence accuracies of 93.8–100%, with abstention rates of 16.7%, 17.8%, and 7.5%, respectively (Fig. 6d). These findings demonstrate application of the fixed classifier and decision rule across the evaluated settings, while the independent Macquarie execution additionally establishes feasibility of running the complete workflow on external infrastructure.

The QC agent assigned PROCEED to 23 cases, FLAG to 14, and ESCALATE to three (Supplementary Fig. 12). These states characterized processing evidence but did not identify additional diagnostic errors beyond abstention in this cohort: non-PROCEED status identified one of the three errors, which abstention also intercepted, whereas abstention identified two additional cases not identified by the QC agent. Withholding all non-PROCEED cases would additionally have withheld 16 correct predictions. Thus, successful independent execution and structured QC reporting did not establish an improvement in diagnostic accuracy from the agentic layer.

## Discussion

PedDx combines clinically relevant pediatric brain tumor classification with a framework for evaluating how diagnostic AI could be independently tested and used responsibly. Its strongest diagnostic results were observed for medulloblastoma, LGG, and DMG/DIPG, including in external cohorts and independent testing on institutional infrastructure. Beyond differential diagnosis, the study examined uncertainty, abstention, clinical and biological OOD inputs, and controlled imaging perturbations within an executable workflow. These complementary assessments address questions central to prospective evaluation: whether performance transfers across institutions, when predictions should be withheld, which unfamiliar inputs remain confidently misclassified, and whether another institution can reproduce the complete processing and reporting pathway.

The medulloblastoma findings are particularly relevant. Preoperative diagnosis of this malignant embryonal tumor, which is the most common malignant brain tumors of childhood, can inform diagnostic assessment and planning ^1,2^. Medulloblastoma management requires coordinated staging and multimodal treatment, making its distinction from other pediatric tumors an important clinical question, although definitive diagnosis and treatment planning require pathological and molecular characterization ^4,9^. Suspicion of a medulloblastoma tumor can prompt whole-spine MRI before surgery, when feasible, avoiding postoperative changes that can mimic or obscure leptomeningeal dissemination ^26^. In the CHOA external cohort, PedDx correctly classified 89 of 102 medulloblastomas before abstention and 78 of 86 retained cases after applying the prespecified rule. It also correctly classified 11 of 13 LGGs and all five DMGs before abstention; all retained LGG and DMG cases were correct. Independent testing by the Macquarie team on WCH data yielded 92.5% accuracy across 40 medulloblastoma, LGG, and DMG cases, with 36 of 37 retained predictions correct. These findings support high diagnostic performance and generalizability of the PedDx model for these three diagnoses, particularly medulloblastoma, while the smaller LGG and DMG samples in the external set warrant cautious interpretation.

Previous studies have established the feasibility of pediatric brain tumor classification across institutions ^9,10,13,27^. Quon et al. demonstrated multicenter detection and classification of posterior fossa tumors using T2-weighted MRI ^10^. Lee et al.’s FL-PedBrain extended this field through an international federated platform for tumor segmentation and classification, including independent external holdouts ^27^. More recently, Jin et al. evaluated a T2-weighted MRI pipeline for segmentation, classification, and molecular subtyping using internal and external cohorts ^13^. PedDx builds on these advances by bringing foundation model adaptation, confidence-guided abstention, complementary OOD analyses, controlled failure testing, and independently executed agentic orchestration into a single study. One of the main distinctive contributions of PedDx is the breadth and integration of evidence used to characterize both diagnostic capability and conditions relevant to real-world translation.

The foundation model approach provides a practical basis for this framework. PedDx adapts frozen representations from 3DINO ^25^, a self-supervised model developed for transferable three-dimensional medical image analysis. A lightweight classifier uses features from anatomically defined crops of T2-weighted MRI, without requiring a tumor mask for diagnostic inference, providing an application and evaluation of pretrained volumetric representations in a clinically focused diagnostic setting. Parameter efficiency offers a feasible adaptation strategy where labeled pediatric imaging data are limited.

Uncertainty and abstention were incorporated as safeguards for clinical translation. Communicating uncertainty and allowing a model to defer are recognized approaches to supporting responsible medical decision making ^15,17^. In PedDx, calibration and the abstention threshold were fixed using discovery data and transferred without external adjustment. The rule intercepted 14 of 23 replication errors and 12 of 24 CHOA errors, while also withholding five and eleven correctly classified cases, respectively. This trade-off is central to its intended use: abstention reduces the occasions on which an uncertain prediction is presented as an actionable diagnosis, at the cost of additional cases requiring review. Accuracy among retained predictions must therefore be interpreted alongside coverage and the burden of deferral ^17^. The safeguard helps temper overconfident use of model outputs but cannot eliminate confidently incorrect predictions.

Independent execution provides an additional dimension of evidence beyond conventional external validation. The Macquarie evaluation demonstrated that the complete PedDx workflow could be executed on external institutional infrastructure without developer involvement, with all 40 examinations completing processing and generating structured reports. This operational reproducibility addresses an important aspect of AI generalizability that cannot be established by evaluating classifier predictions on externally acquired data alone. Consistent with FUTURE-AI recommendations emphasizing traceability, documentation, and continuous quality control ^24^, the agentic framework integrates preprocessing, QC, inference, and reporting while maintaining records of intermediate processing findings and potential failures. Language-model agents coordinate predefined tools and summarize structured QC evidence, enabling case-level reports that support troubleshooting, interpretation of model behavior, and retrospective review of processing decisions. Importantly, the agents cannot modify diagnostic probabilities or abstention decisions, preserving separation between diagnostic inference and workflow orchestration. This architecture illustrates how agentic systems can support transparent and independently reproducible evaluation of medical AI, although the added value of language-model orchestration over deterministic workflows remains to be established.

The OOD and perturbation analyses revealed vulnerabilities that would not be apparent from conventional external validation alone. Clinical OOD testing examined changes in treatment status, image quality, and anatomical tumor location; biological OOD testing assessed tumors outside the supported diagnostic categories; and controlled perturbations characterized sensitivity to acquisition artifacts and residual registration errors. Although confidence generally decreased under clinical distribution shift, substantial errors persisted among retained predictions, and 15 of 22 biologically unfamiliar tumors received a modeled diagnosis without abstention. These findings demonstrate that calibration and confidence-based deferral, while useful within the intended population, do not reliably identify all unsupported inputs or prevent confidently incorrect predictions under distribution shift, consistent with prior evidence regarding uncertainty degradation ^20^. This reinforces the importance of the robustness and risk assessment principles emphasized by FUTURE-AI ^24^ and the need for comprehensive validation and characterization of model limitations advocated by AI-RAPNO ^23^. Importantly, the OOD scores evaluated here provided limited additional protection, indicating that confidence, QC measures, and distribution-shift detection should be considered complementary rather than interchangeable safeguards. Systematically documenting these failure modes can inform future eligibility criteria, quality-control procedures, escalation pathways, and post-deployment monitoring strategies.

Several limitations define the scope of the findings. Diagnostic performance was uneven across histologies: none of the five ATRTs or four ependymomas in the CHOA cohort were correctly classified before abstention. PedDx therefore cannot currently be considered a reliable five-histology differential diagnostic system, and its strongest evidence concerns medulloblastoma, LGG, and DMG. Retrospective selection, class imbalance, and small rare-tumor and biological OOD samples further constrain generalizability. Larger, more diverse training cohorts may improve discrimination of underrepresented histologies, including ATRT and ependymoma, and enable expansion to currently unsupported diagnoses such as ETMR. Reliance on T2-weighted MRI excludes potentially useful information from complementary sequences, particularly diffusion-weighted imaging (DWI), for distinguishing medulloblastoma, ependymoma, and pilocytic astrocytoma.

Although independent execution demonstrated technical feasibility and reproducible reporting, the relatively small external cohort and its restricted histological composition limit conclusions about broader clinical performance. Furthermore, agent-assigned QC states did not identify additional diagnostic errors beyond confidence-based abstention in this cohort, and the incremental benefits of agentic orchestration for error detection, efficiency, or usability remain unestablished. Finally, the framework has not been evaluated prospectively in clinical practice, and its effects on clinician decision-making, workflow burden, and patient outcomes remain unknown.

In conclusion, PedDx illustrates the importance of evaluating diagnostic AI systems beyond conventional classification accuracy through complementary assessments of uncertainty, distribution shift, failure modes, and independent end-to-end execution. These evaluations provide evidence of both diagnostic strengths and limitations, while structured QC and reporting offer a foundation for traceability and troubleshooting. Future studies should assess the framework in prospective clinical settings, including its effects on clinician decision-making, reliance on AI predictions, review burden, and consequences of incorrect diagnoses or unnecessary abstention. Such evaluations align with DECIDE-AI recommendations emphasizing transparent reporting of early-stage clinical AI evaluation, including safety, human factors, and clinical utility ^28^. By combining diagnostic modeling with systematic reliability assessment and independently reproducible execution, PedDx provides a framework for investigating the requirements and limitations of AI-assisted pediatric brain tumor diagnosis.

## Methods

### Study design and cohorts

We retrospectively reviewed imaging and clinical data from the Children’s Brain Tumor Network (CBTN), which is a publicly-accessible biorepository, and the Children’s Hospital of Philadelphia (CHOP) under an approved IRB protocol 18-015588. External data from Children’s Healthcare of Atlanta (CHOA) were obtained under an approved IRB protocol STUDY00001561 and a Data Transfer Agreement between CHOP and CHOA. The independent framework evaluation used examinations from patients diagnosed at Women’s and Children’s Hospital (WCH), Adelaide, Australia, under ethics approval 2022HRE00079 obtained from the Women’s and Children’s Health Network (WCHN) Human Research Ethics Committee. Data were selected and processed by Macquarie University (Sydney, Australia) under a Data Transfer Agreement with WCHN.

Subjects were eligible if they had a posterior fossa tumor and available MRI. Across the four sources, 1,181 subjects were identified; 170 were excluded for substantial motion artifact or preprocessing failure, leaving 1,011 subjects (Supplementary Fig. 1).

The in-distribution (ID) cohort comprised 775 treatment-naïve cases representing medulloblastoma (MB), pediatric low-grade glioma (LGG), diffuse midline glioma including diffuse intrinsic pontine glioma (DMG/DIPG), ependymoma (EPN), and atypical teratoid/rhabdoid tumor (ATRT). Diagnoses of medulloblastoma, LGG, EPN, and ATRT were confirmed histopathologically, as documented in CBTN records or patient charts. DMG/DIPG diagnoses were established by biopsy-based histopathology or radiologic assessment.

The 606 CBTN/CHOP cases were divided into discovery (n = 492) and replication (n = 114) cohorts using an approximately 80/20 split stratified by diagnosis. CHOA cases (n = 129) were reserved for external classifier evaluation, and Macquarie cases (n = 40) for independent evaluation of the complete framework. Cohort characteristics and diagnostic counts are provided in Table 1; scanner and acquisition characteristics extracted from T2-weighted DICOM headers are summarized in Supplementary Table 1b.

The out-of-distribution (OOD) cohort comprised 236 subjects: 22 patients with rare posterior fossa tumor histologies not represented during model training (biological OOD; Supplementary Table 1a) and 214 patients meeting one or more clinical OOD criteria. The clinical OOD criteria were prior treatment (n = 176), suboptimal image quality (n = 32), and tumor located outside the posterior fossa (n = 39); 33 subjects met two criteria. Previously treated examinations included 151 after resection and 25 after chemotherapy and/or radiotherapy without resection. These cohorts were used to characterize behavior outside the development distribution.

### Image preprocessing and model architecture

All imaging data were processed using our institutional pre-processing framework (https://github.com/d3b-center/peds-brain-seg-pipeline-public) ^29,30^, which integrates components from the Cancer Imaging Phenomics Toolkit (CaPTk, version 1.8.1; https://www.cbica.upenn.edu/captk) ^31,32^. MRI scans were first co-registered to the SRI24 atlas ^33^ with isotropic resampling to 1 mm³, providing a fixed input size of 240 × 240 × 155 voxels, then underwent skull stripping (https://github.com/d3b-center/peds-brain-auto-skull-strip) ^29,34^ and intensity normalization to a [0–255] range following removal of intensity outliers.

The posterior fossa region was defined anatomically in SRI24 space before model fitting and applied unchanged across cohorts. The expanded region occupied i ∈ [56, 184), j ∈ [100, 212), and k ∈ [0, 76), yielding a 128 × 112 × 76-voxel box. The adopted posterior fossa multi-crop configuration (PF_MC) extracted 18 views at offsets Δi, Δj ∈ {−8, 0, +8} and Δk ∈ {0, +8} voxels. Inferior translations were constrained by the image boundary at k = 0. Each crop was resized by trilinear interpolation to 112 × 112 × 112 voxels, matching the pretrained backbone input. These fixed offsets provided deterministic sampling rather than stochastic augmentation. The extraction regions and sampling envelope are illustrated in Supplementary Fig. 13.

Each resized crop was normalized independently: intensities were clipped at the 0.05th and 99.95th percentiles of its brain voxels and linearly rescaled to [−1, 1], with background mapped to −1. Brain voxels were defined as those exceeding 2% of the crop’s 99.5th intensity percentile. Computing percentiles over brain voxels avoided the predominance of zero-valued background in the posterior fossa region.

Features were extracted using 3DINO-ViT-L ^25^, a self-supervised vision transformer pretrained on approximately 100,000 three-dimensional scans from more than ten organs. Its stride-16 patch embedding produces 343 patch tokens for a 112³ input, with a token width of 1,024. The backbone remained frozen throughout. Final-block class (CLS) tokens were averaged across the 18 views to form one 1,024-dimensional subject-level representation. Features were standardized using training-set statistics and classified by a five-class multilayer perceptron (MLP) with two 256-unit hidden layers, GELU activations, dropout, and a final softmax output. Averaging occurred in feature space before classification.

### Model development and comparative evaluation

#### Model selection and internal validation

Model development used the discovery cohort exclusively. Candidate configurations shared five outer cross-validation folds; within each outer training fold, five-fold inner cross-validation selected hyperparameters by mean macro-AUC. The primary grid comprised six extraction regions, two representations, and five classification heads (60 combinations). Extraction strategies included the anatomically restricted posterior fossa box, a native 112³ posterior fossa cube requiring no resizing, and head-centered controls that excluded part of the inferior posterior fossa. Region definitions and comparisons are provided in Supplementary Table 29, Supplementary Note 10, and Supplementary Tables 30a,b.

The representations were the final-block CLS token (1,024 dimensions) and its concatenation with the mean patch token (2,048 dimensions; Supplementary Note 11 and Supplementary Table 31). Heads comprised a multiclass linear probe, multinomial logistic regression after standardization and principal-component analysis (PCA), a multiclass MLP with one or two hidden layers, and one-versus-rest linear and MLP configurations. Linear and MLP heads were trained for 300 epochs with AdamW and a cosine learning-rate schedule; logistic regression used L-BFGS. Hyperparameter search spaces and comparative results are provided in Supplementary Tables 32–33. A randomly initialized backbone served as a negative control (Supplementary Note 13 and Supplementary Table 34).

Selection proceeded sequentially: extraction region was compared across the ten representation–head combinations, representation within the selected region, and head within the selected region and representation. Multiplicity was controlled separately at each stage. A prespecified parsimony rule favored the lower-dimensional representation when differences were not significant. The MLP was chosen by overall rank rather than demonstrated superiority over the other multiclass heads. These comparisons therefore describe conditional selection stages rather than independent effects of each design component.

Replication results were not used for selection. Candidate configurations were evaluated on replication after their hyperparameters had been fixed using discovery; the deployed configuration was subsequently chosen by the discovery-only staged procedure. For deployment, separate non-nested five-fold cross-validation over a 32-point MLP grid selected a common configuration for all folds. The final model was refitted on all 492 discovery subjects and evaluated on the held-out cohorts.

#### Multi-crop sensitivity analysis and supervised benchmark

After fixing the region, representation, and head, we varied the number of views (1, 8, 18, or 50) at an 8-voxel maximum offset, and the maximum offset (4, 8, 12, or 16 voxels) at 18 views. In-plane positions were {0}, {−8, +8}, {−8, 0, +8}, or {−8, −4, 0, +4, +8}, respectively, with through-plane positions constrained to nonnegative offsets. The offset sweep used {−m, 0, +m} in-plane and {0, +m} through-plane. Each configuration used a newly fitted head with the same architecture, hyperparameters, and subject-level folds; standardization was fitted within each training fold. The single centered crop was analyzed separately from multi-view configurations.

The supervised benchmark was a MedicalNet-pretrained 3D ResNet-34 fine-tuned end-to-end on the same discovery subjects, folds, and preprocessing. It received the complete registered volume rather than posterior fossa crops. Five independent one-versus-rest classifiers were tuned by five-fold discovery cross-validation using validation AUC-ROC and then refitted on the complete discovery cohort. Class-specific temperatures were fitted to out-of-fold discovery predictions. Cases with no calibrated class probability ≥0.50 were abstained; otherwise, a softmax transformation of the five calibrated outputs was used to select the highest-scoring class and calculate confidence and certainty. Because the independent sigmoid outputs do not form a mutually exclusive distribution, primary benchmark comparisons used class-wise uncalibrated probabilities for both models, summarized by macro-averaging.

#### Calibration, uncertainty, and abstention

A single temperature T was fitted by minimizing negative log-likelihood on the selected configuration’s out-of-fold discovery predictions ^16^. For logit zₖ, calibrated probabilities were pₖ = exp(zₖ/T)/Σⱼ₌₁⁵ exp(zⱼ/T). The fitted temperature was applied unchanged to replication and both external cohorts. Calibration was assessed before and after scaling using reliability diagrams, top-label expected calibration error (ECE; ten equal-width bins), Brier score, and negative log-likelihood.

Prediction confidence was maxₖ pₖ. Normalized entropy was H = −Σₖ₌₁⁵ pₖ ln(pₖ)/ln(5), and distributional certainty was 1 − H. The top-two margin was the difference between the largest and second-largest probabilities. Distributional certainty summarizes concentration of the probability vector and does not have the frequency interpretation of a calibrated class probability.

The model assigned the most probable class when confidence was ≥0.50 and otherwise abstained. Confidence tiers were high (≥0.80), moderate (≥0.50 and <0.80), and abstained (<0.50). Abstained outputs retained class probabilities and uncertainty measures but did not assign a diagnostic label. Model weights, calibration, and the abstention threshold were fixed for held-out evaluation, with no refitting on external data.

### Distribution-shift and robustness analyses

#### Clinical and biological distribution shift

The frozen classifier was evaluated on clinical and biological OOD data without retraining or changing its decision rule. Twenty post-hoc OOD scores covered probability, logit, feature-distance, crop-disagreement, and hybrid approaches. Feature distances were computed in the 1,024-dimensional backbone and 256-dimensional penultimate-head spaces; crop disagreement used the 18 views before feature averaging. Larger scores consistently indicated greater shift. Score definitions and fixed parameters are provided in Supplementary Tables 35a,b.

All reference statistics—including class-specific feature statistics, nearest-neighbor banks, ReAct clipping thresholds, and ViM subspaces and scaling—were estimated from discovery training data. Feature-distance scores for replication, CHOA, and OOD subjects were averaged across five fold-specific discovery reference banks; scores requiring only deployment logits were computed directly from the frozen classifier. CHOA OOD and feature-space analyses included its 102 medulloblastomas for class-matched comparison; the remaining 27 cases contributed to classification evaluation only.

Discovery out-of-fold scores defined the ID reference distribution, and replication served as an independent ID control. Outcomes included OOD AUC-ROC, average precision, false-positive rate at 95% sensitivity, and the proportion exceeding the ID reference’s 95th percentile. Candidate scores were compared with maximum softmax probability. Score orientations were fixed before analysis; reverse separation (AUC-ROC <0.5) was reported separately rather than counted as successful detection.

#### Feature-space analysis

We characterized standardized backbone embeddings and penultimate-head activations using class centroids and a shared Ledoit–Wolf covariance estimated from discovery. Discovery subjects were scored against statistics from the other four folds. Mahalanobis distances were expressed as discovery-distribution percentiles. Class separability was measured using silhouette coefficients and the fraction of each subject’s 22 nearest neighbors sharing its class. Fold-respecting linear probes used the top k principal components ranked by explained variance or class-discriminative power, with PCA and rankings fitted within training folds. Geometric conclusions relied on the frozen space because the deployed head’s training-subject separability is optimistic.

#### Controlled imaging perturbations

Fifty replication cases were randomly selected with histologic stratification (ATRT, 5; DMG/DIPG, 7; EPN, 5; LGG, 14; MB, 19). TorchIO v0.21.3 ^21^ simulated ghosting and motion in k-space on unprocessed images. Ghosting used four ghosts along the phase-encoding axis at intensities 0.3, 0.6, and 0.9. Motion combined k-space from two rigid displacements with rotations/translations of 3, 6, or 10 degrees/mm. Parameters were fixed at each severity, with deterministic subject–condition seeds. Perturbed images underwent the same containerized preprocessing; negative intensities were set to zero after skull stripping and before normalization.

To assess classifier sensitivity to residual misalignment, rotations were applied after preprocessing, immediately before inference, about the left–right, anterior–posterior, and superior–inferior axes at ±5°, ±10°, ±15°, ±20°, ±30°, and ±45°. Rotations were image-centered, with linear interpolation, zero padding, and no translation or scaling. The 42 perturbations per subject produced 2,100 volumes. The deployed model, temperature, and threshold were unchanged, and rerunning unperturbed images reproduced all reference predictions.

Each condition was compared with the same subject’s unperturbed prediction. Outcomes were accuracy without abstention, abstention rate, accuracy among predicted cases, class-change frequency, and total-variation distance, ½Σₖ|pₖ − pₖ,ref|. The breaking angle was the smallest absolute rotation causing a class change or new abstention. Perturbation-induced errors were incorrect results among subjects correctly classified at baseline; the ability of confidence to identify incorrect results was assessed by AUC-ROC.

### Agentic workflow and application

#### Agent responsibilities and execution

The framework comprised a quality-control (QC) agent and a prediction agent (Fig. 7a), implemented through the Anthropic Messages API function-calling interface using Claude Opus 5 (model identifier claude-opus-5). Model settings and prompts were controlled by one versioned configuration (Supplementary Table 36). Agents received scalar measurements and categorical tool records, never images, and could not change the prescribed processing order. The QC agent invoked input inspection, acquisition QC, pipeline execution, and checks of registration, skull stripping, segmentation, containment, and normalization, within a 16-turn limit. It interpreted these records and assigned a QC state; the measurements themselves were computed by deterministic tools.

**Fig. 7.**
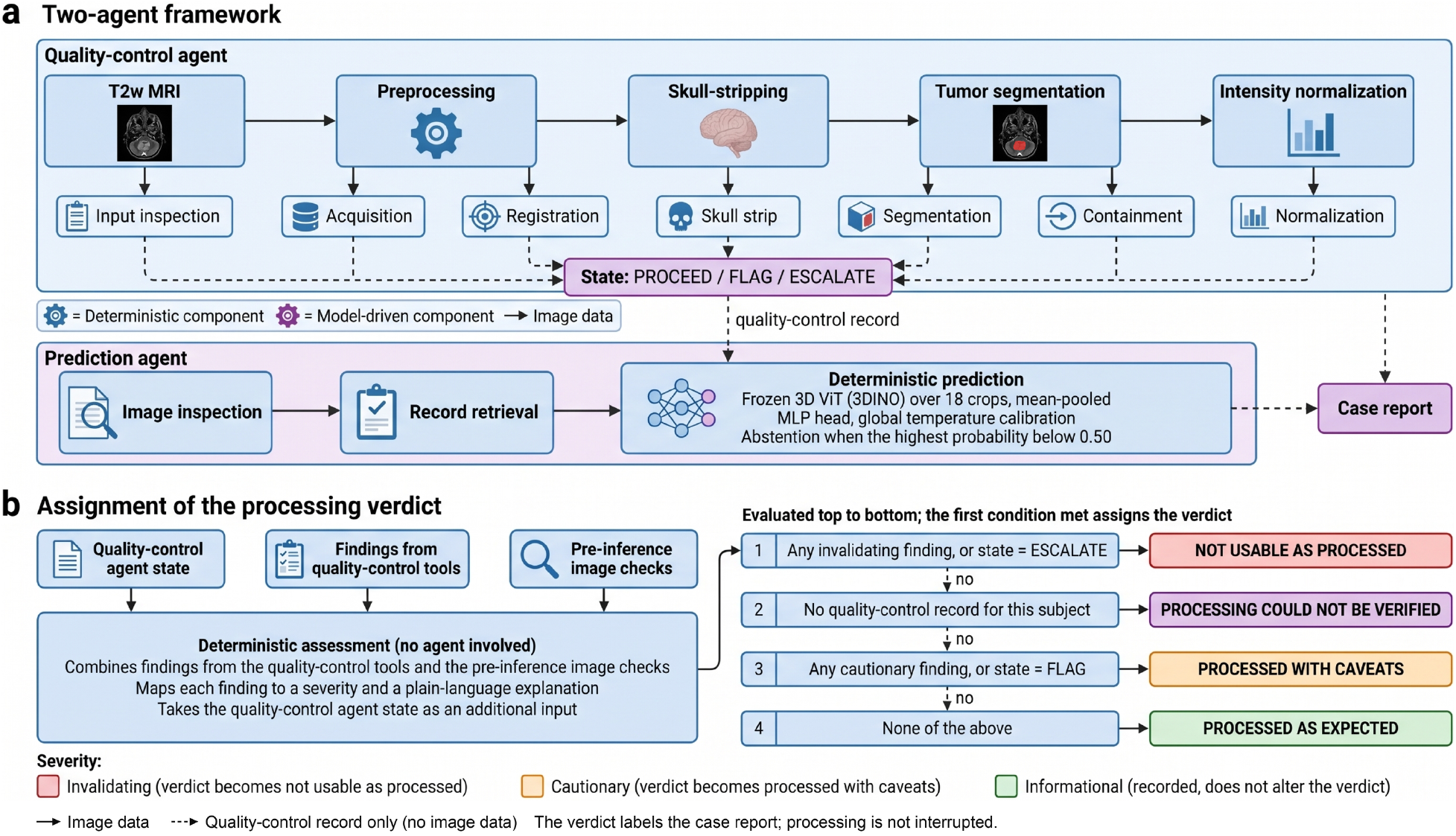
Two-agent framework and assignment of the processing verdict. **a**, Architecture of the framework. Upper lane: the quality-control agent invokes four containerized processing stages in fixed order (preprocessing, skull stripping, tumor segmentation, intensity normalization) and seven checks that assess their output. Each check returns a record of scalar measurements, from which the agent assigns a state of PROCEED, FLAG or ESCALATE. Lower lane: the prediction agent locates the preprocessed T2-weighted volume, reads the quality-control record and invokes the deterministic prediction routine, a frozen three-dimensional vision transformer with a calibrated multilayer perceptron head and an abstention threshold. Both agents contribute to a single case report. Box fill distinguishes deterministic components from the two values a model assigns. Solid arrows carry image data and dashed arrows carry scalar records only; no image data reach the model at any point. **b**, Assignment of the processing verdict. Findings raised by the quality-control tools and by the image checks performed immediately before inference are combined with the agent state in a single deterministic assessment involving neither agent. Each finding is mapped to one of three severity levels. One of four verdicts is then assigned by strict precedence, the first condition met determining the verdict, so that a single invalidating finding withholds the result irrespective of how many checks passed. Subjects with no quality-control record are assigned processing could not be verified. Informational findings are recorded and do not alter the verdict. The verdict labels the case report; processing was not interrupted, and every subject reaching inference received a prediction.

The prediction agent used image inspection, QC-record retrieval, and prediction tools, within an eight-turn limit. Before inference, the T2-weighted volume was checked for atlas geometry, 1-mm isotropic spacing, and LPS orientation. The agent invoked the classifier described above and reported its output without changing the class order, confidence values, or abstention decision. The agents supported orchestration and evidence reporting rather than diagnostic inference. Verbatim prompts are provided in Supplementary Note 16; tool definitions and thresholds are available in the code repository.

#### Quality-control measurements and states

Table 4 summarizes QC measurements and thresholds. For preprocessing segmentation, absent channels were populated by duplication of available T2-weighted or FLAIR images to satisfy the network’s four-channel input. In such cases, only whole-tumor volume was reported and tissue-specific volumes were withheld as non-interpretable. Subjects lacking both T2-weighted and FLAIR imaging were not processed; absence of T2-weighted imaging required escalation because the classifier uses that sequence. Available and duplicated channels were recorded.

**Table 4.** Quality-control checks, the measurements each returns, and the thresholds at which a finding was raised.

| **Check** | **Measurement** | **Finding raised at** |
|---|---|---|
| Input inspection | Sequences present; channels duplicated | Neither T2-weighted nor FLAIR present (not processed); T2-weighted absent (escalated) |
| Acquisition | Voxel dimensions; through-plane axis; plane | Two-dimensional acquisition in a non-axial plane |
| Registration | Rotation angle; translation magnitude | Within-subject: >10° or >15 mm. Subject-to-atlas: >25° |
| Skull stripping | Mask volume; large components; left-right asymmetry | <250 mL; more than one large component; asymmetry >0.15; >1500 mL only with a structural finding |
| Tumor segmentation | Label volumes; whole-tumor volume; large components | Empty; whole-tumor <0.5 mL; unexpected label values; more than one large component; edema ≥0.5 mL with tumor <0.5 mL |
| Containment | Fraction of tumor voxels outside the brain mask | >0.02 (0.005 to 0.02 recorded as minor) |
| Intensity normalization | Per-channel extrema and percentiles | Maximum >255; 99th minus 1st percentile <1 × 10<sup>−6</sup>; fewer than four channels |

FLAIR, fluid-attenuated inversion recovery.

Acquisitions were classified as two-dimensional when the largest voxel dimension exceeded 1.5 times the smallest; the orientation of that axis determined the acquisition plane. Non-axial two-dimensional acquisitions were flagged, whereas three-dimensional acquisitions were accepted in any orientation. Registration rotation was derived as arccos[(trace(R) − 1)/2] from the transform’s rotation submatrix, and translation as the vector norm. Translation >50 mm identified subject-to-atlas transforms; other transforms were treated as within-subject. Thresholds are listed in Table 4. Optimizer line-search warnings alone were not failures when transforms remained within bounds.

Brain-mask volumes were voxel counts multiplied by voxel volume. Components containing >5% of positive voxels were considered large; left–right asymmetry was the absolute hemispheric voxel-count difference divided by their sum. Volumes >1,500 mL were flagged only with fragmentation or asymmetry. Tumor labels followed our pediatric segmentation convention ^30,34^: enhancing, nonenhancing, and cystic components formed tumor core; edema was measured separately. Whole-tumor volume was the union of nonzero labels, and components containing >10% of tumor voxels were considered large. The edema-with-minimal-tumor check applied only when tissue-specific labels were interpretable. Containment used a brain mask on the segmentation grid; absence of a matching mask was flagged. Outside-brain fractions were calculated over segmentation-positive voxels. Normalization checks used nonzero-voxel extrema and percentiles for each channel.

The QC agent assigned PROCEED for satisfactory evidence, FLAG for usable outputs requiring review, or ESCALATE for failure or an unresolved high-consequence signal. Missing checks were treated as absent evidence, precluded PROCEED, and limited QC confidence to low. Otherwise, QC confidence was high for complete, concordant findings away from thresholds, moderate for borderline or partly conflicting findings, and low for provisional or conflicting evidence. This agent-assigned confidence was distinct from computed diagnostic confidence. The state, supporting findings, and evidence that could change the assessment were retained in the case record. The full finding and disposition mappings are provided in Supplementary Tables 37–38.

#### Processing verdict and reporting

A deterministic assessment combined preprocessing and pre-inference findings into a processing verdict (Fig. 7b), with findings mapped to three severity levels and four verdicts assigned by strict precedence. Missing QC records were labeled “processing could not be verified.” This verdict qualified interpretation of the case report; QC states did not interrupt inference or suppress generated outputs. Each subject reaching inference received a prediction or confidence-based abstention, accompanied by its QC findings.

The report included the processing verdict, measured findings and their consequences, stage-wise QC records, agent-assigned QC state and confidence, calibrated class probabilities, prediction confidence, distributional certainty, and abstention status. It also reported the runner-up class, top-two margin, and fraction of crops agreeing with the reported class. The prediction agent could not substitute its own confidence or diagnostic label.

The browser application submitted a subject identifier and axial T2-weighted MRI, with FLAIR, T1-weighted, and contrast-enhanced T1-weighted MRI optional (Fig. 8). Preprocessing containers, agents, and report generation executed asynchronously on the server, with progress and agent output streamed to the interface. Users could inspect preprocessed images and segmentation overlays in three planes alongside probabilities, QC findings, and the processing verdict. API credentials remained server-side.

**Fig. 8.**
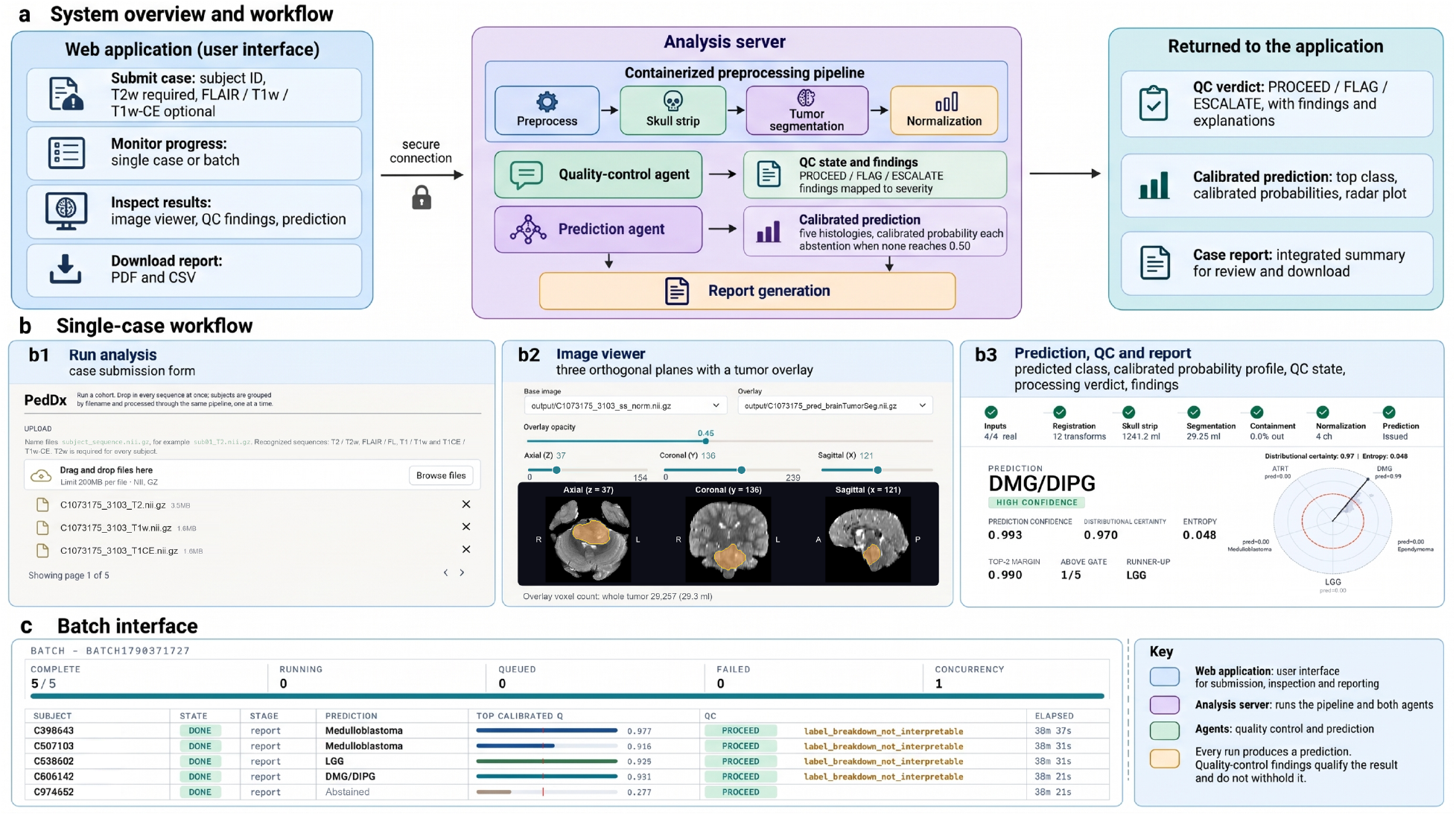
The PedDx application. **a**, System architecture. The browser submits cases and displays results; the containerized preprocessing pipeline, both agents and report generation execute server-side. The quality-control agent assigns a state and findings, the prediction agent reports the deterministic calibrated prediction, and both contribute to a single case report returned to the application. **b**, Single-case workflow. b1, case submission, with axial T2-weighted imaging required and FLAIR, T1-weighted and contrast-enhanced T1-weighted imaging optional. b2, image viewer for the preprocessed volume with the tumor segmentation overlaid, in axial, coronal and sagittal planes, with adjustable overlay opacity. b3, integrated view showing the predicted histology, the calibrated probability of each class, the quality-control state, the processing verdict, the findings raised, and access to the case report. **c**, Batch interface, showing per-subject state, stage, prediction, calibrated probability, quality-control state and findings across a cohort, with the results table and case reports available for download. Both agents execute server-side and the application does not expose the model API credential. Every run produces a result (a prediction or an abstention); the quality-control state and findings qualify the result and do not withhold it.

### Independent external evaluation

External evaluation separated classifier performance from independent execution of the complete framework. The development team evaluated the fixed classifier on CHOA data (n = 129). The Macquarie team independently selected and processed 40 examinations on its own infrastructure without involvement from the development team, assessing preprocessing, QC, inference, and reporting. Model weights, calibration, and the abstention rule were unchanged.

Evaluation summarized processing completion, QC-record availability, QC-state distribution, diagnostic accuracy before and after abstention, abstention frequency, and confidence-tier performance. At Macquarie, macro-averaged performance among predicted cases was calculated across MB, LGG, and DMG/DIPG, the three represented histologies; ATRT and EPN were absent. We also examined associations between QC state and correctness and compared prediction confidence, distributional certainty, and ordinal QC state as indicators of classification error.

### Statistical analysis

Discovery metrics were calculated within each fold and averaged, with standard deviations across folds; subject-level paired comparisons used pooled out-of-fold predictions. Table 2 reports class-wise sensitivity, specificity, F1, and balanced accuracy at a calibrated probability threshold of 0.50, whereas CHOA metrics in Table 3 use one-versus-rest labels derived from argmax predictions without abstention. Class-wise balanced accuracy was the mean of sensitivity and specificity, and macro averages weighted classes equally. Five-class accuracy and balanced accuracy were calculated from argmax labels. Performance after abstention used retained cases; abstention rates used the full evaluated cohort. Confidence intervals for CHOA accuracy were subject-level percentile bootstrap intervals (4,000 resamples), with accuracy after abstention recomputed on the cases retained in each resample. For the Macquarie evaluation (n = 40), where bootstrap percentile intervals reach 100%, Wilson score intervals were used. AUC-ROC used continuous class scores and the Mann–Whitney rank statistic with ties accounted for.

Paired subject-level bootstrap resampling estimated between-model differences, percentile 95% confidence intervals, and two-sided P values as twice the smaller tail proportion, bounded to [1/B, 1] for B resamples. Model-selection, multi-crop, and OOD comparisons used 2,000 resamples. Extraction regions were assessed by Friedman tests of within-context ranks across ten representation–head contexts, paired bootstraps within context, and two-sided sign tests of direction across contexts. Holm–Bonferroni adjustment was applied separately within each selection family; a difference required an adjusted P <0.05 and consistent direction across contexts. Multi-crop comparisons used the adopted 18-view, 8-voxel configuration as reference, with separate correction for view-count and offset sweeps.

ResNet comparisons used class-stratified paired bootstrap resampling (4,000 resamples) of uncalibrated out-of-fold scores, paired DeLong tests for per-class AUC-ROC, and exact McNemar tests for argmax accuracy. Benjamini–Hochberg adjustment was applied within each comparison table and reported as q values. OOD separation from discovery used two-sided Mann–Whitney U tests against AUC-ROC = 0.5, with Holm adjustment across 20 scores within each stratum. Reference and evaluation subjects were resampled independently for OOD confidence intervals; score comparisons shared resampled individuals. Comparisons with maximum softmax probability were Holm-adjusted within stratum, and class-matched comparisons within cell.

Perturbation outcomes used subject-level bootstrap confidence intervals (2,000 resamples). Correctness was compared with unperturbed predictions by exact two-sided McNemar tests, with Holm correction within ghosting, motion, or each rotation-axis family. Severity trends used logistic generalized estimating equations with exchangeable within-subject correlation and severity scaled from 0 to 1 (absolute angle/45° for rotation). In the independent framework evaluation, correctness for PROCEED versus other states was compared using Fisher’s exact test; error discrimination was summarized by AUC-ROC. All tests were two-sided with α = 0.05.

Training used Python 3.11.3, NumPy 1.26.4, and PyTorch 2.1.2; analyses used Python 3.13.9, NumPy 2.3.5, SciPy 1.16.3, scikit-learn 1.8.0, and PyTorch 2.10.0. Random processes used seed 42, with subject–condition seeds for perturbations as described above.

### Reporting summary

The TRIPOD+AI checklist is provided in Supplementary Note 17 (Supplementary Table 39).

## Data availability

Raw CBTN files can be accessed through https://cbtn.org/. CBTN processed and annotated data can be made available upon reasonable request to the corresponding author. The external data from Children’s Healthcare of Atlanta and Women’s and Children’s Hospital cannot be shared because of institutional data-sharing restrictions; requests may be directed to the respective institutions and are subject to ethics approval and a data transfer agreement.

## Author contributions

Conceptualization: Anahita Fathi Kazerooni, Ali Nabavizadeh, Arastoo Vossough, Fadel Batal; Methodology: Anahita Fathi Kazerooni, Fadel Batal; Computer programming: Fadel Batal, Anahita Fathi Kazerooni, Dimosthenis Chrysochoou, Meen Chul Kim, Celine Lee; Software: Fadel Batal, Deep Gandhi; Statistical analysis: Anahita Fathi Kazerooni, Fadel Batal; Interpretation of the data: Anahita Fathi Kazerooni, Fadel Batal, Ali Nabavizadeh, Arastoo Vossough; Validation: Anahita Fathi Kazerooni, Fadel Batal, Amin Tavallaii, Ghasem Azemi; Analysis: Anahita Fathi Kazerooni, Fadel Batal, Pengjia Zhu, Andrew Chiang, Deep Gandhi, Amin Tavallaii; Data collection: Anahita Fathi Kazerooni, Fadel Batal, Bhavyasri Vunnava, Eswar Damaraju, Anurag Gottipati, Jordan R. Hansford, Mikaela Doig, Amanda Luck, Sanaz Varshochi, Sahand Adib Moradi, Ariana M. Familiar, Neda Khalili, Nastaran Khalili, Paarth Jain, Yog Vaghani, Kartik Reddy, Arastoo Vossough; Data curation: Fadel Batal, Bhavyasri Vunnava, Deep Gandhi, Amin Tavallaii, Minh-Son To, Amanda Luck, Mikaela Doig; Funding acquisition: Adam C. Resnick, Phillip B. Storm, Antonio Di Ieva, Jordan R. Hansford, Mikaela Doig; Supervision: Anahita Fathi Kazerooni, Ali Nabavizadeh, Antonio Di Ieva, Jordan R. Hansford, Kartik Reddy; Writing & editing: all authors.

## Code availability

The PedDx application code, including the agent configuration, prompts and tool definitions, will be made publicly available at https://github.com/d3b-center/PedDx upon publication.

## Acknowledgements

National Institutes of Health (NIH) grants 75N91019D00024, Supplement 3U2CHL156291-03S2.

Work performed by the Australian team was supported by the Tour de Cure Grant Program 2023 (RSP-440-2024) and a Firefly Children’s Foundation 2026 Grant (26-50119473). JRH is supported partially through the Hospital Research and Jamie McClurg Foundations.

