## Supplementary Information for "PedDx: An End-to-End Deep Learning Framework for Pediatric Posterior Fossa Tumor Diagnosis with Agentic AI–Enabled Independent External Evaluation"

[1] Center for Data-Driven Discovery in Biomedicine (D3b), The Children's Hospital of Philadelphia, Philadelphia, PA, USA. [2] Division of Neurosurgery, The Children's Hospital of Philadelphia, Philadelphia, PA, USA. [3] Department of Neurosurgery, Perelman School of Medicine, University of Pennsylvania, Philadelphia, PA, USA. [4] Department of Radiology and Imaging Sciences, Emory University School of Medicine, Atlanta, GA, USA. [5] Department of Pediatrics, Emory University School of Medicine, Atlanta, GA, USA. [6] Computational NeuroSurgery (CNS) Lab, Macquarie Medical School, Faculty of Medicine, Health and Human Sciences, Macquarie University, Sydney, Australia. [7] Michael Rice Centre for Haematology and Oncology, Women's and Children's Hospital, Adelaide, SA, Australia. [8] Paediatric Neuro-Oncology, Precision Cancer Medicine, South Australian Health and Medical Research Institute, Adelaide, SA, Australia. [9] Flinders Health and Medical Research Institute, Flinders University, Bedford Park, SA, Australia. [10] South Australia Medical Imaging, Flinders Medical Centre, Adelaide, SA, Australia. [11] South Australian ImmunoGENomics Cancer Institute, Adelaide University, Adelaide, SA, Australia. [12] Department of Neurosurgery, Nepean Hospital, Kingswood, New South Wales, Australia. [13] Department of Radiology, Children's Healthcare of Atlanta, Atlanta, GA, USA. [14] Department of Radiology, Children's Hospital of Philadelphia, Philadelphia, PA, USA. [15] Department of Radiology, Perelman School of Medicine, University of Pennsylvania, Philadelphia, PA, USA.

#### Contents

Supplementary Figs. 1–13; Supplementary Tables 1–39 (including 1a,b; 2a,b; 30a,b; 35a,b); Supplementary Notes 1–17.


- Supplementary Note 1: additional cohort description (Supplementary Fig. 1 and Supplementary Tables 1a,b)
- Supplementary Note 2: sensitivity to multi-crop sampling geometry (Supplementary Tables 2a,b)
- Supplementary Note 3: comparison with a supervised convolutional baseline (Supplementary Tables 3–10)
- Supplementary Note 4: probability calibration and uncertainty quantification (Supplementary Figs. 2–3)
- Supplementary Note 5: external CHOA cohort, five-class results (Supplementary Table 11)
- Supplementary Note 6: behavior under distribution shift (Supplementary Tables 12–21, Supplementary Figs. 4–7)
- Supplementary Note 7: geometry of the frozen representation (Supplementary Tables 22–25, Supplementary Fig. 8)
- Supplementary Note 8: robustness to simulated imaging perturbations (Supplementary Tables 26–27, Supplementary Figs. 9–10)
- Supplementary Note 9: independent evaluation at Macquarie University (Supplementary Table 28, Supplementary Figs. 11–12)
- Supplementary Note 10: anatomical region and multi-crop sampling (Supplementary Tables 29 and 30a,b, Supplementary Fig. 13)
- Supplementary Note 11: final-block [CLS] versus pooled patch-token representations (Supplementary Table 31)
- Supplementary Note 12: classification head comparison (Supplementary Tables 32–33)
- Supplementary Note 13: contribution of pretraining (Supplementary Table 34)
- Supplementary Note 14: post-hoc out-of-distribution scores (Supplementary Tables 35a,b)
- Supplementary Note 15: agentic framework configuration (Supplementary Tables 36–38)
- Supplementary Note 16: agent system prompts
- Supplementary Note 17: reporting checklist, TRIPOD+AI (Supplementary Table 39)
- Supplementary References

### Supplementary Note 1 | Additional cohort description

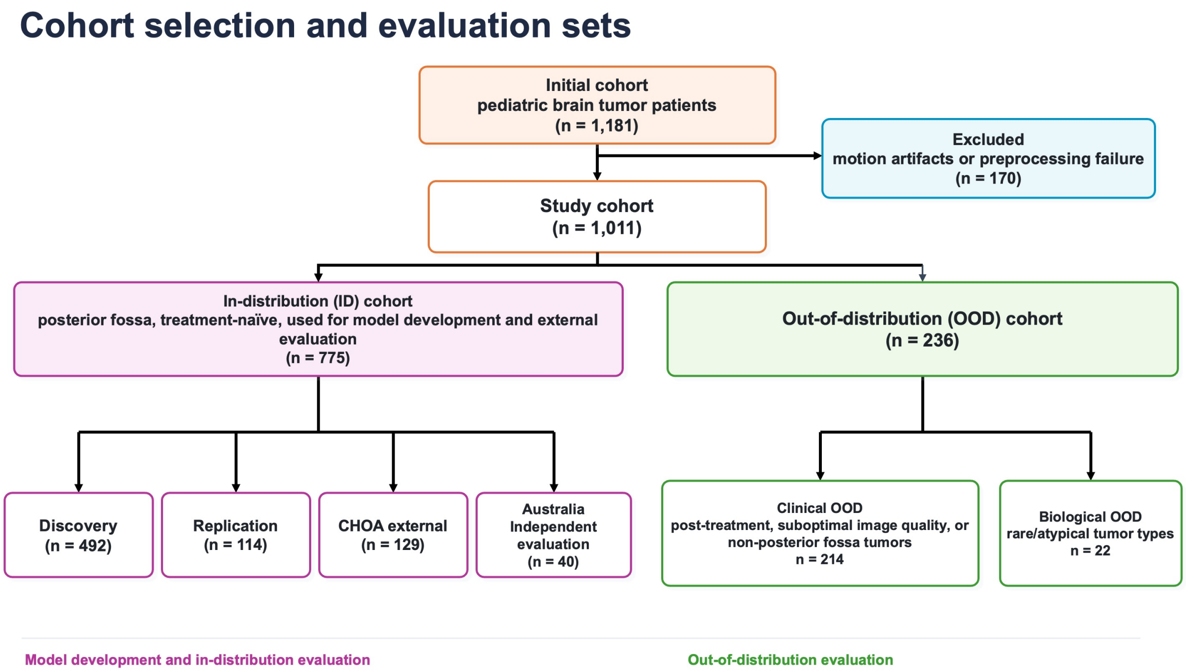


**Supplementary Fig. 1 | Study cohort flow diagram.** Composition of the full study cohort (n = 1,011) following exclusion of cases with motion artifacts or preprocessing failure (n = 170). The in-distribution cohort (n = 775) was subdivided into discovery (n = 492), replication (n = 114), external (CHOA, n = 129) and independent evaluation (Macquarie University, n = 40) sets. The out-of-distribution cohort (n = 236) was divided into clinical OOD (n = 214) and biological OOD (n = 22) subgroups.

The OOD cohort comprised 236 subjects and was used to characterize model behavior under clinically and biologically distinct conditions outside the model development distribution rather than to establish expected diagnostic performance in these settings. The biological OOD subgroup (n = 22) consisted of posterior fossa tumor entities not represented during model training (Supplementary Table 1a). The clinical OOD subgroup comprised 214 subjects meeting one or more predefined criteria: previously treated imaging (n = 176), suboptimal image quality (n = 32), or tumor extent not confined to the posterior fossa (n = 39). Because these categories were not mutually exclusive, 33 subjects (15%) satisfied two of the three clinical OOD criteria. Among the previously treated cases, 151 examinations were obtained after surgical resection, including MB (n = 93), LGG (n = 23), ATRT (n = 1), and EPN (n = 34), thereby introducing postoperative changes such as resection cavities and altered anatomy. An additional 25 examinations were obtained after chemotherapy and/or radiotherapy without surgical resection, including DMG/DIPG (n = 22), ATRT (n = 1), LGG (n = 1), and EPN (n = 1).

**Supplementary Table 1a | Patient characteristics of the out-of-distribution cohort.**

| Characteristic | Out-of-distribution cohort |
|---|---|
| Overall, n | 236 |
| Age, years, median (IQR) | 8.0 (4.4-13.0) |
| **Sex, n (%)** | |
| Male | 138 (58.5%) |

| Characteristic | Out-of-distribution cohort |
| --- | --- |
| Female | 98 (41.5%) |
| **Histologic diagnosis, n** | |
| Medulloblastoma | 104 |
| Ependymoma | 41 |
| DMG/DIPG | 31 |
| LGG | 27 |
| ATRT | 11 |
| Rare histologies (biological OOD) | 22 |
| **Rare histology breakdown, n** | |
| Hemangioblastoma | 6 |
| Ganglioglioma | 4 |
| Embryonal tumor with multilayered rosettes (ETMR) | 2 |
| Ewing sarcoma | 2 |
| Glial-neuronal tumor, NOS | 1 |
| Schwannoma | 1 |
| Germinoma | 1 |
| Cavernoma | 1 |
| Metastatic secondary tumor | 1 |
| Langerhans cell histiocytosis | 1 |
| Craniopharyngioma seeding | 1 |
| Neuroblastoma seeding | 1 |
| **Reason for clinical distribution shift, n** | |
| Post-therapy/post-operative imaging | 176 |
| Tumor not confined to the posterior fossa | 39 |
| Suboptimal image quality | 32 |
| ≥2 clinical OOD criteria | 33 |

Values are n unless otherwise indicated. The OOD cohort comprised 236 cases. Histologic diagnosis reflects the underlying tumor type and is mutually exclusive. The biological OOD subgroup comprised 22 cases with rare histologies. Clinical OOD criteria were assessed among the clinical OOD subgroup (n = 214) and were not mutually exclusive; 33 cases met more than one clinical OOD criterion. Abbreviations: ATRT, atypical teratoid/rhabdoid tumor; DMG/DIPG, diffuse midline glioma/diffuse intrinsic pontine glioma; ETMR, embryonal tumor with multilayered rosettes; LGG, low-grade glioma; NOS, not otherwise specified; OOD, out-of-distribution.

Scanner and acquisition characteristics of the discovery, replication and CHOA cohorts are given in Supplementary Table 1b.

**Supplementary Table 1b | Scanner and acquisition characteristics of the discovery, replication and CHOA cohorts**, from the DICOM header of the T2-weighted series used for inference. T2 geometry is summarized over 480 discovery and 114 replication examinations. Other vendors in discovery: Hitachi (n = 2, one at 0.7 T) and Toshiba (n = 1). Counts exclude examinations with missing values and may therefore sum to less than the number of examinations.

| Characteristic | Discovery (n = 492) | Replication (n = 114) | CHOA (n = 129) |
| --- | --- | --- | --- |
| Scanner information available, n | 490 | 114 | 129 |
| Vendor: Siemens / GE / Philips / other, n | 390 / 81 / 15 / 3 | 89 / 21 / 4 / 0 | 108 / 12 / 9 / 0 |
| Field strength: 1.5 T / 3 T / 0.7 T, n | 257 / 232 / 1 | 60 / 54 / 0 | 67 / 62 / 0 |
| 3 T, % | 47.3 | 47.4 | 48.1 |
| Scanner models, n | 37 | 23 | 20 |
| T2 slice thickness, mm, median (IQR) | 3.0 (2.0–4.0) | 3.0 (2.0–4.0) | 4.0 (3.0–5.0) |
| T2 in-plane spacing, mm, median (IQR) | 0.52 (0.45–0.66) | 0.53 (0.47–0.72) | 0.57 (0.47–0.63) |

| | | | |
|---|---|---|---|
| T2 acquisition: 2D / 3D, n | 405 / 72 | 92 / 20 | 124 / 5 |

#### Supplementary Note 2 | Sensitivity to multi-crop sampling geometry

**Methods.** After the extraction region, feature representation and classification head had been fixed, the sensitivity of the adopted PF_MC configuration to sampling geometry was evaluated within the discovery cohort. The adopted configuration used 18 views with a maximum translational offset of 8 voxels.

The MLP architecture and hyperparameters were held fixed, and a new head was fitted within the same folds for each configuration. Differences in macro-AUC relative to the adopted configuration were evaluated by subject-level paired bootstrap (ref. S16) with Holm adjustment (ref. S19) within the view-count and offset analyses; the single centered crop was treated as a separate comparison of single- versus multi-crop sampling.

**Results.** At the adopted configuration, multi-crop sampling improved discrimination relative to a single centered crop (Supplementary Table 2a). The adopted 18-view configuration achieved a macro-AUC of 0.8995 compared with 0.8769 for a single crop (Δ = +0.0226, 95% CI 0.006–0.040; Holm-adjusted p = 0.024).

**Supplementary Table 2a | Sensitivity to number of sampled views.** Discovery out-of-fold performance for the adopted PF_MC/final-block [CLS]/multiclass-MLP configuration. The maximum translational offset was held at 8 voxels while the number of sampled views was varied. Differences in macro-AUC were evaluated using 2,000 subject-level paired bootstrap resamples relative to the adopted 18-view configuration, with Holm adjustment across comparisons.

| Number of views | Macro-AUC | 95% CI | Δ vs 18 views | 95% CI of Δ | Holm-adjusted p |
|---|---|---|---|---|---|
| 1 (single crop) | 0.8769 | 0.8492–0.9026 | -0.0226 | -0.0397 to -0.0060 | 0.024 |
| 8 | 0.8985 | 0.8737–0.9213 | -0.0009 | -0.0068 to 0.0054 | 0.744 |
| **18 (adopted)** | **0.8995** | **0.8734–0.9232** | | **Reference** | |
| 50 | 0.9022 | 0.8776–0.9250 | +0.0028 | -0.0005 to 0.0061 | 0.204 |

Among multi-crop configurations, performance was stable across the number of sampled views. Eight views yielded a macro-AUC of 0.8985, which did not differ significantly from 18 views (Δ = −0.0009; Holm-adjusted $p = 0.744$). Increasing to 50 views yielded a macro-AUC of 0.9022 and did not significantly improve performance relative to 18 views (Δ = +0.0028, 95% CI −0.001 to +0.006; Holm-adjusted $p = 0.204$), despite requiring approximately 2.8-fold more crop extractions.

Performance was similarly stable across the tested translational offsets when the number of views was fixed at 18. Macro-AUCs were 0.8962, 0.8995, 0.8972 and 0.8962 for offsets of 4, 8, 12 and 16 voxels, respectively. Each alternative differed from the adopted 8-voxel offset by no more than 0.0032 macro-AUC, with no significant pairwise differences after Holm correction ($p = 0.590$ for each comparison).

**Supplementary Table 2b | Sensitivity to translational offset magnitude.** Discovery out-of-fold performance for the adopted PF_MC/final-block [CLS]/multiclass-MLP configuration. The number of views was fixed at 18 while the translational offset magnitude was varied. Differences in macro-AUC were evaluated using 2,000 subject-level paired bootstrap resamples relative to the adopted 8-voxel configuration, with Holm adjustment across comparisons.

| Offset magnitude, voxels | Macro-AUC | 95% CI | Δ vs 8 voxels | 95% CI of Δ | Holm-adjusted p |
|---|---|---|---|---|---|
| 4 | 0.8962 | 0.8705–0.9190 | -0.0032 | -0.0120 to 0.0050 | 0.590 |
| **8 (adopted)** | **0.8995** | **0.8761–0.9236** | | **Reference** | |
| 12 | 0.8972 | 0.8732–0.9194 | -0.0023 | -0.0098 to 0.0057 | 0.590 |
| 16 | 0.8962 | 0.8695–0.9196 | -0.0032 | -0.0149 to 0.0089 | 0.590 |

These results support the use of multi-crop sampling while indicating limited sensitivity to the exact sampling density or translational offset within the tested range. The 18-view, 8-voxel configuration was therefore retained for the final model.

#### Supplementary Note 3 | Comparison with a supervised convolutional baseline

**Methods.** For the supervised deep learning benchmark, a 3D ResNet-34 (ref. S24) initialized with MedicalNet (ref. S25) pretrained weights was fine-tuned end-to-end using the same discovery cohort, cross-validation assignments, and image preprocessing pipeline. In contrast to the 3DINO-ViT (ref. 26 of the main text; vision transformer, ref. S26) model, the ResNet-34 received the complete registered 240 × 240 × 155 voxel volume and did not require posterior fossa cropping.

All values in this Note were computed from uncalibrated (raw) probabilities for both models, because the baseline's five independent sigmoid outputs have no common calibrated scale. Calibrated 3DINO values, which are reported in the main text, are given in Table 2 of the main text. The comparison with the ResNet-34 baseline used uncalibrated probabilities for both models, a subject-level paired bootstrap stratified by class (4,000 resamples) of per-class and macro-averaged AUC-ROC and average precision on pooled discovery out-of-fold predictions, paired DeLong tests (ref. S17) for per-class AUC-ROC and an exact McNemar test (ref. S18) for argmax accuracy; P values were adjusted with the Benjamini–Hochberg procedure (ref. S20) within each table and are reported as q

**Supplementary Table 3 | Output-structure diagnostics.** Range and mean of the sum of the five class probabilities, and proportion of subjects with zero, one, or two or more classes at or above 0.5.

| Cohort | Model | n | Sum of probabilities, range | Mean | 0 classes ≥ 0.5 | 1 class ≥ 0.5 | 2+ classes ≥ 0.5 |
|---|---|---|---|---|---|---|---|
| **Discovery** | PedDx v2 (3DINO) | 492 | 1.000 – 1.000 | 1.000 | 13.4% | 86.6% | 0.0% |
| **Discovery** | PedDx v1 (ResNet) | | 0.262 – 3.633 | 1.796 | 7.3% | 27.2% | 65.4% |
| **Replication** | PedDx v2 (3DINO) | 114 | 1.000 – 1.000 | 1.000 | 13.2% | 86.8% | 0.0% |
| **Replication** | PedDx v1 (ResNet) | | 0.022 – 2.505 | 1.048 | 16.7% | 71.9% | 11.4% |

**Supplementary Table 4 | Per-class AUC-ROC on discovery for both models, mean-of-fold and pooled.** Mean-of-fold computes the metric within each fold and averages; pooled computes one metric over all out-of-fold predictions. Pooled Δ is PedDx v2 minus PedDx v1; q, Benjamini–Hochberg-adjusted P from the subject-level paired bootstrap; Better, the model favored at q < 0.05**.**

| Class | PedDx v2 (3DINO), mean-of-fold | PedDx v2 (3DINO), pooled | PedDx v1 (ResNet), mean-of-fold | PedDx v1 (ResNet), pooled | Pooled Δ (v2 − v1) | q | Better |
|---|---|---|---|---|---|---|---|
| Macro (one-vs-rest) | 0.901 | 0.900 | 0.888 | 0.777 | +0.123 | 0.0007 | 3DINO |
| ATRT | 0.899 | 0.897 | 0.902 | 0.689 | +0.208 | 0.0007 | 3DINO |
| DMG | 0.942 | 0.941 | 0.954 | 0.908 | +0.033 | 0.0911 | — |
| Ependymoma | 0.809 | 0.810 | 0.759 | 0.569 | +0.241 | 0.0010 | 3DINO |
| LGG | 0.933 | 0.933 | 0.934 | 0.856 | +0.077 | 0.0010 | 3DINO |
| Medulloblastoma | 0.921 | 0.920 | 0.889 | 0.862 | +0.058 | 0.0007 | 3DINO |

**Supplementary Table 5 | Median predicted probability for each class within each discovery fold, by model.** Shaded rows give the spread across folds (largest minus smallest median). PedDx v2, 3DINO; PedDx v1, ResNet-34.

| Model | Fold | ATRT | DMG | Ependymoma | LGG | Medulloblastoma |
|---|---|---|---|---|---|---|
| **PedDx v2** | fold 1 | 0.0063 | 0.0120 | 0.0098 | 0.1071 | 0.1971 |
| **PedDx v2** | fold 2 | 0.0070 | 0.0175 | 0.0212 | 0.0454 | 0.2912 |
| **PedDx v2** | fold 3 | 0.0109 | 0.0347 | 0.0212 | 0.0649 | 0.1872 |

| Model | Fold | ATRT | DMG | Ependymoma | LGG | Medulloblastoma |
|---|---|---|---|---|---|---|
| **PedDx v2** | fold 4 | 0.0078 | 0.0191 | 0.0125 | 0.1261 | 0.1982 |
| **PedDx v2** | fold 5 | 0.0103 | 0.0180 | 0.0215 | 0.0703 | 0.2634 |
| **PedDx v1** | fold 1 | 0.8086 | 0.0015 | 0.0000 | 0.3066 | 0.1533 |
| **PedDx v1** | fold 2 | 0.0000 | 0.0000 | 0.9922 | 0.0001 | 0.4883 |
| **PedDx v1** | fold 3 | 0.0002 | 0.1943 | 0.7500 | 0.3799 | 0.6074 |
| **PedDx v1** | fold 4 | 0.1890 | 0.0048 | 0.0450 | 0.0464 | 0.1450 |
| **PedDx v1** | fold 5 | 0.0392 | 0.0003 | 0.9844 | 0.7539 | 0.2119 |
| **PedDx v2** | spread (max − min) | 0.0046 | 0.0227 | 0.0118 | 0.0807 | 0.1040 |
| **PedDx v1** | spread (max − min) | 0.8086 | 0.1943 | 0.9922 | 0.7538 | 0.4624 |

**Supplementary Table 6 | Per-class one-vs-rest metrics on discovery, mean ± s.d.** across five folds. Δ is PedDx v2 (3DINO) minus PedDx v1 (ResNet-34), with a subject-level paired bootstrap of 2,000 resamples ; q, Benjamini–Hochberg-adjusted P across the comparison family.

| Class | Metric | PedDx v2 (3DINO) | PedDx v1 (ResNet-34) | Δ | Δ 95% CI | q | Better (q < 0.05) |
|---|---|---|---|---|---|---|---|
| ATRT | AUC-ROC | 0.899 ± 0.062 | 0.902 ± 0.067 | -0.003 | [-0.067, +0.061] | 0.9690 | — |
| ATRT | Average precision | 0.478 ± 0.196 | 0.452 ± 0.182 | +0.026 | [-0.108, +0.207] | 0.6426 | — |
| ATRT | Sensitivity | 0.200 ± 0.200 | 0.400 ± 0.469 | -0.200 | [-0.320, -0.080] | 0.0168 | PedDx v1 (ResNet) |
| ATRT | Specificity | 0.987 ± 0.009 | 0.806 ± 0.337 | +0.181 | [+0.156, +0.205] | 0.0010 | PedDx v2 (3DINO) |
| ATRT | PPV | 0.300 ± 0.298 | 0.107 ± 0.121 | +0.193 | [-0.005, +0.393] | 0.0913 | — |
| ATRT | F1 | 0.239 ± 0.237 | 0.138 ± 0.150 | +0.101 | [-0.039, +0.223] | 0.2647 | — |
| ATRT | Balanced accuracy | 0.594 ± 0.097 | 0.603 ± 0.133 | -0.009 | [-0.074, +0.056] | 0.7958 | — |
| DMG | AUC-ROC | 0.942 ± 0.058 | 0.954 ± 0.028 | -0.012 | [-0.038, +0.010] | 0.4186 | — |
| DMG | Average precision | 0.879 ± 0.083 | 0.843 ± 0.033 | +0.035 | [-0.033, +0.092] | 0.4706 | — |
| DMG | Sensitivity | 0.782 ± 0.107 | 0.620 ± 0.289 | +0.162 | [+0.087, +0.245] | 0.0010 | PedDx v2 (3DINO) |
| DMG | Specificity | 0.976 ± 0.019 | 0.951 ± 0.063 | +0.025 | [+0.003, +0.047] | 0.0415 | PedDx v2 (3DINO) |
| DMG | PPV | 0.867 ± 0.103 | 0.772 ± 0.167 | +0.095 | [-0.010, +0.241] | 0.1214 | — |
| DMG | F1 | 0.820 ± 0.093 | 0.638 ± 0.201 | +0.182 | [+0.106, +0.265] | 0.0010 | PedDx v2 (3DINO) |
| DMG | Balanced accuracy | 0.879 ± 0.058 | 0.786 ± 0.125 | +0.093 | [+0.053, +0.135] | 0.0010 | PedDx v2 (3DINO) |
| Ependymoma | AUC-ROC | 0.809 ± 0.088 | 0.759 ± 0.031 | +0.050 | [-0.023, +0.128] | 0.2739 | — |
| Ependymoma | Average precision | 0.374 ± 0.133 | 0.242 ± 0.094 | +0.132 | [+0.026, +0.257] | 0.0275 | PedDx v2 (3DINO) |
| Ependymoma | Sensitivity | 0.082 ± 0.075 | 0.629 ± 0.511 | -0.546 | [-0.632, -0.439] | 0.0010 | PedDx v1 (ResNet) |
| Ependymoma | Specificity | 0.998 ± 0.005 | 0.409 ± 0.540 | +0.589 | [+0.578, +0.598] | 0.0010 | PedDx v2 (3DINO) |
| Ependymoma | PPV | 0.500 ± 0.500 | 0.245 ± 0.423 | +0.255 | [-0.196, +0.554] | 0.4576 | — |
| Ependymoma | F1 | 0.139 ± 0.127 | 0.134 ± 0.089 | +0.005 | [-0.135, +0.136] | 0.9777 | — |
| Ependymoma | Balanced accuracy | 0.540 ± 0.037 | 0.519 ± 0.031 | +0.021 | [-0.026, +0.073] | 0.5060 | — |
| LGG | AUC-ROC | 0.933 ± 0.008 | 0.934 ± 0.028 | -0.001 | [-0.026, +0.026] | 0.9805 | — |
| LGG | Average precision | 0.882 ± 0.014 | 0.874 ± 0.045 | +0.008 | [-0.030, +0.048] | 0.7034 | — |
| LGG | Sensitivity | 0.734 ± 0.065 | 0.754 ± 0.261 | -0.020 | [-0.101, +0.057] | 0.6735 | — |
| LGG | Specificity | 0.907 ± 0.022 | 0.769 ± 0.217 | +0.138 | [+0.099, +0.177] | 0.0010 | PedDx v2 (3DINO) |
| LGG | PPV | 0.790 ± 0.043 | 0.697 ± 0.206 | +0.093 | [+0.034, +0.152] | 0.0047 | PedDx v2 (3DINO) |
| LGG | F1 | 0.760 ± 0.041 | 0.663 ± 0.103 | +0.096 | [+0.031, +0.159] | 0.0088 | PedDx v2 (3DINO) |
| LGG | Balanced accuracy | 0.821 ± 0.032 | 0.762 ± 0.073 | +0.059 | [+0.013, +0.103] | 0.0200 | PedDx v2 (3DINO) |
| Medulloblastoma | AUC-ROC | 0.921 ± 0.021 | 0.889 ± 0.017 | +0.032 | [+0.006, +0.058] | 0.0282 | PedDx v2 (3DINO) |
| Medulloblastoma | Average precision | 0.883 ± 0.041 | 0.825 ± 0.030 | +0.058 | [+0.013, +0.097] | 0.0200 | PedDx v2 (3DINO) |
| Medulloblastoma | Sensitivity | 0.811 ± 0.082 | 0.779 ± 0.163 | +0.032 | [-0.026, +0.084] | 0.3813 | — |
| Medulloblastoma | Specificity | 0.881 ± 0.021 | 0.821 ± 0.119 | +0.060 | [+0.016, +0.106] | 0.0183 | PedDx v2 (3DINO) |
| Medulloblastoma | PPV | 0.810 ± 0.034 | 0.760 ± 0.111 | +0.050 | [-0.003, +0.105] | 0.1076 | — |
| Medulloblastoma | F1 | 0.809 ± 0.050 | 0.751 ± 0.058 | +0.059 | [+0.014, +0.104] | 0.0200 | PedDx v2 (3DINO) |
| Medulloblastoma | Balanced accuracy | 0.846 ± 0.042 | 0.800 ± 0.043 | +0.046 | [+0.010, +0.081] | 0.0200 | PedDx v2 (3DINO) |

**Supplementary Table 7 | Macro one-vs-rest metrics on discovery, mean ± s.d. across five folds.** Δ is PedDx v2 (3DINO) minus PedDx v1 (ResNet).

| Metric | PedDx v2 (3DINO) | PedDx v1 (ResNet) | Δ | 95% CI | q | Better |
|---|---|---|---|---|---|---|
| **Balanced accuracy** | 0.736 ± 0.029 | 0.694 ± 0.041 | +0.042 | [+0.019, +0.066] | 0.0047 | PedDx v2 (3DINO) |
| **F1** | 0.553 ± 0.068 | 0.465 ± 0.054 | +0.088 | [+0.039, +0.134] | 0.0018 | PedDx v2 (3DINO) |
| **AUC-ROC** | 0.901 ± 0.026 | 0.888 ± 0.010 | +0.013 | [-0.010, +0.036] | 0.3327 | — |

| Metric | PedDx v2 (3DINO) | PedDx v1 (ResNet) | Δ | 95% CI | q | Better |
|---|---|---|---|---|---|---|
| **Sensitivity** | 0.522 ± 0.057 | 0.636 ± 0.172 | -0.115 | [-0.155, -0.072] | 0.0010 | PedDx v1 (ResNet) |
| **Specificity** | 0.950 ± 0.007 | 0.751 ± 0.135 | +0.198 | [+0.185, +0.212] | 0.0010 | PedDx v2 (3DINO) |
| **Average precision** | 0.699 ± 0.063 | 0.647 ± 0.040 | +0.052 | [+0.011, +0.097] | 0.0210 | PedDx v2 (3DINO) |
| **PPV** | 0.654 ± 0.151 | 0.516 ± 0.121 | +0.137 | [+0.034, +0.208] | 0.0096 | PedDx v2 (3DINO) |

**Supplementary Table 8 | Macro one-vs-rest metrics on discovery, pooled over all out-of-fold predictions.**

| Metric | PedDx v2 (3DINO) | PedDx v1 (ResNet) | Δ | 95% CI | q | Better |
|---|---|---|---|---|---|---|
| **Balanced accuracy** | 0.735 | 0.693 | +0.042 | [+0.010, +0.076] | 0.0139 | PedDx v2 (3DINO) |
| **F1** | 0.563 | 0.479 | +0.085 | [+0.028, +0.140] | 0.0057 | PedDx v2 (3DINO) |
| **AUC-ROC** | 0.900 | 0.777 | +0.123 | [+0.086, +0.159] | 0.0007 | PedDx v2 (3DINO) |
| **Sensitivity** | 0.521 | 0.635 | -0.114 | [-0.176, -0.052] | 0.0010 | PedDx v1 (ResNet) |
| **Specificity** | 0.950 | 0.751 | +0.199 | [+0.181, +0.217] | 0.0007 | PedDx v2 (3DINO) |
| **Average precision** | 0.664 | 0.515 | +0.149 | [+0.098, +0.209] | 0.0007 | PedDx v2 (3DINO) |
| **PPV** | 0.734 | 0.447 | +0.287 | [+0.137, +0.389] | 0.0007 | PedDx v2 (3DINO) |

**Results.** Paired DeLong tests on the pooled predictions agree with the bootstrap. An exact McNemar test on the argmax re-derivation gives 0.742 against 0.530, with 140 subjects correct for the frozen model alone and 36 for the baseline alone ($p < 0.0001$).

**Supplementary Table 9 | Paired DeLong and exact McNemar tests, discovery, pooled.** q applies to the DeLong family within the cohort.

| Target | PedDx v2 (3DINO) | PedDx v1 (ResNet) | Statistic | p | q |
|---|---|---|---|---|---|
| **ATRT AUC-ROC** | 0.897 | 0.689 | z = 3.35 | 0.0008 | 0.0010 |
| **DMG/DIPG AUC-ROC** | 0.941 | 0.908 | z = 1.84 | 0.0653 | 0.0653 |
| **Ependymoma AUC-ROC** | 0.810 | 0.569 | z = 4.02 | 0.0001 | 0.0001 |
| **LGG AUC-ROC** | 0.933 | 0.856 | z = 4.13 | 0.0000 | 0.0001 |
| **Medulloblastoma AUC-ROC** | 0.920 | 0.862 | z = 3.94 | 0.0001 | 0.0001 |
| **Argmax accuracy** | 0.742 | 0.530 | 3DINO-only correct 140, ResNet-only correct 36 | 0.0000 | — |

On the replication cohort each model is a single network with hyperparameters locked on discovery, so the between-fold inconsistency that drives the pooled discovery result cannot arise. Here the two models are indistinguishable: no per-class or macro difference survives false-discovery correction, and the exact McNemar test on argmax accuracy gives 0.798 against 0.833 in favor of the baseline ($p = 0.56$).

**Supplementary Table 10 | Macro one-vs-rest metrics on replication, a single held-out evaluation of one model per system.**

| Metric | PedDx v2 (3DINO) | PedDx v1 (ResNet-34) | Δ | p | q | Better |
|---|---|---|---|---|---|---|
| **Balanced accuracy** | 0.792 | 0.786 | +0.006 | 0.8290 | 0.9697 | — |
| **F1** | 0.666 | 0.618 | +0.048 | 0.3680 | 0.8192 | — |
| **AUC-ROC** | 0.951 | 0.903 | +0.048 | 0.0070 | 0.1027 | — |
| **Sensitivity** | 0.610 | 0.615 | -0.006 | 0.8970 | 0.9771 | — |
| **Specificity** | 0.974 | 0.957 | +0.017 | 0.1110 | 0.6105 | — |
| **Average precision** | 0.782 | 0.699 | +0.082 | 0.0370 | 0.3256 | — |
| **PPV** | 0.766 | 0.626 | +0.140 | 0.0880 | 0.5531 | — |

#### Supplementary Note 4 | Probability calibration and uncertainty quantification

Calibration results are reported in the main text; reliability diagrams and the distributions of confidence and entropy are shown in Supplementary Figs. 2 and 3.$m = p_{(1)}^{cal} - p_{(2)}^{cal} p_{(1)}^{cal} p_{(2)}^{cal}$

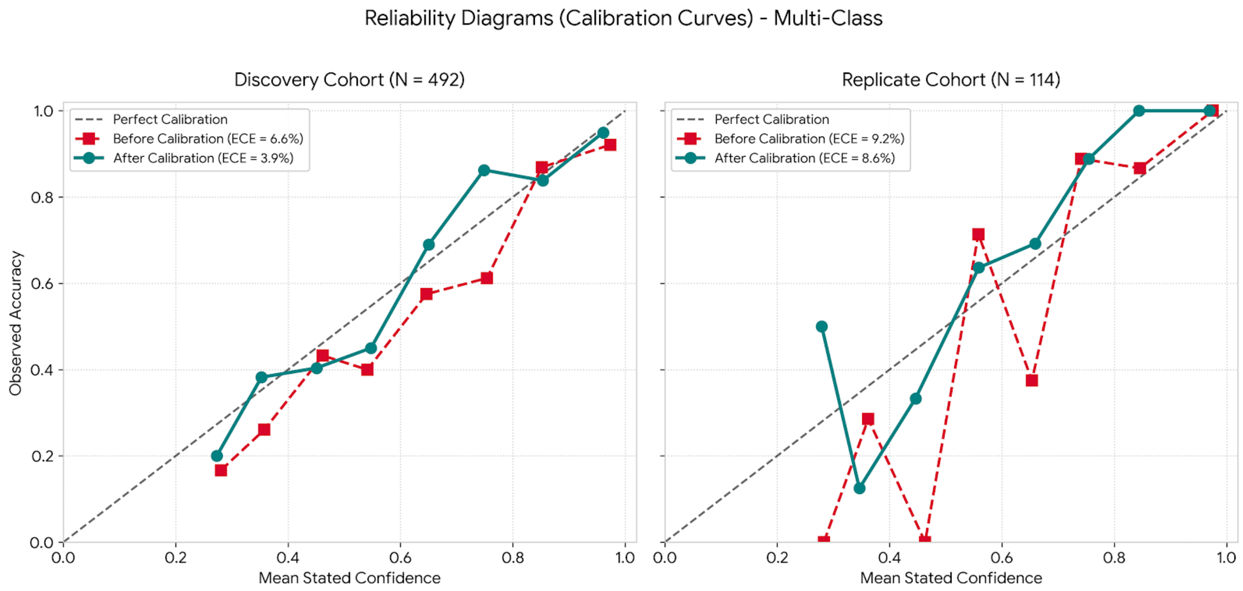


**Supplementary Fig. 2 | Multi-class reliability diagrams before and after temperature scaling. a**, discovery cohort (n = 492). Top-label observed accuracy plotted against mean stated probability across ten equal-width bins using pooled out-of-fold predictions before calibration (red dashed curve with square markers; ECE=6.6%) and after temperature scaling (teal solid curve with circular markers; ECE = 3.9%). **b**, replication cohort (n = 114). Top-label calibration performance evaluated on the replication set before calibration (red dashed curve; ECE = 9.2%) and after applying the fixed discovery-derived temperature parameter (T = 1.30, teal solid curve; ECE = 8.6%). The dashed grey diagonal line represents ideal calibration (y = x). ECE, expected calibration error.

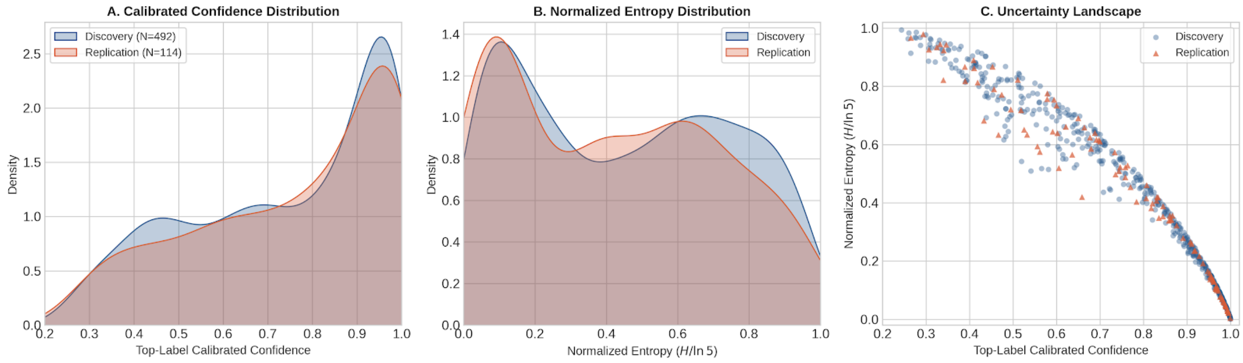


**Supplementary Fig. 3 | Comparative distributions of calibrated confidence, normalized entropy, and prediction uncertainty landscape. a**, Kernel density estimates of top-label calibrated confidence (maximum predicted class probability) for the discovery (n = 492, blue) and replication (n = 114, orange) cohorts. **b**, Kernel density estimates of normalized Shannon entropy (H / ln 5) evaluating overall probability distribution uncertainty across five target classes for the discovery (blue) and replication (orange) cohorts. **c**, Prediction uncertainty landscape displaying top-label calibrated confidence (x-axis) versus normalized entropy (y-axis) for discovery (blue circular markers) and replication (orange triangular markers) samples. Probabilities reflect post-temperature scaling (T = 1.30) model outputs.

#### Supplementary Note 5 | External CHOA cohort: five-class results

The external CHOA cohort comprised 129 patients: 102 with medulloblastoma and 27 with LGG (n = 13), DMG (n = 5), ATRT (n = 5) or ependymoma (n = 4). Overall results and the confusion matrix are given in Supplementary Table 11. Per-class results are in Table 3 of the main text.

**Supplementary Table 11 | Overall results on the CHOA cohort and by histology group.** Accuracy without abstention assigns every case to its most probable class; a case was abstained when its maximum calibrated probability was below 0.50. Values in brackets are percentages and subject-level bootstrap 95% confidence intervals (2,000 resamples). Errors intercepted, errors without abstention on which the model abstained. Lower panel: confusion matrix without abstention (rows, true histology; columns, predicted), with the number of abstained cases in brackets.

| Group | n | Correct without abstention, n (%; 95% CI) | Abstained, n (%) | Correct among predicted, n/N (%; 95% CI) | Errors intercepted |
|---|---|---|---|---|---|
| All (n = 129) | 129 | 105 (81.4; 74.4–87.6) | 23 (17.8) | 94/106 (88.7; 82.1–94.3) | 12/24 |
| Medulloblastoma | 102 | 89 (87.3; 80.4–93.1) | 16 (15.7) | 78/86 (90.7; 84.5–96.4) | 5/13 |
| Non-medulloblastoma | 27 | 16 (59.3; 40.7–77.8) | 7 (25.9) | 16/20 (80.0; 61.9–95.2) | 7/11 |

| True \ predicted | ATRT | DMG | Ependymoma | LGG | Medulloblastoma | Total |
|---|---|---|---|---|---|---|
| ATRT | 0 | 0 | 1 (1) | 1 (1) | 3 (2) | 5 |
| DMG | 0 | 5 (0) | 0 | 0 | 0 | 5 |
| Ependymoma | 1 (0) | 0 | 0 | 0 | 3 (1) | 4 |
| LGG | 0 | 1 (1) | 0 | 11 (0) | 1 (1) | 13 |
| Medulloblastoma | 0 | 0 | 2 (2) | 11 (3) | 89 (11) | 102 |

#### Supplementary Note 6 | Behavior under distribution shift

**Methods.** The OOD analyses are described in the main text (Methods). Score definitions and fixed parameters are given in Supplementary Note 14.

**Results.** Accuracy without abstention fell from 0.742–0.814 in the in-distribution cohorts to 0.381, 0.281 and 0.179 in the post-treatment, image-quality and not-PF-confined strata (Supplementary Table 12). Median confidence decreased less steeply, from 0.80–0.83 to 0.54–0.61, and abstention increased to 35.9–37.5%. Confidence therefore responded to shift, but not in proportion to the loss of accuracy.

**Supplementary Table 12 | Accuracy and confidence by cohort.** Confidence is the largest calibrated probability; distributional certainty is one minus the entropy normalized by its maximum. Accuracy is undefined for the biological stratum, whose histologies lie outside the label space. Predicted cases are those with a calibrated top-class probability ≥ 0.50.

| Cohort | n | Accuracy without abstention | Accuracy, predicted cases | Accuracy, abstained cases | Median confidence | Median distributional certainty | Median entropy | Abstained |
|---|---|---|---|---|---|---|---|---|
| **ID (CV)** | 492 | 0.742 | 0.831 | 0.375 | 0.802 | 0.555 | 0.716 | 19.5% |
| **ID (replication)** | 114 | 0.798 | 0.905 | 0.263 | 0.830 | 0.599 | 0.645 | 16.7% |
| **External (CHOA)** | 129 | 0.814 | 0.887 | 0.478 | 0.802 | 0.591 | 0.658 | 17.8% |
| **Post-treatment** | 176 | 0.381 | 0.391 | 0.364 | 0.551 | 0.258 | 1.195 | 37.5% |
| **Not PF-confined** | 39 | 0.179 | 0.240 | 0.071 | 0.608 | 0.291 | 1.140 | 35.9% |
| **Image quality** | 32 | 0.281 | 0.300 | 0.250 | 0.535 | 0.233 | 1.235 | 37.5% |
| **Biological OOD** | 22 | – | – | – | 0.573 | 0.332 | 1.074 | 31.8% |

We compared out-of-fold discovery scores with the independent replication cohort as a negative control to establish the expected baseline for in-distribution behavior. None of the evaluated scores significantly separated the two cohorts after Holm adjustment; the largest deviation from chance corresponded to an AUC-ROC of 0.447, and all adjusted P values exceeded 0.05.

**Supplementary Table 13 | Separation of the replication cohort from discovery by each post-hoc score (in-distribution control). An AUC-ROC of 0.5 indicates no separation; P values are Holm-adjusted across the 20 scores.**

| Score | AUC-ROC vs discovery | CI low | CI high | p | Holm P |
|---|---|---|---|---|---|
| **Rel. Mahalanobis (256-d)** | 0.447 | 0.397 | 0.496 | 0.077 | 0.868 |
| **Energy** | 0.453 | 0.396 | 0.511 | 0.117 | 0.868 |
| **Crop cosine spread** | 0.454 | 0.394 | 0.512 | 0.130 | 0.868 |
| **Crop dispersion** | 0.455 | 0.397 | 0.514 | 0.132 | 0.868 |
| **NNGuide** | 0.455 | 0.399 | 0.513 | 0.132 | 0.868 |
| **MaxLogit** | 0.457 | 0.400 | 0.516 | 0.157 | 0.868 |
| **Crop mutual information** | 0.459 | 0.401 | 0.514 | 0.174 | 0.868 |
| **GEN** | 0.459 | 0.400 | 0.518 | 0.176 | 0.868 |
| **ReAct + Energy** | 0.464 | 0.411 | 0.521 | 0.235 | 0.868 |
| **Entropy** | 0.465 | 0.406 | 0.521 | 0.250 | 0.868 |
| **MSP** | 0.471 | 0.411 | 0.531 | 0.326 | 0.868 |
| **MSP + kNN (rank avg)** | 0.474 | 0.416 | 0.534 | 0.386 | 0.868 |
| **kNN (256-d)** | 0.474 | 0.419 | 0.529 | 0.387 | 0.868 |
| **Crop argmax instability** | 0.474 | 0.419 | 0.532 | 0.377 | 0.868 |
| **ViM** | 0.477 | 0.419 | 0.535 | 0.441 | 0.868 |
| **kNN (1024-d)** | 0.495 | 0.438 | 0.551 | 0.860 | 0.868 |
| **Mahalanobis (1024-d)** | 0.505 | 0.449 | 0.563 | 0.868 | 0.868 |
| **Rel. Mahalanobis (1024-d)** | 0.517 | 0.468 | 0.572 | 0.564 | 0.868 |
| **Crop energy s.d.** | 0.535 | 0.474 | 0.594 | 0.246 | 0.868 |
| **Mahalanobis (256-d)** | 0.538 | 0.480 | 0.597 | 0.201 | 0.868 |

To characterize how distribution shift altered the outputs of the classifier per class, calibrated probabilities were examined separately for positive cases (true class matched the classifier) and negative cases (true class differed from the classifier). Biological OOD cases were negative for every trained classifier because their histologies were outside the five-class label space. Boxplots summarize cohort- and class-specific distributions (Supplementary Fig. 4), while kernel density estimates show the corresponding positive and negative distribution shapes (Supplementary Fig. 5).

Under in-distribution conditions, medulloblastoma, DMG/DIPG and LGG positive cases were concentrated toward high calibrated probabilities, while their negative distributions were predominantly near zero. Clinical OOD conditions primarily disrupted the positive distributions. Medulloblastoma probabilities shifted downward and broadened across post-treatment, non-posterior-fossa and image-quality cases. DMG/DIPG similarly showed a pronounced loss of the sharply high-probability in-distribution pattern in post-treatment cases, with probabilities extending across a substantially wider intermediate range. LGG positive cases remained comparatively elevated but became markedly broader under clinical shift.

ATRT and ependymoma exhibited greater positive-case dispersion already under in-distribution conditions, making the direction of change under shift less uniform than for medulloblastoma or DMG/DIPG. Across classifiers, negative-case probabilities generally remained concentrated at the low end of the scale, but several clinical OOD cohorts developed broader upper tails, consistent with increased susceptibility to off-target assignments. Biological OOD cases did not produce uniformly near-zero probabilities despite being negative for every trained class; the broadest intermediate distribution was observed for LGG, with additional non-zero tails for other classifiers. These patterns motivated evaluation of label-free post-hoc detection scores rather than relying on the calibrated class probability alone.

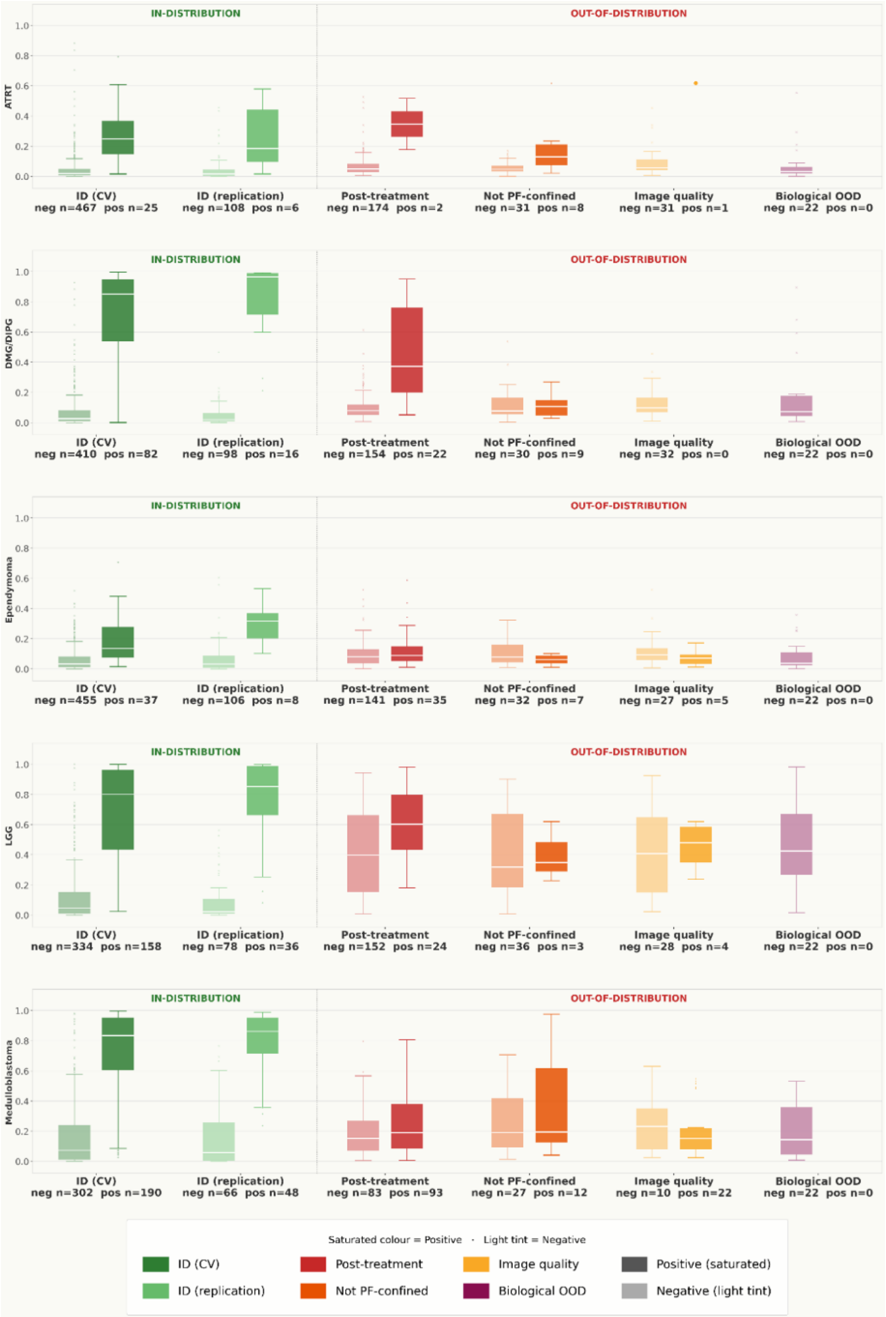


**Supplementary Fig. 4 | Class-specific calibrated probability distributions across in-distribution and shifted cohorts.** For each class, saturated boxes denote positive cases and lighter boxes denote negative cases. Discovery cross-validation and replication are shown as in-distribution controls; post-treatment, tumors not confined to the posterior fossa, inadequate image quality and biological OOD are shown as shifted cohorts. Biological OOD contains no positive cases for any trained classifier because its histologies lie outside the five-class label space.

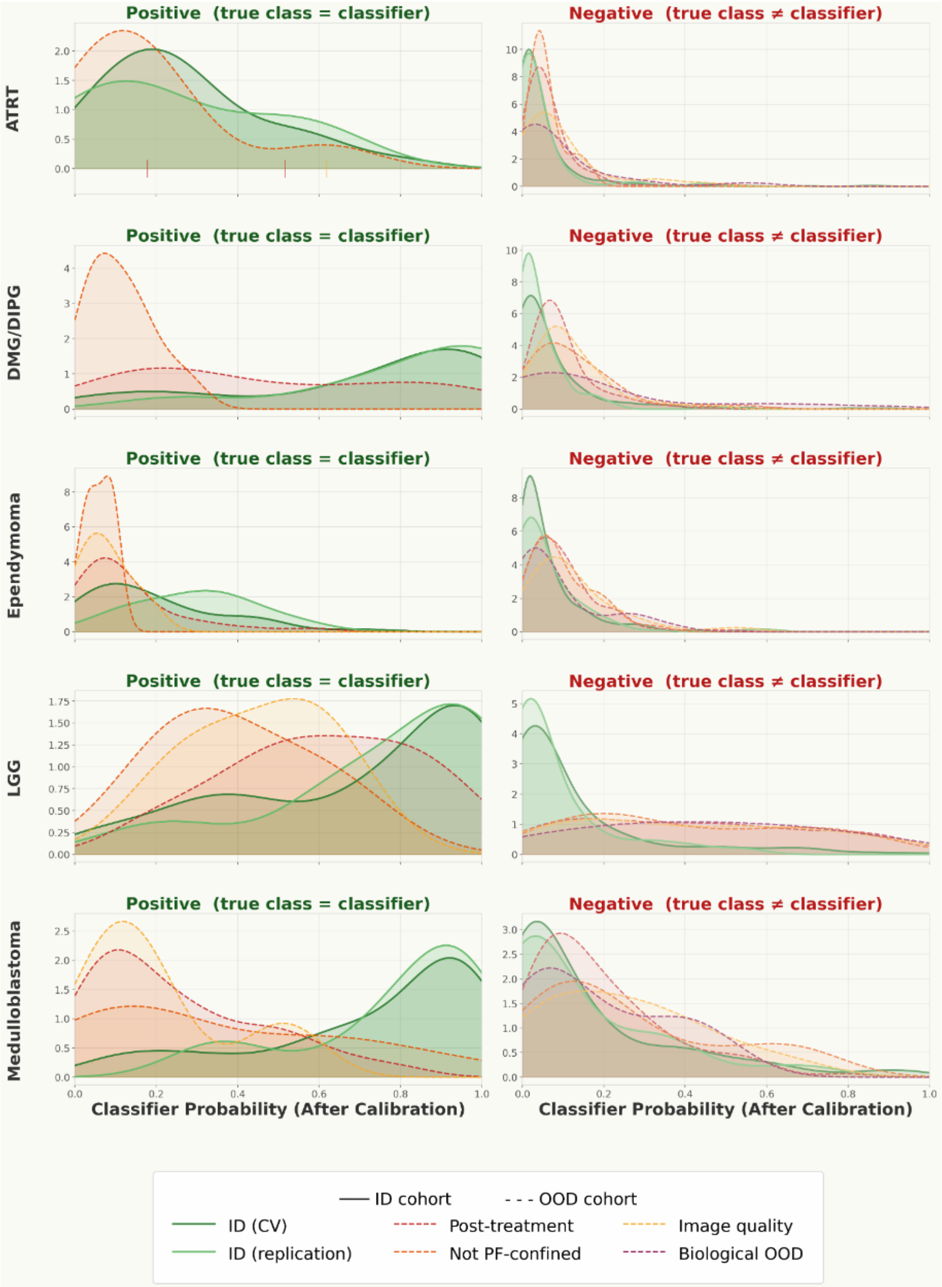


**Supplementary Fig. 5 | Kernel density estimates of calibrated per-class classifier probabilities.** Positive cases (left) correspond to subjects whose true histology matches the classifier; negative cases (right) correspond to all other histologies. Solid green curves denote in-distribution cohorts and dashed colored curves denote shifted cohorts.

**Supplementary Table 14 | AUC-ROC for separating each cohort from discovery,** all twenty scores, sorted by the mean across the four out-of-distribution strata. CHOA MB, CHOA medulloblastomas (n = 102).

| | In-distribution | | Out-of-distribution | | | | |
|---|---|---|---|---|---|---|---|
| **Score** | **External (CHOA MB)** | **ID (replication)** | **Biological OOD** | **Image quality** | **Not PF-confined** | **Post-treatment** | **Mean OOD** |
| **Energy** | 0.474 | 0.453 | 0.653 | 0.774 | 0.746 | 0.728 | 0.725 |
| **GEN** | 0.486 | 0.459 | 0.656 | 0.774 | 0.743 | 0.727 | 0.725 |
| **NNGuide** | 0.467 | 0.455 | 0.657 | 0.770 | 0.743 | 0.722 | 0.723 |
| **MaxLogit** | 0.474 | 0.458 | 0.666 | 0.764 | 0.733 | 0.724 | 0.722 |
| **ReAct + Energy** | 0.469 | 0.464 | 0.651 | 0.768 | 0.737 | 0.722 | 0.720 |
| **Entropy** | 0.490 | 0.465 | 0.669 | 0.759 | 0.724 | 0.714 | 0.717 |
| **MSP** | 0.490 | 0.470 | 0.687 | 0.732 | 0.703 | 0.698 | 0.705 |
| **kNN (256-d)** | 0.429 | 0.474 | 0.657 | 0.749 | 0.715 | 0.689 | 0.703 |
| **MSP + kNN (rank avg)** | 0.532 | 0.474 | 0.610 | 0.721 | 0.708 | 0.664 | 0.676 |
| **Crop argmax instability** | 0.410 | 0.474 | 0.716 | 0.683 | 0.641 | 0.660 | 0.675 |
| **Crop mutual information** | 0.433 | 0.459 | 0.716 | 0.651 | 0.654 | 0.660 | 0.670 |
| **ViM** | 0.450 | 0.477 | 0.613 | 0.705 | 0.644 | 0.634 | 0.649 |
| **Rel. Mahalanobis (1024-d)** | 0.517 | 0.517 | 0.571 | 0.672 | 0.666 | 0.616 | 0.631 |
| **Mahalanobis (1024-d)** | 0.501 | 0.505 | 0.500 | 0.707 | 0.610 | 0.599 | 0.604 |
| **Rel. Mahalanobis (256-d)** | 0.508 | 0.447 | 0.547 | 0.556 | 0.617 | 0.598 | 0.579 |
| **Crop dispersion** | 0.383 | 0.455 | 0.491 | 0.592 | 0.582 | 0.641 | 0.576 |
| **Crop cosine spread** | 0.382 | 0.455 | 0.491 | 0.591 | 0.582 | 0.641 | 0.576 |
| **kNN (1024-d)** | 0.563 | 0.495 | 0.484 | 0.621 | 0.617 | 0.557 | 0.570 |
| **Mahalanobis (256-d)** | 0.454 | 0.538 | 0.506 | 0.491 | 0.424 | 0.428 | 0.462 |
| **Crop energy s.d.** | 0.586 | 0.535 | 0.358 | 0.301 | 0.345 | 0.359 | 0.341 |

**Supplementary Table 15 | Number of scores separating each cohort from discovery at a Holm-adjusted two-sided *P* below 0.05.** Counts include scores significant in the reverse direction (AUC-ROC < 0.5): crop energy s.d. in the post-treatment, not PF-confined and image-quality strata, Mahalanobis (256-d) in the post-treatment stratum, and both significant scores for CHOA (crop dispersion and crop cosine spread). Counts in the expected direction are 18, 13 and 14 for the post-treatment, not PF-confined and image-quality strata, respectively. CHOA MB, CHOA medulloblastomas (n = 102).

| **Cohort** | **Scores tested** | **Detecting after Holm** | **Best AUC-ROC** |
|---|---|---|---|
| **Image quality** | 20 | 15 | 0.774 |
| **Not PF-confined** | 20 | 14 | 0.746 |
| **Post-treatment** | 20 | 20 | 0.728 |
| **Biological OOD** | 20 | 2 | 0.716 |
| **External (CHOA MB)** | 20 | 2 | 0.586 |
| **ID (replication)** | 20 | 0 | 0.538 |

**Supplementary Table 16 | Complete detection metrics against discovery.** AUC-ROC intervals are stratified bootstrap with 2,000 resamples; detection at ID95 is the fraction of the stratum above the 95th percentile of discovery. CHOA MB, CHOA medulloblastomas (n = 102).

| Cohort | Score | n | AUC-ROC | CI lo | CI hi | Holm P | AUPR-out | FPR@95TPR | det@ID95 |
|---|---|---|---|---|---|---|---|---|---|
| **ID (replication)** | Mahalanobis (256-d) | 114 | 0.538 | 0.480 | 0.597 | 0.868 | 0.221 | 0.913 | 0.061 |
| **ID (replication)** | Crop energy s.d. | 114 | 0.535 | 0.474 | 0.594 | 0.868 | 0.217 | 0.955 | 0.079 |
| **ID (replication)** | Rel. Mahalanobis (1024-d) | 114 | 0.517 | 0.468 | 0.572 | 0.868 | 0.190 | 0.864 | 0.053 |
| **ID (replication)** | Mahalanobis (1024-d) | 114 | 0.505 | 0.449 | 0.563 | 0.868 | 0.192 | 0.959 | 0.053 |
| **ID (replication)** | kNN (1024-d) | 114 | 0.495 | 0.438 | 0.551 | 0.868 | 0.182 | 0.904 | 0.018 |
| **ID (replication)** | ViM | 114 | 0.477 | 0.419 | 0.535 | 0.868 | 0.188 | 0.913 | 0.053 |
| **ID (replication)** | Crop argmax instability | 114 | 0.474 | 0.419 | 0.532 | 0.868 | 0.179 | 1.000 | 0.009 |
| **ID (replication)** | kNN (256-d) | 114 | 0.474 | 0.419 | 0.529 | 0.868 | 0.176 | 0.937 | 0.035 |
| **ID (replication)** | MSP + kNN (rank avg) | 114 | 0.474 | 0.416 | 0.534 | 0.868 | 0.179 | 0.917 | 0.026 |
| **ID (replication)** | MSP | 114 | 0.471 | 0.411 | 0.531 | 0.868 | 0.183 | 0.959 | 0.061 |
| **ID (replication)** | Entropy | 114 | 0.465 | 0.406 | 0.521 | 0.868 | 0.180 | 0.959 | 0.061 |
| **ID (replication)** | ReAct + Energy | 114 | 0.464 | 0.411 | 0.521 | 0.868 | 0.178 | 0.947 | 0.044 |
| **ID (replication)** | GEN | 114 | 0.459 | 0.400 | 0.518 | 0.868 | 0.177 | 0.951 | 0.053 |
| **ID (replication)** | Crop mutual information | 114 | 0.459 | 0.401 | 0.514 | 0.868 | 0.177 | 0.935 | 0.035 |
| **ID (replication)** | MaxLogit | 114 | 0.457 | 0.400 | 0.516 | 0.868 | 0.177 | 0.941 | 0.053 |
| **ID (replication)** | NNGuide | 114 | 0.455 | 0.399 | 0.513 | 0.868 | 0.173 | 0.943 | 0.035 |
| **ID (replication)** | Crop dispersion | 114 | 0.455 | 0.397 | 0.514 | 0.868 | 0.175 | 0.961 | 0.061 |
| **ID (replication)** | Crop cosine spread | 114 | 0.454 | 0.394 | 0.512 | 0.868 | 0.175 | 0.961 | 0.061 |
| **ID (replication)** | Energy | 114 | 0.453 | 0.396 | 0.511 | 0.868 | 0.174 | 0.941 | 0.044 |
| **ID (replication)** | Rel. Mahalanobis (256-d) | 114 | 0.447 | 0.397 | 0.496 | 0.868 | 0.157 | 0.860 | 0.000 |
| **External (CHOA MB)** | Crop energy s.d. | 102 | 0.586 | 0.531 | 0.640 | 0.109 | 0.213 | 0.900 | 0.049 |
| **External (CHOA MB)** | kNN (1024-d) | 102 | 0.563 | 0.500 | 0.623 | 0.613 | 0.225 | 0.931 | 0.118 |
| **External (CHOA MB)** | MSP + kNN (rank avg) | 102 | 0.532 | 0.471 | 0.595 | 0.967 | 0.191 | 0.917 | 0.049 |
| **External (CHOA MB)** | Rel. Mahalanobis (1024-d) | 102 | 0.517 | 0.463 | 0.569 | 0.967 | 0.165 | 0.856 | 0.010 |
| **External (CHOA MB)** | Rel. Mahalanobis (256-d) | 102 | 0.508 | 0.462 | 0.557 | 0.967 | 0.157 | 0.744 | 0.000 |
| **External (CHOA MB)** | Mahalanobis (1024-d) | 102 | 0.501 | 0.442 | 0.562 | 0.967 | 0.172 | 0.929 | 0.049 |
| **External (CHOA MB)** | MSP | 102 | 0.490 | 0.431 | 0.550 | 0.967 | 0.163 | 0.882 | 0.029 |
| **External (CHOA MB)** | Entropy | 102 | 0.490 | 0.433 | 0.544 | 0.967 | 0.162 | 0.878 | 0.010 |
| **External (CHOA MB)** | GEN | 102 | 0.486 | 0.429 | 0.546 | 0.967 | 0.163 | 0.911 | 0.010 |
| **External (CHOA MB)** | MaxLogit | 102 | 0.474 | 0.416 | 0.532 | 0.967 | 0.160 | 0.919 | 0.010 |
| **External (CHOA MB)** | Energy | 102 | 0.474 | 0.416 | 0.532 | 0.967 | 0.159 | 0.917 | 0.010 |
| **External (CHOA MB)** | ReAct + Energy | 102 | 0.469 | 0.411 | 0.527 | 0.967 | 0.158 | 0.911 | 0.010 |
| **External (CHOA MB)** | NNGuide | 102 | 0.467 | 0.409 | 0.525 | 0.967 | 0.156 | 0.913 | 0.020 |
| **External (CHOA MB)** | Mahalanobis (256-d) | 102 | 0.454 | 0.394 | 0.516 | 0.967 | 0.152 | 0.935 | 0.010 |
| **External (CHOA MB)** | ViM | 102 | 0.450 | 0.392 | 0.511 | 0.967 | 0.152 | 0.939 | 0.000 |
| **External (CHOA MB)** | Crop mutual information | 102 | 0.433 | 0.372 | 0.494 | 0.505 | 0.163 | 0.939 | 0.059 |
| **External (CHOA MB)** | kNN (256-d) | 102 | 0.429 | 0.373 | 0.491 | 0.371 | 0.144 | 0.972 | 0.010 |
| **External (CHOA MB)** | Crop argmax instability | 102 | 0.410 | 0.352 | 0.470 | 0.054 | 0.148 | 1.000 | 0.000 |
| **External (CHOA MB)** | Crop dispersion | 102 | 0.383 | 0.329 | 0.438 | 0.004 | 0.131 | 0.953 | 0.010 |
| **External (CHOA MB)** | Crop cosine spread | 102 | 0.382 | 0.325 | 0.438 | 0.004 | 0.131 | 0.955 | 0.010 |
| **Post-treatment** | Energy | 176 | 0.728 | 0.688 | 0.767 | 0.000 | 0.407 | 0.652 | 0.091 |
| **Post-treatment** | GEN | 176 | 0.727 | 0.686 | 0.766 | 0.000 | 0.416 | 0.661 | 0.102 |
| **Post-treatment** | MaxLogit | 176 | 0.723 | 0.683 | 0.761 | 0.000 | 0.410 | 0.654 | 0.102 |
| **Post-treatment** | NNGuide | 176 | 0.722 | 0.682 | 0.760 | 0.000 | 0.401 | 0.648 | 0.085 |
| **Post-treatment** | ReAct + Energy | 176 | 0.722 | 0.682 | 0.758 | 0.000 | 0.402 | 0.683 | 0.091 |

| Cohort | Score | n | AUC-ROC | CI lo | CI hi | Holm P | AUPR-out | FPR@95TPR | det@ID95 |
|---|---|---|---|---|---|---|---|---|---|
| **Post-treatment** | Entropy | 176 | 0.714 | 0.674 | 0.754 | 0.000 | 0.406 | 0.691 | 0.097 |
| **Post-treatment** | MSP | 176 | 0.698 | 0.655 | 0.737 | 0.000 | 0.393 | 0.699 | 0.114 |
| **Post-treatment** | kNN (256-d) | 176 | 0.689 | 0.648 | 0.731 | 0.000 | 0.367 | 0.654 | 0.068 |
| **Post-treatment** | MSP + kNN (rank avg) | 176 | 0.664 | 0.618 | 0.712 | 0.000 | 0.407 | 0.852 | 0.108 |
| **Post-treatment** | Crop argmax instability | 176 | 0.660 | 0.617 | 0.705 | 0.000 | 0.347 | 1.000 | 0.023 |
| **Post-treatment** | Crop mutual information | 176 | 0.660 | 0.618 | 0.701 | 0.000 | 0.329 | 0.652 | 0.023 |
| **Post-treatment** | Crop dispersion | 176 | 0.641 | 0.595 | 0.687 | 0.000 | 0.388 | 0.841 | 0.148 |
| **Post-treatment** | Crop cosine spread | 176 | 0.641 | 0.590 | 0.686 | 0.000 | 0.388 | 0.841 | 0.153 |
| **Post-treatment** | ViM | 176 | 0.635 | 0.593 | 0.675 | 0.000 | 0.307 | 0.667 | 0.023 |
| **Post-treatment** | Rel. Mahalanobis (1024-d) | 176 | 0.616 | 0.574 | 0.658 | 0.000 | 0.302 | 0.744 | 0.028 |
| **Post-treatment** | Mahalanobis (1024-d) | 176 | 0.599 | 0.557 | 0.644 | 0.000 | 0.306 | 0.823 | 0.034 |
| **Post-treatment** | Rel. Mahalanobis (256-d) | 176 | 0.598 | 0.554 | 0.644 | 0.000 | 0.288 | 0.768 | 0.006 |
| **Post-treatment** | kNN (1024-d) | 176 | 0.557 | 0.504 | 0.606 | 0.025 | 0.352 | 0.945 | 0.165 |
| **Post-treatment** | Mahalanobis (256-d) | 176 | 0.428 | 0.382 | 0.475 | 0.009 | 0.226 | 0.939 | 0.023 |
| **Post-treatment** | Crop energy s.d. | 176 | 0.359 | 0.314 | 0.406 | 0.000 | 0.199 | 0.951 | 0.028 |
| **Not PF-confined** | Energy | 39 | 0.746 | 0.679 | 0.809 | 0.000 | 0.159 | 0.553 | 0.154 |
| **Not PF-confined** | NNGuide | 39 | 0.743 | 0.674 | 0.805 | 0.000 | 0.167 | 0.535 | 0.154 |
| **Not PF-confined** | GEN | 39 | 0.743 | 0.674 | 0.803 | 0.000 | 0.163 | 0.555 | 0.154 |
| **Not PF-confined** | ReAct + Energy | 39 | 0.737 | 0.668 | 0.799 | 0.000 | 0.155 | 0.581 | 0.154 |
| **Not PF-confined** | MaxLogit | 39 | 0.733 | 0.666 | 0.793 | 0.000 | 0.153 | 0.553 | 0.128 |
| **Not PF-confined** | Entropy | 39 | 0.724 | 0.656 | 0.791 | 0.000 | 0.154 | 0.545 | 0.154 |
| **Not PF-confined** | kNN (256-d) | 39 | 0.715 | 0.646 | 0.779 | 0.000 | 0.158 | 0.563 | 0.128 |
| **Not PF-confined** | MSP + kNN (rank avg) | 39 | 0.708 | 0.630 | 0.782 | 0.000 | 0.186 | 0.720 | 0.128 |
| **Not PF-confined** | MSP | 39 | 0.703 | 0.639 | 0.765 | 0.000 | 0.135 | 0.553 | 0.128 |
| **Not PF-confined** | Rel. Mahalanobis (1024-d) | 39 | 0.666 | 0.594 | 0.736 | 0.006 | 0.122 | 0.756 | 0.103 |
| **Not PF-confined** | Crop mutual information | 39 | 0.654 | 0.574 | 0.728 | 0.012 | 0.114 | 0.793 | 0.103 |
| **Not PF-confined** | ViM | 39 | 0.645 | 0.583 | 0.704 | 0.019 | 0.097 | 0.530 | 0.077 |
| **Not PF-confined** | Crop argmax instability | 39 | 0.641 | 0.569 | 0.714 | 0.019 | 0.104 | 1.000 | 0.000 |
| **Not PF-confined** | Rel. Mahalanobis (256-d) | 39 | 0.617 | 0.550 | 0.681 | 0.076 | 0.090 | 0.626 | 0.000 |
| **Not PF-confined** | kNN (1024-d) | 39 | 0.617 | 0.518 | 0.716 | 0.076 | 0.175 | 0.904 | 0.205 |
| **Not PF-confined** | Mahalanobis (1024-d) | 39 | 0.610 | 0.534 | 0.689 | 0.088 | 0.097 | 0.724 | 0.051 |
| **Not PF-confined** | Crop cosine spread | 39 | 0.582 | 0.483 | 0.680 | 0.117 | 0.147 | 0.900 | 0.231 |
| **Not PF-confined** | Crop dispersion | 39 | 0.582 | 0.482 | 0.683 | 0.117 | 0.147 | 0.900 | 0.231 |
| **Not PF-confined** | Mahalanobis (256-d) | 39 | 0.425 | 0.339 | 0.512 | 0.117 | 0.064 | 0.907 | 0.077 |
| **Not PF-confined** | Crop energy s.d. | 39 | 0.345 | 0.264 | 0.429 | 0.012 | 0.055 | 0.939 | 0.026 |
| **Image quality** | GEN | 32 | 0.774 | 0.708 | 0.836 | 0.000 | 0.146 | 0.618 | 0.094 |
| **Image quality** | Energy | 32 | 0.774 | 0.707 | 0.837 | 0.000 | 0.137 | 0.600 | 0.125 |
| **Image quality** | NNGuide | 32 | 0.770 | 0.703 | 0.830 | 0.000 | 0.136 | 0.604 | 0.094 |
| **Image quality** | ReAct + Energy | 32 | 0.769 | 0.701 | 0.829 | 0.000 | 0.136 | 0.626 | 0.125 |
| **Image quality** | MaxLogit | 32 | 0.764 | 0.695 | 0.823 | 0.000 | 0.133 | 0.602 | 0.094 |
| **Image quality** | Entropy | 32 | 0.759 | 0.693 | 0.823 | 0.000 | 0.136 | 0.604 | 0.062 |
| **Image quality** | kNN (256-d) | 32 | 0.749 | 0.681 | 0.811 | 0.000 | 0.131 | 0.520 | 0.156 |
| **Image quality** | MSP | 32 | 0.732 | 0.666 | 0.792 | 0.000 | 0.120 | 0.587 | 0.125 |
| **Image quality** | MSP + kNN (rank avg) | 32 | 0.721 | 0.636 | 0.799 | 0.000 | 0.181 | 0.681 | 0.156 |
| **Image quality** | Mahalanobis (1024-d) | 32 | 0.707 | 0.628 | 0.782 | 0.001 | 0.154 | 0.632 | 0.125 |
| **Image quality** | ViM | 32 | 0.705 | 0.643 | 0.764 | 0.001 | 0.124 | 0.494 | 0.094 |
| **Image quality** | Crop argmax instability | 32 | 0.683 | 0.611 | 0.755 | 0.003 | 0.103 | 0.624 | 0.031 |

| Cohort | Score | n | AUC-ROC | CI lo | CI hi | Holm P | AUPR-out | FPR@95TPR | det@ID95 |
|---|---|---|---|---|---|---|---|---|---|
| **Image quality** | Rel. Mahalanobis (1024-d) | 32 | 0.672 | 0.599 | 0.743 | 0.008 | 0.109 | 0.724 | 0.094 |
| **Image quality** | Crop mutual information | 32 | 0.651 | 0.588 | 0.716 | 0.024 | 0.082 | 0.622 | 0.000 |
| **Image quality** | kNN (1024-d) | 32 | 0.621 | 0.509 | 0.730 | 0.111 | 0.126 | 0.837 | 0.219 |
| **Image quality** | Crop dispersion | 32 | 0.592 | 0.483 | 0.698 | 0.252 | 0.112 | 0.949 | 0.188 |
| **Image quality** | Crop cosine spread | 32 | 0.591 | 0.488 | 0.694 | 0.252 | 0.112 | 0.949 | 0.188 |
| **Image quality** | Rel. Mahalanobis (256-d) | 32 | 0.556 | 0.471 | 0.634 | 0.583 | 0.065 | 0.776 | 0.000 |
| **Image quality** | Mahalanobis (256-d) | 32 | 0.491 | 0.397 | 0.589 | 0.866 | 0.096 | 0.862 | 0.094 |
| **Image quality** | Crop energy s.d. | 32 | 0.301 | 0.215 | 0.384 | 0.001 | 0.041 | 0.986 | 0.000 |
| **Biological OOD** | Crop argmax instability | 22 | 0.716 | 0.644 | 0.789 | 0.009 | 0.073 | 0.624 | 0.000 |
| **Biological OOD** | Crop mutual information | 22 | 0.716 | 0.629 | 0.800 | 0.012 | 0.082 | 0.596 | 0.091 |
| **Biological OOD** | MSP | 22 | 0.687 | 0.586 | 0.774 | 0.054 | 0.075 | 0.644 | 0.045 |
| **Biological OOD** | Entropy | 22 | 0.669 | 0.566 | 0.757 | 0.122 | 0.066 | 0.657 | 0.045 |
| **Biological OOD** | MaxLogit | 22 | 0.666 | 0.569 | 0.754 | 0.133 | 0.066 | 0.709 | 0.045 |
| **Biological OOD** | kNN (256-d) | 22 | 0.658 | 0.565 | 0.746 | 0.171 | 0.064 | 0.776 | 0.000 |
| **Biological OOD** | NNGuide | 22 | 0.658 | 0.564 | 0.749 | 0.171 | 0.063 | 0.685 | 0.045 |
| **Biological OOD** | GEN | 22 | 0.656 | 0.562 | 0.746 | 0.171 | 0.063 | 0.695 | 0.000 |
| **Biological OOD** | Energy | 22 | 0.653 | 0.563 | 0.742 | 0.179 | 0.062 | 0.711 | 0.000 |
| **Biological OOD** | ReAct + Energy | 22 | 0.651 | 0.552 | 0.741 | 0.179 | 0.062 | 0.677 | 0.000 |
| **Biological OOD** | ViM | 22 | 0.613 | 0.515 | 0.707 | 0.637 | 0.082 | 0.667 | 0.136 |
| **Biological OOD** | MSP + kNN (rank avg) | 22 | 0.610 | 0.494 | 0.724 | 0.637 | 0.082 | 0.850 | 0.045 |
| **Biological OOD** | Rel. Mahalanobis (1024-d) | 22 | 0.571 | 0.456 | 0.681 | 0.997 | 0.088 | 0.805 | 0.136 |
| **Biological OOD** | Rel. Mahalanobis (256-d) | 22 | 0.547 | 0.442 | 0.648 | 0.997 | 0.047 | 0.793 | 0.000 |
| **Biological OOD** | Mahalanobis (256-d) | 22 | 0.506 | 0.386 | 0.626 | 0.997 | 0.082 | 0.825 | 0.136 |
| **Biological OOD** | Mahalanobis (1024-d) | 22 | 0.500 | 0.389 | 0.622 | 0.997 | 0.071 | 0.774 | 0.136 |
| **Biological OOD** | Crop dispersion | 22 | 0.491 | 0.353 | 0.628 | 0.997 | 0.126 | 0.972 | 0.136 |
| **Biological OOD** | Crop cosine spread | 22 | 0.491 | 0.342 | 0.641 | 0.997 | 0.126 | 0.972 | 0.136 |
| **Biological OOD** | kNN (1024-d) | 22 | 0.484 | 0.354 | 0.617 | 0.997 | 0.054 | 0.943 | 0.045 |
| **Biological OOD** | Crop energy s.d. | 22 | 0.358 | 0.250 | 0.464 | 0.238 | 0.036 | 0.884 | 0.045 |

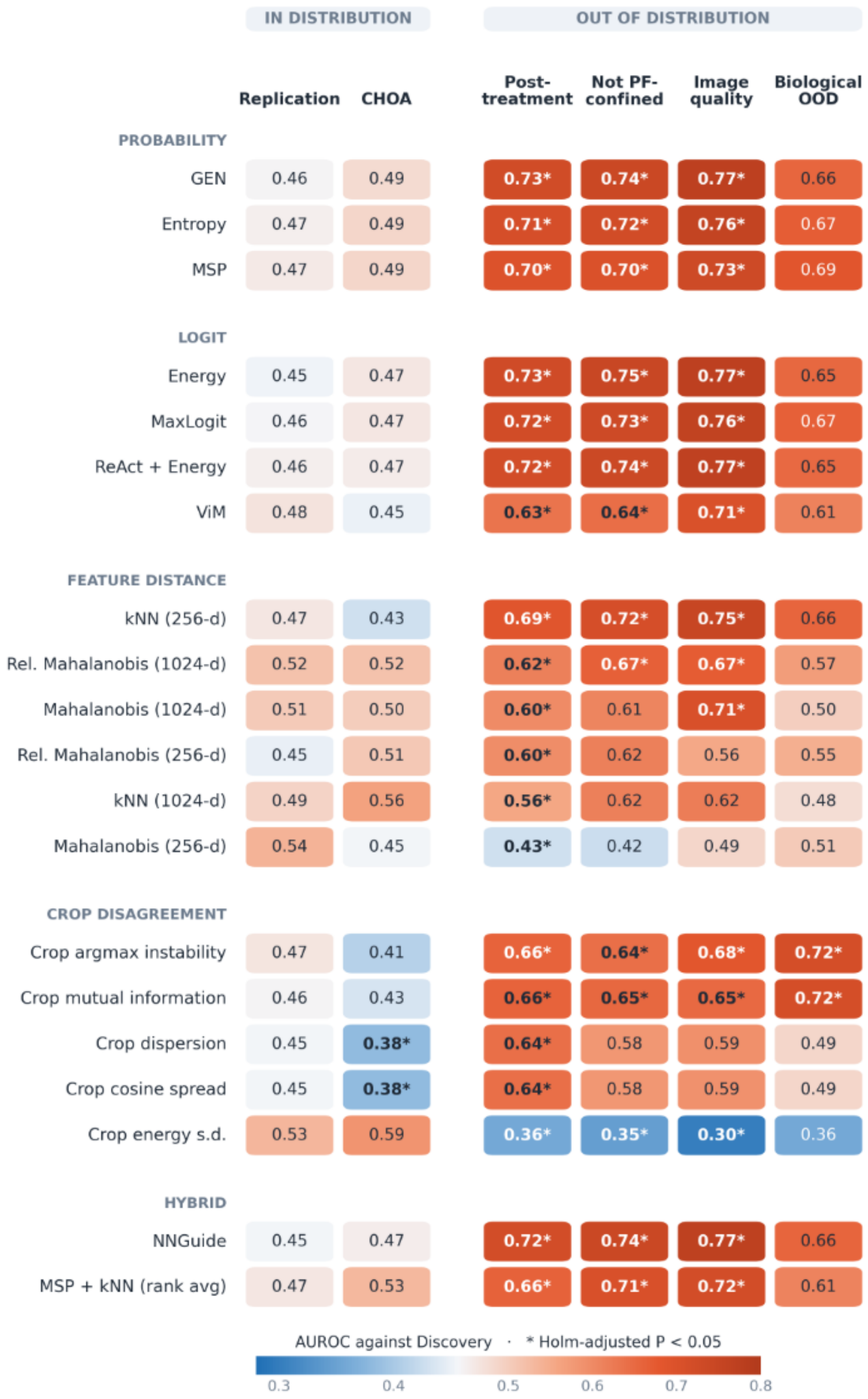


**Supplementary Fig. 6 | Detection AUC-ROC against discovery for all twenty post-hoc scores.** Rows are scores grouped by family (probability, logit, feature distance, crop disagreement, hybrid) and ordered within family by mean AUC-ROC across the out-of-distribution strata; columns are the two in-distribution cohorts (replication, CHOA) and the four out-of-distribution strata. Each cell is the AUC-ROC for separating the cohort from the out-of-fold discovery reference (n = 492); 0.5 is chance and values below 0.5 indicate separation in the reverse direction. *Holm-adjusted P < 0.05 within cohort.

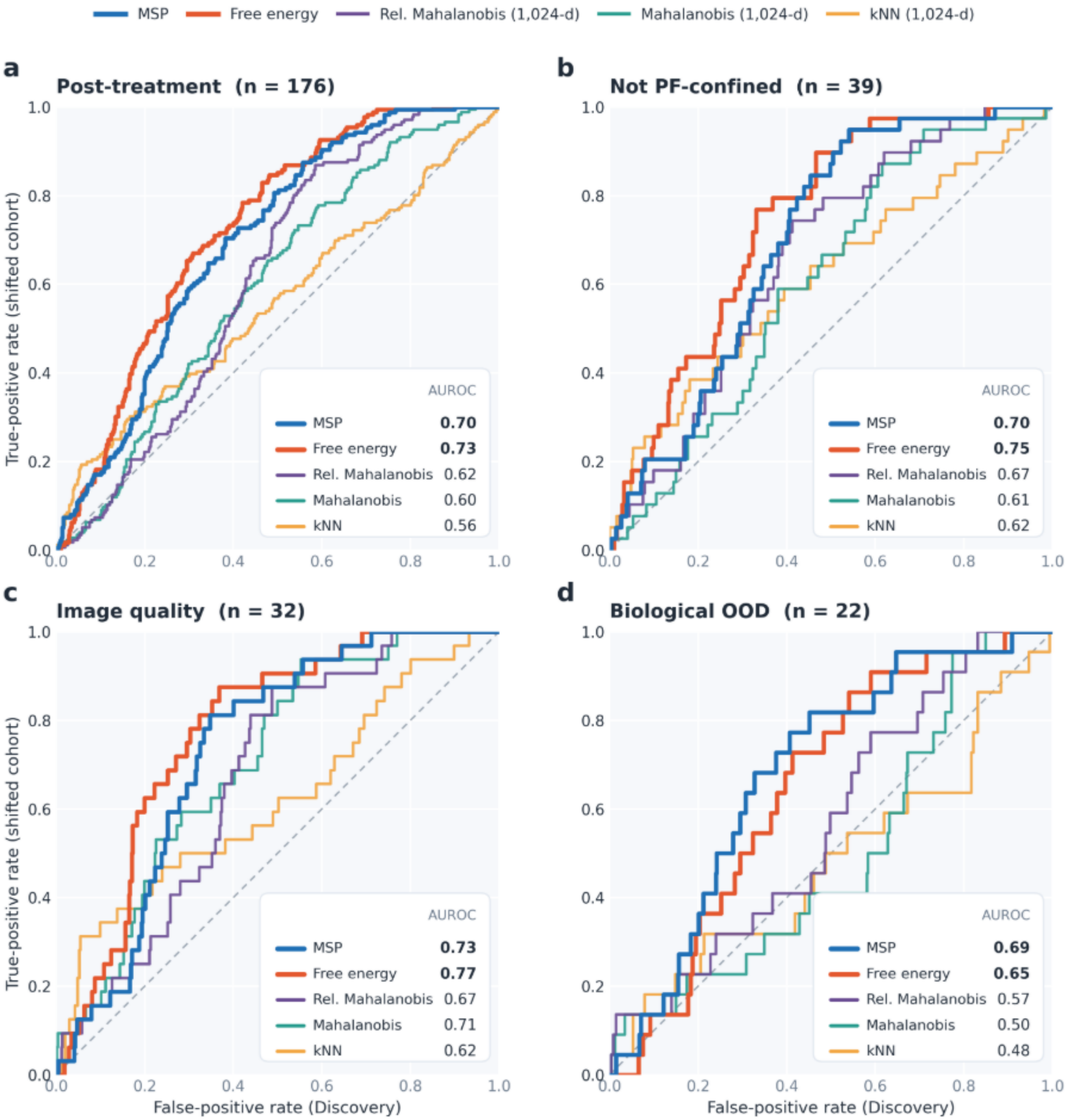


**Supplementary Fig. 7 | Receiver operating characteristic curves contrasting softmax and logit scores with feature-distance scores.** **a**, Post-treatment (n = 176). **b**, Not confined to the posterior fossa (n = 39). **c**, Inadequate image quality (n = 32). **d**, Biological OOD (n = 22). Thick lines show the maximum softmax probability (MSP) and free energy; thin lines show feature-distance scores computed in the frozen 1,024-dimensional representation (Mahalanobis, relative Mahalanobis and k-nearest-neighbour distance). The reference is the out-of-fold discovery cohort (n = 492); AUC-ROC values are given in each panel, and the dashed line marks chance. Distance scores in the 256-dimensional penultimate layer are shown in Supplementary Fig. 6.

**Supplementary Table 17 | Scores exceeding the maximum softmax probability after Holm adjustment, subject-level paired bootstrap of 2,000 resamples.** Candidate and baseline scores were resampled together (paired comparison).

| Stratum | Score | Δ vs MSP | CI lo | CI hi | Holm P |
|---|---|---|---|---|---|
| **Image quality** | GEN | 0.0415 | 0.0207 | 0.0645 | 0.0075 |
| **Image quality** | MaxLogit | 0.0318 | 0.0141 | 0.0511 | 0.0140 |
| **Image quality** | Entropy | 0.0271 | 0.0124 | 0.0422 | 0.0075 |
| **Not PF-confined** | Energy | 0.0434 | 0.0222 | 0.0658 | 0.0065 |
| **Not PF-confined** | NNGuide | 0.0406 | 0.0230 | 0.0589 | 0.0065 |
| **Not PF-confined** | GEN | 0.0400 | 0.0236 | 0.0575 | 0.0065 |
| **Not PF-confined** | ReAct + Energy | 0.0340 | 0.0126 | 0.0561 | 0.0220 |
| **Not PF-confined** | MaxLogit | 0.0298 | 0.0140 | 0.0458 | 0.0065 |
| **Not PF-confined** | Entropy | 0.0216 | 0.0110 | 0.0328 | 0.0065 |
| **Post-treatment** | Energy | 0.0300 | 0.0140 | 0.0460 | 0.0040 |
| **Post-treatment** | GEN | 0.0285 | 0.0153 | 0.0411 | 0.0040 |
| **Post-treatment** | MaxLogit | 0.0254 | 0.0146 | 0.0365 | 0.0040 |
| **Post-treatment** | NNGuide | 0.0243 | 0.0100 | 0.0385 | 0.0040 |
| **Post-treatment** | ReAct + Energy | 0.0236 | 0.0090 | 0.0383 | 0.0120 |
| **Post-treatment** | Entropy | 0.0162 | 0.0077 | 0.0250 | 0.0040 |

The strata differ in histological composition, and the CHOA comparison includes only medulloblastomas, whereas 39% of the discovery cohort is medulloblastoma. Any pooled comparison therefore confounds distribution shift with class mix. Detection was repeated with histology held fixed, comparing subjects of one class against subjects of the same class in discovery.

**Supplementary Table 18 | Class-matched detection using free energy.** Cells with fewer than five stratum subjects are omitted; cells with five to ten subjects are shown but are imprecise. ATRT is not shown for the post-treatment stratum (n = 2). CHOA MB, CHOA medulloblastomas (n = 102).

| Stratum | Class | n ref | n stratum | AUC-ROC | CI lo | CI hi | det@ID95 |
|---|---|---|---|---|---|---|---|
| **External (CHOA MB)** | Medulloblastoma | 190 | 102 | 0.521 | 0.452 | 0.591 | 0.059 |
| **Image quality** | Ependymoma | 37 | 5 | 0.562 | 0.284 | 0.841 | 0.200 |
| **Image quality** | Medulloblastoma | 190 | 22 | 0.817 | 0.707 | 0.928 | 0.182 |
| **Not PF-confined** | ATRT | 25 | 8 | 0.430 | 0.205 | 0.655 | 0.000 |
| **Not PF-confined** | DMG/DIPG | 82 | 9 | 0.787 | 0.605 | 0.970 | 0.333 |
| **Not PF-confined** | Ependymoma | 37 | 7 | 0.641 | 0.403 | 0.879 | 0.143 |
| **Not PF-confined** | Medulloblastoma | 190 | 12 | 0.739 | 0.574 | 0.903 | 0.167 |
| **Post-treatment** | DMG/DIPG | 82 | 22 | 0.746 | 0.619 | 0.873 | 0.136 |
| **Post-treatment** | Ependymoma | 37 | 35 | 0.605 | 0.474 | 0.736 | 0.200 |
| **Post-treatment** | LGG | 158 | 24 | 0.702 | 0.579 | 0.825 | 0.042 |
| **Post-treatment** | Medulloblastoma | 190 | 93 | 0.787 | 0.727 | 0.848 | 0.150 |

**Supplementary Table 19 | Pooled against class-matched detection.** The composition effect is pooled minus class-matched, re-weighted by the stratum's own class frequencies. CHOA MB, CHOA medulloblastomas (n = 102).

| Stratum | Score | Pooled | Class-matched | Composition effect | Classes used |
|---|---|---|---|---|---|
| **External (CHOA MB)** | Crop mutual information | 0.433 | 0.496 | -0.063 | 1 |
| **External (CHOA MB)** | Energy | 0.474 | 0.521 | -0.047 | 1 |

| Stratum | Score | Pooled | Class-matched | Composition effect | Classes used |
|---|---|---|---|---|---|
| **External (CHOA MB)** | Entropy | 0.490 | 0.526 | -0.036 | 1 |
| **External (CHOA MB)** | MSP | 0.490 | 0.526 | -0.035 | 1 |
| **External (CHOA MB)** | Rel. Mahalanobis (256-d) | 0.508 | 0.523 | -0.015 | 1 |
| **External (CHOA MB)** | kNN (256-d) | 0.429 | 0.477 | -0.048 | 1 |
| **ID (replication)** | Crop mutual information | 0.459 | 0.465 | -0.006 | 5 |
| **ID (replication)** | Energy | 0.453 | 0.453 | -0.000 | 5 |
| **ID (replication)** | Entropy | 0.465 | 0.466 | -0.001 | 5 |
| **ID (replication)** | MSP | 0.470 | 0.475 | -0.004 | 5 |
| **ID (replication)** | Rel. Mahalanobis (256-d) | 0.447 | 0.444 | 0.003 | 5 |
| **ID (replication)** | kNN (256-d) | 0.474 | 0.472 | 0.002 | 5 |
| **Image quality** | Crop mutual information | 0.651 | 0.728 | -0.076 | 2 |
| **Image quality** | Energy | 0.774 | 0.770 | 0.004 | 2 |
| **Image quality** | Entropy | 0.759 | 0.754 | 0.005 | 2 |
| **Image quality** | MSP | 0.732 | 0.733 | -0.000 | 2 |
| **Image quality** | Rel. Mahalanobis (256-d) | 0.556 | 0.541 | 0.014 | 2 |
| **Image quality** | kNN (256-d) | 0.749 | 0.741 | 0.008 | 2 |
| **Not PF-confined** | Crop mutual information | 0.654 | 0.600 | 0.054 | 4 |
| **Not PF-confined** | Energy | 0.746 | 0.663 | 0.083 | 4 |
| **Not PF-confined** | Entropy | 0.724 | 0.633 | 0.091 | 4 |
| **Not PF-confined** | MSP | 0.703 | 0.608 | 0.095 | 4 |
| **Not PF-confined** | Rel. Mahalanobis (256-d) | 0.617 | 0.596 | 0.021 | 4 |
| **Not PF-confined** | kNN (256-d) | 0.715 | 0.617 | 0.098 | 4 |
| **Post-treatment** | Crop mutual information | 0.660 | 0.690 | -0.030 | 4 |
| **Post-treatment** | Energy | 0.728 | 0.734 | -0.006 | 4 |
| **Post-treatment** | Entropy | 0.714 | 0.711 | 0.003 | 4 |
| **Post-treatment** | MSP | 0.698 | 0.695 | 0.003 | 4 |
| **Post-treatment** | Rel. Mahalanobis (256-d) | 0.598 | 0.599 | -0.001 | 4 |
| **Post-treatment** | kNN (256-d) | 0.689 | 0.687 | 0.002 | 4 |

The biological stratum carries no histology inside the label space, so no class matches and it was compared against each discovery class in turn.

**Supplementary Table 20 | Biological out-of-distribution subjects against each discovery class, free energy.**

| Versus discovery class | n ref | n stratum | AUC-ROC | CI lo | CI hi | det@ID95 |
|---|---|---|---|---|---|---|
| **Medulloblastoma** | 190 | 22 | 0.717 | 0.592 | 0.843 | 0.091 |
| **DMG/DIPG** | 82 | 22 | 0.687 | 0.553 | 0.820 | 0.000 |
| **LGG** | 158 | 22 | 0.670 | 0.540 | 0.800 | 0.000 |
| **Ependymoma** | 37 | 22 | 0.429 | 0.279 | 0.579 | 0.000 |
| **ATRT** | 25 | 22 | 0.282 | 0.136 | 0.428 | 0.000 |

The stratum separates from medulloblastoma, diffuse midline glioma and low-grade glioma, does not separate from ependymoma, and separates from atypical teratoid rhabdoid tumor in the opposite direction with an interval excluding chance. The model was therefore less confident on in-distribution

ATRT than on tumors with unseen histologies, so confidence-based abstention could not distinguish them.

**Supplementary Table 21 | Post-hoc scores separating the biological OOD stratum from discovery after Holm adjustment. Both scores measure disagreement among the 18 crops.**

| Score | AUC-ROC | CI lo | CI hi | Holm P | det@ID95 | det lo | det hi |
|---|---|---|---|---|---|---|---|
| **Crop mutual information** | 0.716 | 0.629 | 0.800 | 0.012 | 0.091 | 0.000 | 0.273 |
| **Crop argmax instability** | 0.716 | 0.644 | 0.789 | 0.009 | 0.000 | 0.000 | 0.000 |

#### Supplementary Note 7 | Geometry of the frozen representation

**Methods. T**he analysis is described in the main text (Methods). Covariances used Ledoit–Wolf shrinkage (ref. S13), and class separability was measured by the silhouette coefficient (ref. S15) and by k-nearest-neighbour class purity (k = 22).

**Results.** Feature-distance OOD scores did not outperform the maximum softmax probability on any stratum (Supplementary Tables 14 and 17). The analyses below examine why. In the standardized frozen embedding (z, 1,024 dimensions) the five histologies show essentially no cluster structure (Supplementary Table 22), yet a linear probe on the same features reaches a macro AUC-ROC of 0.887 (Supplementary Table 25): class information is spread across many individually weak directions, which a hyperplane can weight and combine but a distance metric, weighting every direction by its variance, cannot. ATRT and ependymoma, the two classes that fail downstream, have the most negative silhouettes, meaning their subjects are on average closer to other classes than to their own.

The external medulloblastoma cohort lands nearest the medulloblastoma centroid in 81.4% of cases, against a discovery base rate of 39.2% (Supplementary Table 23). No cohort lies outside the in-distribution cloud: acquisition shift is partly visible (cases of inadequate image quality sit at the 79th percentile of the discovery distance distribution), whereas biological shift is not (biological OOD cases sit at the 40th percentile, closer to the center than a typical training subject; Supplementary Table 24). Projecting onto the four out-of-fold Fisher discriminant directions, which span 1.0% of the total variance, lowered the silhouette from 0.008 to −0.020 (Supplementary Table 25). Tumors with unseen histologies were therefore not distinguishable from in-distribution tumors in the frozen feature space and were detected, if at all, only through disagreement among the 18 crop-wise predictions.

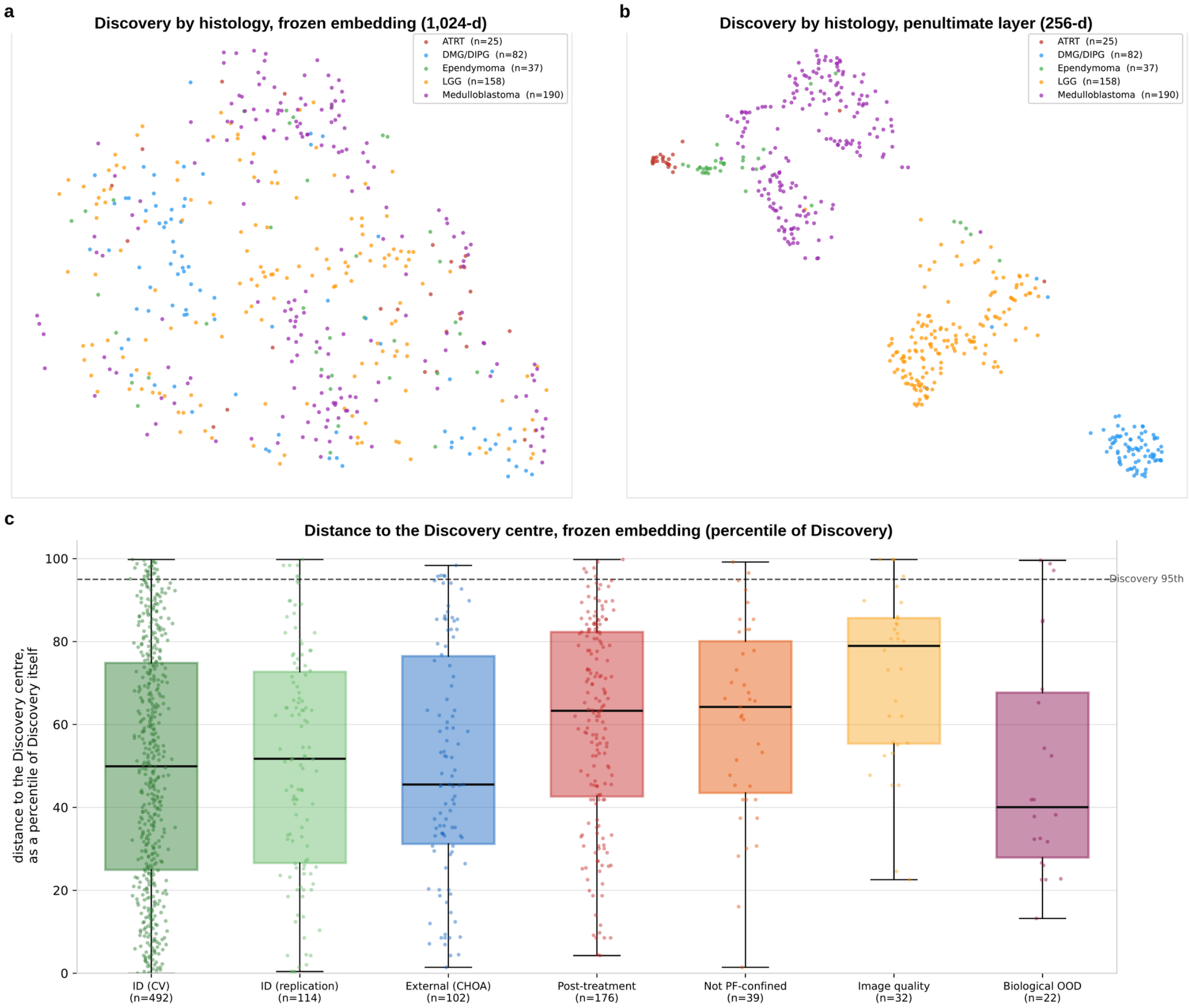


**Supplementary Fig. 8 | Geometry of the frozen and penultimate representations. a**, **b**, UMAP (ref. S14) projections of discovery subjects colored by histology in the frozen 1,024-dimensional embedding (**a**) and the 256-dimensional penultimate layer of the head (**b**); UMAP is used for display only. **c**, Mahalanobis distance of each subject to the discovery centre in the frozen embedding, expressed as a percentile of discovery's own distance distribution; the dashed line marks the discovery 95th percentile.

**Supplementary Table 22 | Class separability on discovery (n = 492).** Silhouette coefficient and k-nearest-neighbor class purity (k = 22; chance purity 0.288). Penultimate-space values use the deployed head on its own training subjects and are therefore optimistic (out of fold: silhouette 0.074, purity 0.680).

| Space | Class | n | Silhouette | KNN purity |
|---|---|---|---|---|
| **Frozen (z, 1,024-d)** | All classes | 492 | 0.008 | 0.495 |
| | ATRT | 25 | −0.038 | 0.189 |
| | DMG/DIPG | 82 | 0.031 | 0.439 |
| | Ependymoma | 37 | −0.020 | 0.138 |
| | LGG | 158 | −0.006 | 0.505 |
| | Medulloblastoma | 190 | 0.021 | 0.621 |
| **Penultimate (h, 256-d)** | All classes | 492 | 0.254 | 0.930 |
| | ATRT | 25 | 0.240 | 0.740 |
| | DMG/DIPG | 82 | 0.365 | 0.950 |

| Space | Class | n | Silhouette | KNN purity |
|---|---|---|---|---|
| | Ependymoma | 37 | 0.203 | 0.576 |
| | LGG | 158 | 0.302 | 0.976 |
| | Medulloblastoma | 190 | 0.178 | 0.977 |

**Supplementary Table 23 | Nearest discovery class centroid in the frozen space, by cohort.** Values are the proportion of subjects in each cohort whose nearest class centroid is the named class. Discovery assignments use centroids fitted on the other four folds. CHOA MB, CHOA medulloblastomas (n = 102).

| Cohort | n | ATRT | DMG/DIPG | Ependymoma | LGG | Medulloblastoma |
|---|---|---|---|---|---|---|
| **Discovery** | 492 | 0.022 | 0.161 | 0.047 | 0.378 | 0.392 |
| **Replication** | 114 | 0.009 | 0.158 | 0.070 | 0.263 | 0.500 |
| **External (CHOA MB)** | 102 | 0.000 | 0.010 | 0.059 | 0.118 | 0.814 |
| **Post-treatment** | 176 | 0.011 | 0.074 | 0.114 | 0.460 | 0.341 |
| **Not PF-confined** | 39 | 0.000 | 0.154 | 0.128 | 0.359 | 0.359 |
| **Image quality** | 32 | 0.000 | 0.062 | 0.156 | 0.469 | 0.312 |
| **Biological OOD** | 22 | 0.000 | 0.273 | 0.091 | 0.545 | 0.091 |

**Supplementary Table 24 | Mahalanobis distance to the discovery centre in the frozen space.** Distance is expressed as a percentile of discovery's own distance distribution (50 = a typical training subject); beyond ID95 is the fraction of the cohort beyond the 95th percentile of discovery. CHOA MB, CHOA medulloblastomas (n = 102).

| Cohort | n | Median percentile of discovery | Fraction beyond ID95 |
|---|---|---|---|
| **Discovery (out of fold)** | 492 | 49.9 | 0.049 |
| **Replication** | 114 | 51.7 | 0.053 |
| **External (CHOA MB)** | 102 | 45.5 | 0.049 |
| **Post-treatment** | 176 | 63.3 | 0.034 |
| **Not PF-confined** | 39 | 64.2 | 0.051 |
| **Image quality** | 32 | 79.0 | 0.125 |
| **Biological OOD** | 22 | 40.0 | 0.136 |

**Supplementary Table 25 | Fold-respecting linear probe on the top k principal components of the frozen embedding.** Components ranked by explained variance or by class-discriminative power (F statistic) within each training fold.

| K | Macro AUC, ranked by variance | Macro AUC, ranked by class power |
|---|---|---|
| 2 | 0.604 | 0.723 |
| 4 | 0.719 | 0.793 |
| 8 | 0.840 | 0.843 |
| 16 | 0.867 | 0.866 |
| 32 | 0.869 | 0.858 |
| 64 | 0.880 | 0.875 |
| 120 | 0.880 | 0.880 |
| 1,024 (all) | 0.887 | 0.887 |

#### Supplementary Note 8 | Robustness to simulated imaging perturbations

**Methods.** The perturbation protocol is described in the main text (Methods). Ghosting and motion were simulated in k-space with TorchIO (version 0.21.3; ref. 21 of the main text) on the unprocessed images, which were then preprocessed with the containerized pipeline (registration to the SRI24 atlas [ref. 32 of the main text], skull stripping and intensity normalization). A random seed was derived for each subject and condition, so that every perturbed volume can be reproduced.

Rotations were applied after preprocessing because registration to SRI24 restores the orientation of a rotated input; rotation before preprocessing would therefore mainly test the registration step. Severity trends were tested with logistic generalized estimating equations (ref. S23); outcomes and tests are described in the main text (Methods, Statistical analysis).

**Results.** Per-condition results for the 42 perturbed conditions and the unperturbed reference are given in Supplementary Table 26, and the severity trends and the response of abstention to errors introduced by perturbation in Supplementary Table 27. Rotation results by axis and angle are shown in Supplementary Fig. 10. The perturbations are illustrated on one subject in Supplementary Fig. 9.

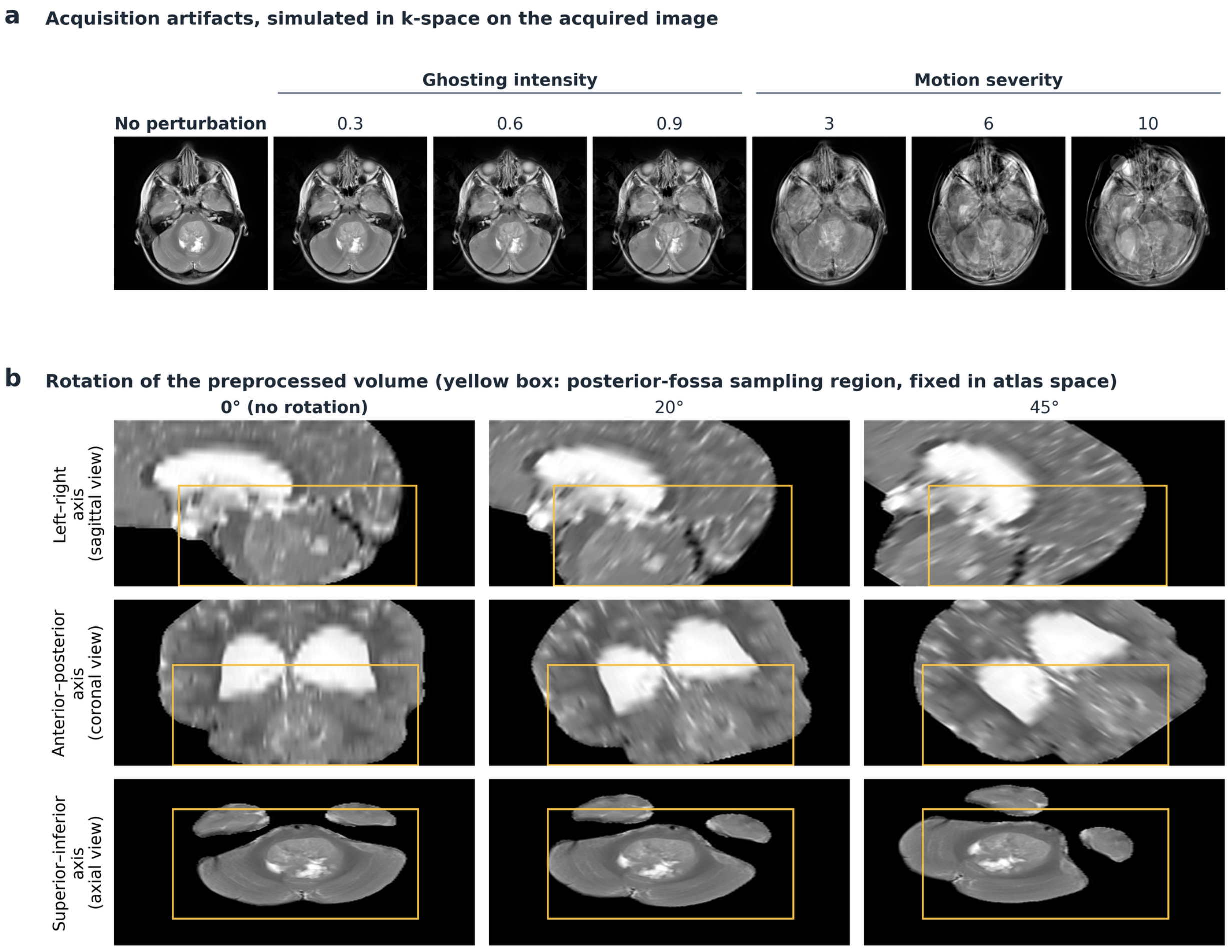


**Supplementary Fig. 9 | The simulated perturbations, shown on one replication subject.** a, Ghosting (intensity 0.3, 0.6 and 0.9) and motion (severity 3, 6 and 10); each image is shown after registration to the SRI24 atlas, at the same axial slice through the tumor as the unperturbed image. b, Rigid rotation of the preprocessed volume by 20° and 45° about the left-right, anterior-posterior, and superior-inferior axes, shown in sagittal, coronal and axial views. The yellow box marks the posterior-fossa region from which the classifier samples; it is fixed in atlas coordinates. Rotations of ±5° to ±45° in both directions were evaluated (Supplementary Table 26).

**Supplementary Table 26 | Classifier performance under each simulated perturbation (n = 50).** Values in brackets are subject-level bootstrap 95% confidence intervals (2,000 resamples). Class changed, proportion of subjects whose predicted class differed from the unperturbed prediction; TV, median total-variation distance between perturbed and unperturbed calibrated probabilities; C→I and

I→C, subjects changing from correct to incorrect and from incorrect to correct; P, exact two-sided McNemar test against the unperturbed reference, Holm-adjusted within each family (ghosting, motion and each rotation axis).

| Condition | Accuracy without abstention, % (95% CI) | Abstained, % (95% CI) | Accuracy on predicted, % (95% CI) | Class changed, % | TV, median | C→I / I→C | P (Holm) |
|---|---|---|---|---|---|---|---|
| No perturbation | 80.0 (68.0–90.0) | 18.0 (8.0–28.0) | 95.1 (87.2–100.0) | 0.0 | 0.000 | 0/0 | — |
| **Ghosting** | | | | | | | |
| Intensity 0.3 | 76.0 (64.0–88.0) | 26.0 (14.0–38.0) | 91.9 (81.4–100.0) | 4.0 | 0.045 | 2/0 | 0.50 |
| Intensity 0.6 | 70.0 (58.0–82.0) | 20.0 (10.0–32.0) | 82.5 (69.8–93.0) | 14.0 | 0.087 | 6/1 | 0.25 |
| Intensity 0.9 | 68.0 (54.0–80.0) | 24.0 (12.0–36.0) | 81.6 (68.4–92.9) | 14.0 | 0.104 | 6/0 | 0.094 |
| **Motion** | | | | | | | |
| Severity 3 | 68.0 (54.0–80.0) | 24.0 (12.0–36.0) | 86.8 (75.0–97.2) | 16.0 | 0.075 | 6/0 | 0.043 |
| Severity 6 | 64.0 (50.0–78.0) | 32.0 (20.0–46.0) | 76.5 (61.8–90.6) | 22.0 | 0.117 | 8/0 | 0.023 |
| Severity 10 | 64.0 (50.0–78.0) | 36.0 (22.0–50.0) | 81.2 (66.7–94.1) | 22.0 | 0.147 | 9/1 | 0.043 |
| **Rotation, left–right axis** | | | | | | | |
| -45° | 44.0 (30.0–58.0) | 48.0 (34.0–62.0) | 50.0 (30.3–69.2) | 54.0 | 0.479 | 20/2 | 0.002 |
| -30° | 58.0 (44.0–72.0) | 40.0 (26.0–54.0) | 73.3 (57.6–88.9) | 38.0 | 0.321 | 14/3 | 0.13 |
| -20° | 74.0 (62.0–86.0) | 34.0 (22.0–48.0) | 84.9 (71.0–95.0) | 18.0 | 0.197 | 5/2 | 1.00 |
| -15° | 80.0 (68.0–90.0) | 30.0 (18.0–44.0) | 85.7 (73.0–95.3) | 12.0 | 0.128 | 2/2 | 1.00 |
| -10° | 82.0 (70.0–92.0) | 20.0 (10.0–32.0) | 87.5 (76.3–95.8) | 8.0 | 0.069 | 1/2 | 1.00 |
| -5° | 82.0 (70.0–92.0) | 16.0 (6.0–28.0) | 92.9 (84.4–100.0) | 4.0 | 0.045 | 0/1 | 1.00 |
| +5° | 86.0 (76.0–94.0) | 16.0 (6.0–28.0) | 92.9 (84.6–100.0) | 6.0 | 0.059 | 0/3 | 1.00 |
| +10° | 82.0 (70.0–92.0) | 14.0 (4.0–24.0) | 88.4 (78.0–97.6) | 12.0 | 0.084 | 2/3 | 1.00 |
| +15° | 78.0 (66.0–90.0) | 20.0 (10.0–32.0) | 87.5 (76.3–97.4) | 18.0 | 0.121 | 4/3 | 1.00 |
| +20° | 76.0 (64.0–88.0) | 22.0 (10.0–34.0) | 87.2 (76.3–97.4) | 22.0 | 0.155 | 5/3 | 1.00 |
| +30° | 68.0 (56.0–80.0) | 26.0 (14.0–38.0) | 75.7 (61.1–89.2) | 26.0 | 0.208 | 9/3 | 1.00 |
| +45° | 50.0 (36.0–64.0) | 30.0 (18.0–42.0) | 60.0 (43.3–76.9) | 44.0 | 0.363 | 16/1 | 0.003 |
| **Rotation, anterior–** | | | | | | | |

| | | | | | | | |
|---|---|---|---|---|---|---|---|
| **posterior axis** | | | | | | | |
| -45° | 58.0 (44.0–72.0) | 50.0 (36.0–64.0) | 76.0 (59.3–91.7) | 34.0 | 0.279 | 13/2 | 0.081 |
| -30° | 78.0 (66.0–90.0) | 26.0 (14.0–40.0) | 89.2 (78.1–97.6) | 16.0 | 0.164 | 3/2 | 1.00 |
| -20° | 80.0 (68.0–90.0) | 16.0 (6.0–26.0) | 88.1 (77.5–97.5) | 12.0 | 0.076 | 2/2 | 1.00 |
| -15° | 80.0 (68.0–90.0) | 16.0 (6.0–26.1) | 88.1 (77.3–97.6) | 10.0 | 0.057 | 2/2 | 1.00 |
| -10° | 82.0 (72.0–92.0) | 12.0 (4.0–22.0) | 86.4 (75.6–95.5) | 8.0 | 0.045 | 1/2 | 1.00 |
| -5° | 84.0 (74.0–94.0) | 14.0 (4.0–24.0) | 90.7 (81.0–97.8) | 4.0 | 0.028 | 0/2 | 1.00 |
| +5° | 82.0 (72.0–92.0) | 22.0 (12.0–34.0) | 94.9 (86.8–100.0) | 2.0 | 0.028 | 0/1 | 1.00 |
| +10° | 82.0 (70.0–92.0) | 18.0 (8.0–28.0) | 92.7 (84.2–100.0) | 4.0 | 0.059 | 0/1 | 1.00 |
| +15° | 80.0 (68.0–90.0) | 18.0 (8.0–30.0) | 90.2 (80.0–97.7) | 8.0 | 0.092 | 1/1 | 1.00 |
| +20° | 80.0 (68.0–90.0) | 26.0 (14.0–40.0) | 89.2 (78.4–97.4) | 12.0 | 0.166 | 2/2 | 1.00 |
| +30° | 70.0 (56.0–82.0) | 48.0 (34.0–62.0) | 84.6 (69.6–96.3) | 28.0 | 0.242 | 6/1 | 1.00 |
| +45° | 46.0 (32.0–60.0) | 54.0 (40.0–68.0) | 65.2 (44.0–83.3) | 54.0 | 0.387 | 19/2 | 0.003 |
| **Rotation, superior–inferior axis** | | | | | | | |
| -45° | 72.0 (60.0–84.0) | 56.0 (42.0–70.0) | 90.9 (76.9–100.0) | 20.0 | 0.244 | 5/1 | 1.00 |
| -30° | 78.0 (66.0–90.0) | 28.0 (16.0–42.0) | 83.3 (70.6–94.6) | 14.0 | 0.128 | 3/2 | 1.00 |
| -20° | 76.0 (64.0–88.0) | 24.0 (12.0–36.0) | 86.8 (75.0–95.6) | 8.0 | 0.064 | 3/1 | 1.00 |
| -15° | 78.0 (66.0–90.0) | 20.0 (10.0–32.0) | 85.0 (72.7–95.0) | 8.0 | 0.054 | 2/1 | 1.00 |
| -10° | 76.0 (64.0–88.0) | 24.0 (14.0–36.0) | 89.5 (79.0–97.6) | 4.0 | 0.030 | 2/0 | 1.00 |
| -5° | 80.0 (68.0–90.0) | 18.0 (8.0–28.0) | 92.7 (83.7–100.0) | 0.0 | 0.032 | 0/0 | 1.00 |
| +5° | 84.0 (74.0–94.0) | 16.0 (6.0–26.0) | 92.9 (84.1–100.0) | 6.0 | 0.033 | 0/2 | 1.00 |
| +10° | 82.0 (70.0–92.0) | 22.0 (12.0–34.0) | 97.4 (91.7–100.0) | 4.0 | 0.049 | 0/1 | 1.00 |
| +15° | 84.0 (74.0–94.0) | 26.0 (14.0–38.0) | 97.3 (91.2–100.0) | 8.0 | 0.064 | 0/2 | 1.00 |
| +20° | 84.0 (74.0–94.0) | 22.0 (12.0–34.0) | 94.9 (87.2–100.0) | 8.0 | 0.099 | 0/2 | 1.00 |
| +30° | 76.0 (62.0–88.0) | 28.0 (16.0–42.0) | 86.1 (74.2–97.1) | 16.0 | 0.160 | 3/1 | 1.00 |
| +45° | 70.0 (56.0–82.0) | 46.0 (32.0–60.0) | 85.2 (70.4–96.7) | 22.0 | 0.248 | 7/2 | 1.00 |

**Supplementary Table 27 | Severity trends and abstention under perturbation.** a, Logistic generalized estimating equations (Supplementary Note 8) for correctness without abstention and for abstention; the odds ratio compares the highest severity with no perturbation. b, Errors introduced by perturbation (incorrect perturbed results in subjects whose unperturbed prediction was correct) and the number that led to abstention; AUC-ROC of prediction confidence for identifying incorrect results; accuracy on predicted cases; and the proportion of all results returned as an incorrect prediction. Ghosting and motion are pooled across levels, and rotation across axes and directions.

| **a, Trend across severity** | **Outcome** | **Odds ratio (95% CI)** | **P** | | |
|---|---|---|---|---|---|
| Ghosting | Correct without abstention | 0.52 (0.31–0.87) | 0.013 | | |
| Ghosting | Abstained | 1.23 (0.67–2.27) | 0.500 | | |
| Motion | Correct without abstention | 0.49 (0.30–0.82) | 0.006 | | |
| Motion | Abstained | 2.55 (1.23–5.28) | 0.012 | | |
| Rotation, left-right axis | Correct without abstention | 0.14 (0.07–0.31) | <0.001 | | |
| Rotation, left-right axis | Abstained | 3.64 (1.73–7.63) | <0.001 | | |
| Rotation, anterior-posterior axis | Correct without abstention | 0.20 (0.08–0.48) | <0.001 | | |
| Rotation, anterior-posterior axis | Abstained | 7.65 (3.03–19.32) | <0.001 | | |
| Rotation, superior-inferior axis | Correct without abstention | 0.56 (0.31–0.98) | 0.043 | | |
| Rotation, superior-inferior axis | Abstained | 5.05 (2.35–10.84) | <0.001 | | |
| **b, Errors introduced by perturbation** | **Errors introduced, n** | **Abstained, n (%)** | **AUC-ROC of confidence for incorrect results** | **Accuracy on predicted, %** | **Predicted incorrectly, % of all cases** |
| No perturbation (n = 50) | — | — | 0.97 | 95.1 | 4.0 |
| Ghosting (150 results) | 14 | 6 (42.9) | 0.89 | 85.2 | 11.3 |
| Motion (150 results) | 23 | 15 (65.2) | 0.86 | 81.7 | 12.7 |
| Rotation up to ±20° (1,200 results) | 34 | 19 (55.9) | 0.86 | 90.2 | 7.8 |
| Rotation 30–45° (600 results) | 118 | 73 (61.9) | 0.75 | 76.9 | 13.8 |

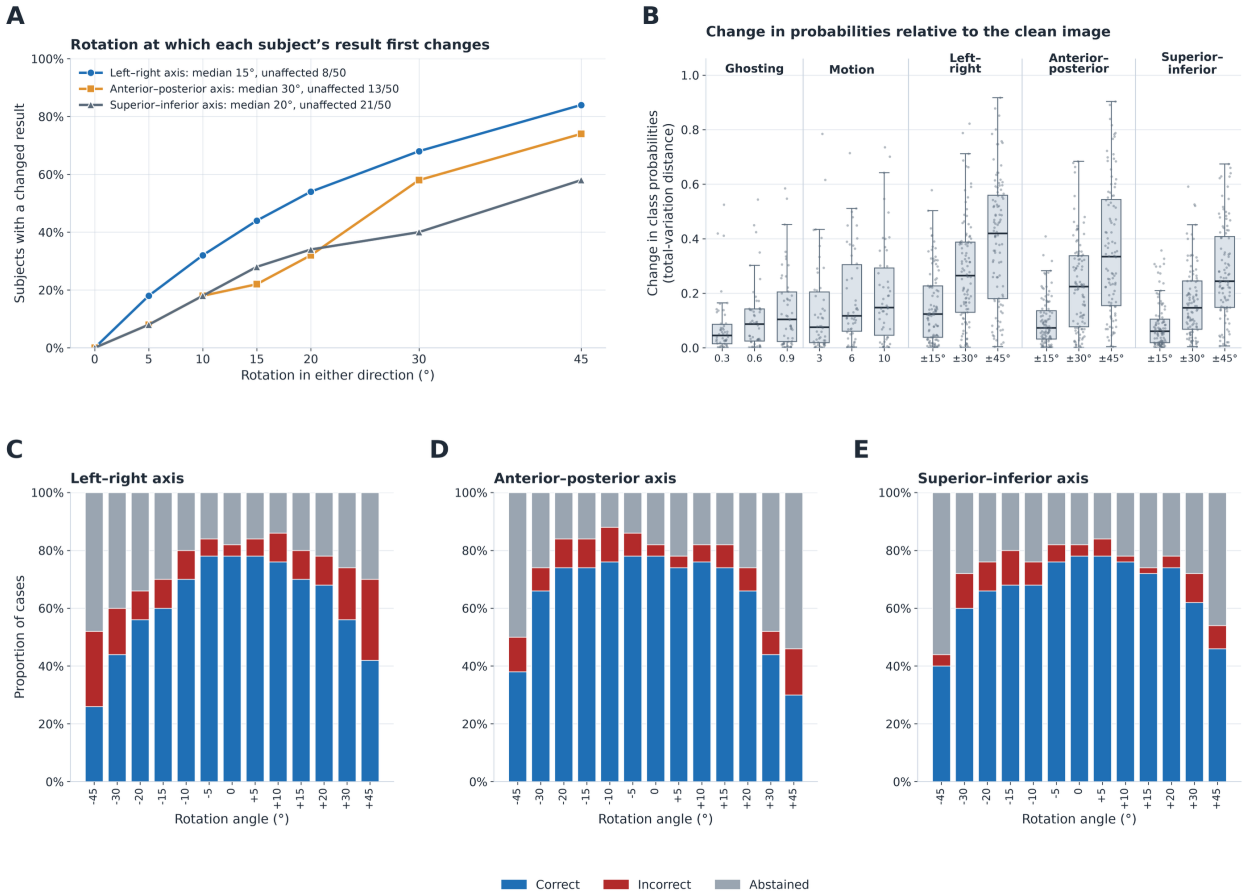


**Supplementary Fig. 10 | Rotation sensitivity and change in class probabilities (n = 50). a**, Cumulative proportion of subjects whose result changed, through a different predicted class or a new abstention, by rotation magnitude about each axis; the legend gives the median breaking angle and the number of subjects unaffected up to ±45°. **b**, Change in the calibrated class-probability vector relative to the same subject's unperturbed result (total-variation distance) at each ghosting and motion level and for rotation of ±15°, ±30° and ±45° about each axis, pooling both directions; boxes show median and IQR. **c–e,** Correct, incorrect and abstained cases at each angle about the left–right (c), anterior–posterior (d) and superior–inferior (e) axes; angle 0 is the unperturbed reference.

#### Supplementary Note 9 | Independent evaluation at Macquarie University

Forty subjects were processed end to end at Macquarie University on the site's own infrastructure through the complete PedDx framework. The cohort comprised medulloblastoma (n = 20), LGG (n = 11) and DMG (n = 9); it contained no ATRT or ependymoma. Per-class performance is reported without abstention, with every case assigned to its most probable class, and with abstention, on the 37 cases for which a prediction was returned (Supplementary Table 28). Probabilities are the calibrated outputs of the multiclass softmax on which the abstention rule operates, and macro averages are taken over the three classes. The confusion matrices are shown in Supplementary Fig. 11.

**Supplementary Table 28 | Per-class performance by the team at Macquarie University on WCH data without and with abstention.** Without abstention, all 40 cases are assigned to the most probable class; with abstention, metrics are computed on the 37 predicted cases. Metrics are one-vs-rest at the arg-max label, and AUC-ROC uses the continuous class probability.

| | Class | n | Balanced accuracy | F1 | AUC-ROC | Sensitivity | Specificity |
|---|---|---|---|---|---|---|---|
| Without abstention (all 40 cases) | DMG/DIPG | 9 | 0.944 | 0.941 | 0.975 | 0.889 | 1.000 |
| | LGG | 11 | 0.937 | 0.909 | 0.991 | 0.909 | 0.966 |
| | Medulloblastoma | 20 | 0.925 | 0.927 | 0.968 | 0.950 | 0.900 |
| | Macro average (3 classes) | 40 | 0.936 | 0.926 | 0.978 | 0.916 | 0.955 |
| With abstention (37 predicted cases) | DMG/DIPG | 9 | 0.944 | 0.941 | 0.984 | 0.889 | 1.000 |
| | LGG | 10 | 1.000 | 1.000 | 1.000 | 1.000 | 1.000 |
| | Medulloblastoma | 18 | 0.974 | 0.973 | 0.971 | 1.000 | 0.947 |
| | Macro average (3 classes) | 37 | 0.973 | 0.971 | 0.985 | 0.963 | 0.982 |

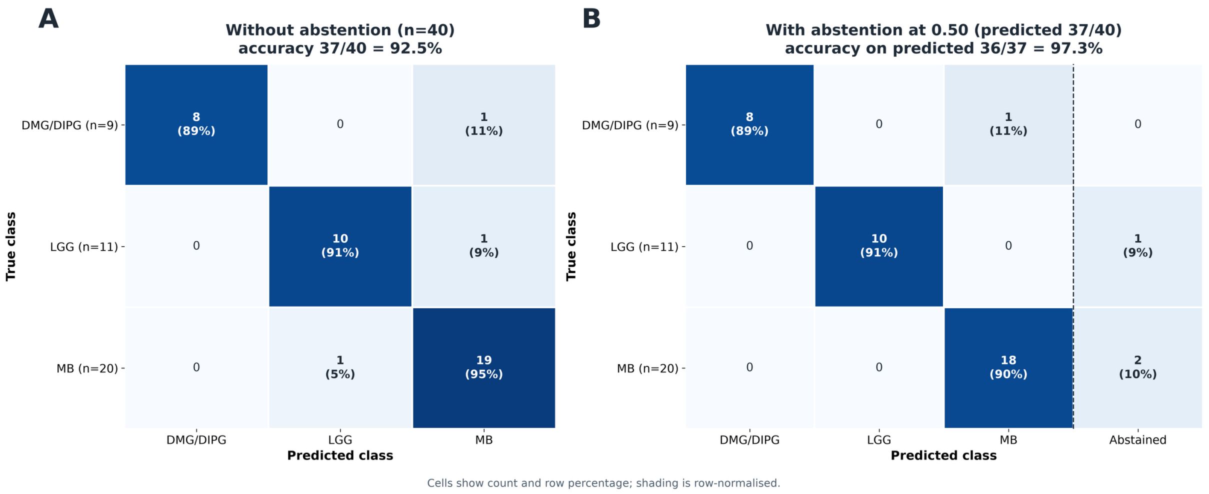


**Supplementary Fig. 11 | Confusion matrices at Macquarie University (n = 40).** a, Without abstention, every case assigned to its most probable class (accuracy 37 of 40, 92.5%). b, With abstention at a calibrated probability of 0.50, abstained cases shown in the final column (accuracy on predicted cases 36 of 37, 97.3%). Cells give the count and the percentage of the row; shading is row-normalised. DMG/DIPG, diffuse midline glioma; MB, medulloblastoma.

The quality-control agent's verdicts were compared with the severity of the deterministic findings, with diagnostic correctness and with the model's own uncertainty (Supplementary Fig. 12). Every invalidating finding led to ESCALATE.

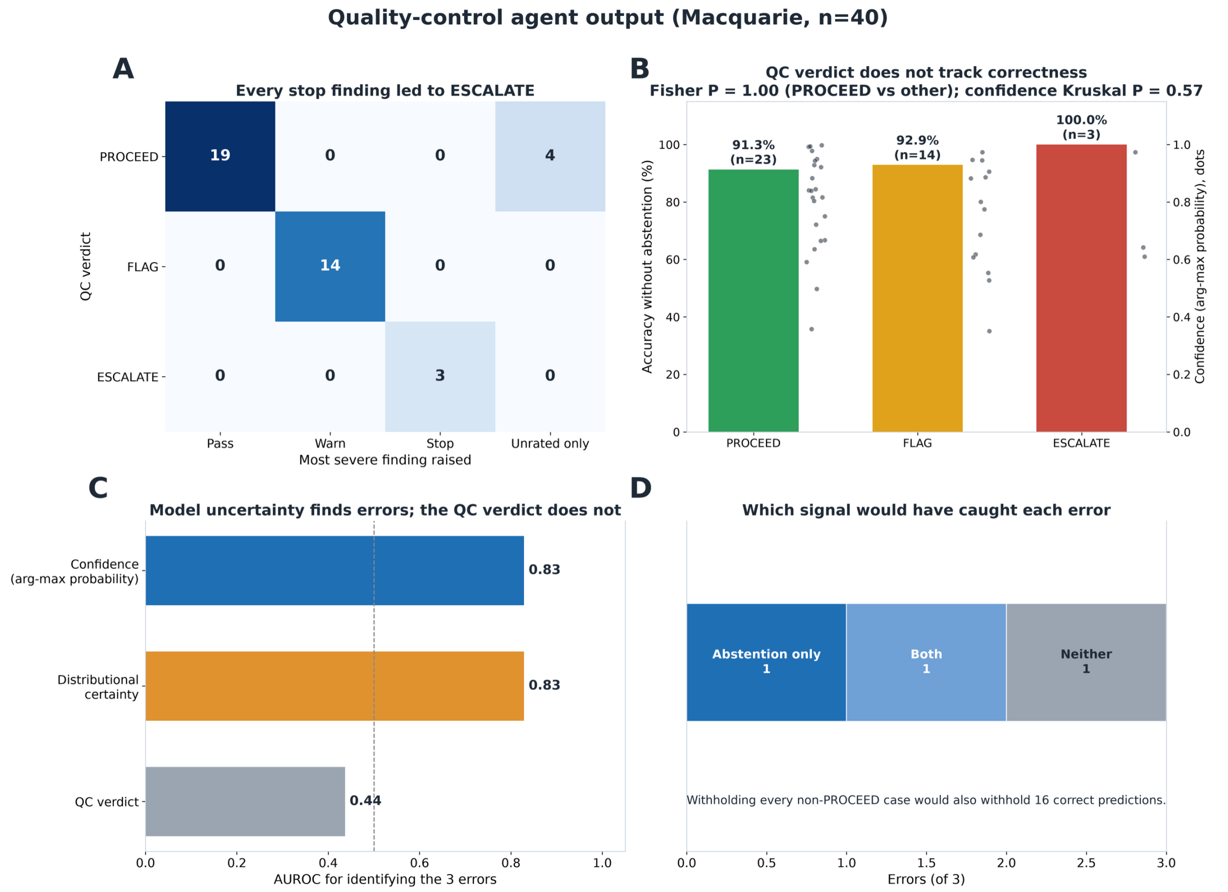


**Supplementary Fig. 12 | Quality-control agent output at Macquarie University (n = 40).** a, Agent verdict against the most severe finding raised by the deterministic checks. b, Accuracy without abstention by verdict (bars) with each case's prediction confidence (dots); the Fisher exact P compares PROCEED with FLAG or ESCALATE, and the Kruskal–Wallis (ref. S22) P compares confidence across verdicts. c, AUC-ROC of prediction confidence, distributional certainty and the ordinal quality-control verdict for identifying the 3 classification errors; the dashed line marks chance. d, Errors that would have been intercepted by abstention only, by a non-PROCEED verdict only, by both or by neither.

### Supplementary Note 10 | Anatomical region and multi-crop sampling

**Methods.** The pretrained backbone operates on fixed 112 × 112 × 112 voxel inputs, whereas the registered T2-weighted volumes have dimensions of 240 × 240 × 155 voxels. The $112^3$ input size was retained to match the spatial dimensions of the released pretrained positional embeddings, which yield a 7 × 7 × 7 grid of 343 patch tokens at the stride-16 patch embedding. Alternative input dimensions would require interpolation of the pretrained positional encoding and were therefore not used. Direct resizing of the complete registered volume to $112^3$ was evaluated but reduced the effective spatial resolution to approximately 2.14 × 2.14 × 1.38 $mm^3$. We therefore evaluated anatomically restricted inputs centered on the posterior fossa, which provided an effective resolution of approximately 1.14 × 1.00 × 0.68 mm after resizing to the backbone input dimensions.

The posterior fossa region was defined directly in SRI24 template space. Its anatomical coordinates were specified before model fitting and were not modified according to classification performance. Because all study volumes were rigidly registered to the same SRI24 template, identical voxel coordinates corresponded to a consistent anatomical region across subjects and were applied unchanged across cohorts. The posterior fossa extent on the template was identified as $i$ = 65–178, $j$ = 105–205 and $k$ = 0–62. To provide additional coverage for residual inter-subject variation after registration, this region was expanded to $i \in [56, 184)$, $j \in [100, 212)$ and $k \in [0, 76)$, producing a 128 × 112 × 76 voxel posterior fossa box and approximately 9 mm of additional lateral coverage on each side of the cerebellum. The anatomical extent of this region and the candidate extraction regions are shown on the SRI24 template in axial, coronal and sagittal views (Supplementary Fig. 13).

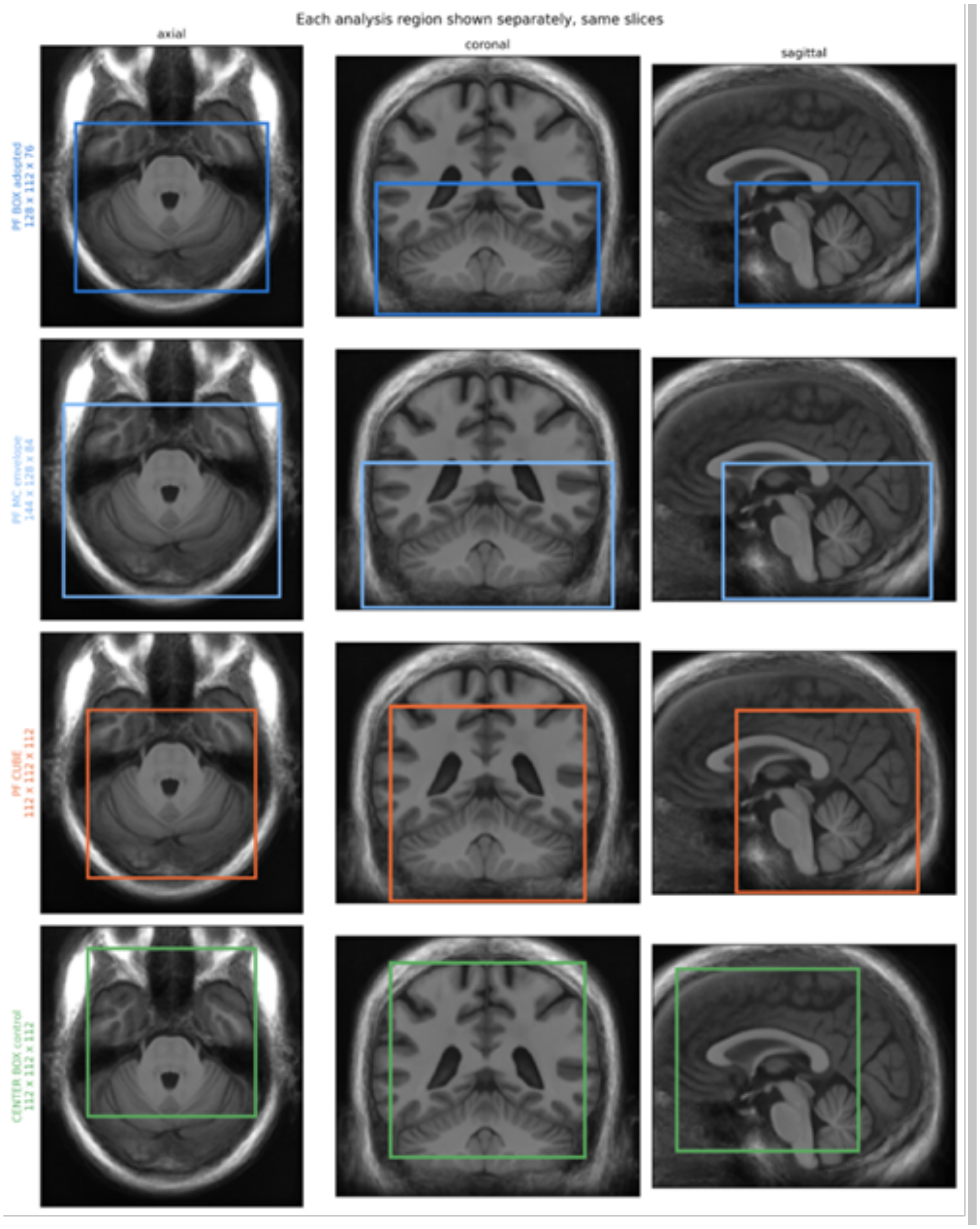


**Supplementary Fig. 13 | Anatomical definition of posterior fossa sampling regions.** Axial, coronal and sagittal views of the SRI24 template showing the adopted posterior fossa base region (PF box; 128 × 112 × 76 voxels), the total spatial envelope covered by the 18 deterministic translations used for posterior fossa multi-crop sampling (PF_MC; 144 × 128 × 84 voxels), a native $112^3$ posterior fossa cube (PF_C), and a $112^3$ head-centered control region. All panels show identical anatomical slice locations.

A native $112^3$ voxel cube could not provide a comparably tight posterior fossa field of view. The foramen magnum lies at the inferior boundary of the registered image volume, such that the crop cannot be extended further inferiorly beyond $k = 0$. Consequently, a 112-voxel superior-inferior extent beginning at the field-of-view boundary necessarily extends approximately 36 mm superior to the defined posterior fossa region and includes additional supratentorial tissue. The 128 × 112 × 76 posterior fossa box was therefore resized to the $112^3$ backbone input. This transformation is anisotropic, corresponding to scaling factors of 0.875 along the i axis, 1.0 along the j axis and 1.474 along the k axis.

Six candidate extraction strategies were compared by nested cross-validation using the discovery cohort, with their definitions and comparative performance reported in Supplementary Table 29 and Supplementary Tables 30a,b, respectively. These experiments were designed to distinguish the effect of anatomical localization from the effect of resampling and multi-view feature aggregation. One posterior fossa control, PF_C, consisted of a native 112 × 112 × 112 voxel cube and therefore required no spatial resizing before backbone input, allowing assessment of whether the interpolation required for the anatomically constrained posterior fossa box materially affected classification performance. Head-centered extraction strategies, C112 and C_MC, were included as anatomical controls. These regions were centered on the head rather than the posterior fossa; their inferior boundary began at $k = 21$ and consequently excluded portions of the posterior cerebellum and inferior posterior fossa. The spatial relationship between the posterior fossa box, the multi-crop sampling envelope, the native $112^3$ posterior fossa cube and the head-centered control region is shown in Supplementary Fig. 13.

The posterior fossa multi-crop strategy, denoted PF_MC, was adopted for model development and evaluation. PF_MC uses the expanded posterior fossa box as its base region and samples it at 18 fixed translational offsets forming a deterministic three-dimensional lattice:

$$(\Delta\mathrm{i},\ \Delta\mathrm{j},\ \Delta\mathrm{k})\ \in\ \{-8,\ 0,\ 8\}\ \times\ \{-8,\ 0,\ 8\}\ \times\ \{0,\ 8\}\ voxels$$

The three positions along each of the i and j axes and two positions along the k axis produce 3 × 3 × 2 = 18 views per subject. The inferior boundary of the posterior fossa coincides with the inferior field-of-view boundary, and negative translations along k would therefore extend beyond the available image volume. The inferior edge was consequently clamped at k = 0, resulting in two rather than three translation levels along this axis. Across all offsets, the 18 crops span a 144 × 128 × 84 voxel sampling envelope around the base 128 × 112 × 76 posterior fossa region.

An offset magnitude of 8 voxels was selected relative to the backbone's 16-voxel patch stride. An 8-voxel translation corresponds to half of one patch stride and alters the alignment of anatomical structures relative to the patch-tokenization grid between views. By contrast, a translation of 16 voxels would shift the image by a complete patch stride and substantially restore the same relative anatomy-to-patch alignment. The offsets were fixed and identical for all subjects; multi-crop sampling was therefore implemented as deterministic sampling of the predefined lattice rather than as stochastic data augmentation.

Percentiles for the crop normalization (main text, Methods) were computed over brain voxels rather than all voxels, a departure from the reference implementation, because about 70% of the posterior fossa box is zero-valued background that would otherwise dominate the intensity range.

**Supplementary Table 29 | Candidate anatomical extraction regions.** Effective voxel size is the physical extent represented by one voxel of the $112^3$ input after resampling.

| Arm | Definition | Box (SRI24 voxels) | Native crop | Resized to $112^3$ | Effective voxel (mm) |
|---|---|---|---|---|---|
| **PF_MC** | Posterior fossa box, shifted by ±8 voxels into 18 positions, each encoded separately and the 18 feature vectors averaged. | i[56:184] j[100:212] k[0:76] | 128 × 112 × 76, ×18 | Yes, each crop | 1.14 × 1.00 × 0.68 |

| Arm | Definition | Box (SRI24 voxels) | Native crop | Resized to $112^3$ | Effective voxel (mm) |
|---|---|---|---|---|---|
| **PF_T** | Identical box, no jitter. One crop. | i[56:184] j[100:212] k[0:76] | 128 × 112 × 76 | Yes, trilinear | 1.14 × 1.00 × 0.68 |
| **PF_C** | Posterior fossa anchored but extended 36 mm superiorly, so no resampling is needed. | i[64:176] j[100:212] k[0:112] | 112 × 112 × 112 | No | 1.00 isotropic |
| **C_MC** | Middle of the head, with PF_MC's exact jitter lattice. | i[64:176] j[64:176] k[21:133] | 112 × 112 × 112, ×18 | No | 1.00 isotropic |
| **C112** | Same center box as C_MC, no jitter. | i[64:176] j[64:176] k[21:133] | 112 × 112 × 112 | No | 1.00 isotropic |
| **G112** | The entire volume squashed to $112^3$. Everything is present, at roughly 2 mm resolution, with about 77% of tokens spent on background. | whole volume | 240 × 240 × 155 | Yes | 2.14 × 2.14 × 1.38 |

**Results.** Regions differed significantly (Friedman test (ref. S21), $\chi^2 = 44.5$, df = 5, $p = 1.9 \times 10^{-8}$) across the ten representation × head contexts (2 representations × 5 heads), with each region scored in all ten so that the comparison is paired (main text, Methods).

**Supplementary Table 30a | Extraction-region ranking on discovery,** averaged across all ten representation × head contexts.

| Arm | Mean rank (1 = best of 6) | Best AUC | Mean AUC | Crops |
|---|---|---|---|---|
| PF_MC | 1.40 | 0.892 | 0.870 | 18 |
| PF_T | 1.90 | 0.887 | 0.854 | 1 |
| C_MC | 2.80 | 0.866 | 0.832 | 18 |
| G112 | 4.00 | 0.837 | 0.799 | 1 |
| C112 | 5.10 | 0.802 | 0.782 | 1 |
| PF_C | 5.80 | 0.804 | 0.768 | 1 |

Ten of fifteen pairwise comparisons survived Holm correction. Both posterior fossa arms (PF_MC, PF_T) outperformed the three arms that either omit the posterior fossa or dilute it with supratentorial tissue (C112, G112, PF_C) in all ten contexts, with no reversal in direction in any context.

**Supplementary Table 30b | PF_MC against each alternative, aggregated across contexts.** A positive delta favors PF_MC.

| Versus region | Description | Δ AUC | Holm P | Separated |
|---|---|---|---|---|
| C112 | centered crop | +0.0926 | 0.0293 | yes |
| G112 | whole brain, downsampled | +0.0675 | 0.0293 | yes |
| PF_T | posterior fossa, single centered crop | +0.0150 | 0.7539 | no |
| PF_C | posterior fossa, cropped without resizing | +0.0944 | 0.0293 | yes |
| C_MC | multi-crop, not confined to the posterior fossa | +0.0319 | 0.0293 | yes |

PF_MC outperformed C_MC which applies the identical 18-offset lattice centered on the head rather than the posterior fossa in 10 of 10 contexts (median Δ = 0.032 macro AUC, Holm-adjusted $p = 0.029$). Since the two arms differ only in placement, not in crop count or lattice geometry, this indicates that the gain associated with PF_MC reflects anatomical placement rather than multi-crop averaging itself.

PF_C, the only posterior-fossa arm requiring no resampling, ranked last of the six arms, below C112. PF_T outperformed PF_C in 10 of 10 contexts (Δ = 0.092, Holm-adjusted $p = 0.029$). A native $112^3$ cube anchored at the foramen magnum must extend approximately 36 mm above the tentorium to avoid resampling, so the PF_C box necessarily incorporates substantial supratentorial tissue; its lower performance is therefore attributable to this anatomical dilution rather than to the interpolation that a native crop avoids.

PF_MC and PF_T were not statistically distinguishable. The median difference favored PF_MC by 0.015 macro AUC, but the direction was consistent in only 4 of 10 contexts (sign test $p = 0.75$, Holm-adjusted $p = 0.75$). PF_MC was adopted on the basis of its first-place rank and the absence of any context in which it performed worse, rather than on a demonstrated advantage over PF_T. PF_T achieves comparable accuracy at one-eighteenth of the extraction cost and may be preferable where that cost is a constraint. The multi-crop question is returned to at the geometry stage (Supplementary Note 2), where it is settled at the fully adopted configuration.

#### Supplementary Note 11 | Comparison of final-block [CLS] and pooled patch-token representations

**Methods.** For each crop, one forward pass yielded the class (CLS) token and the mean of the 343 patch tokens of the last transformer block. With the region fixed at PF_MC, the CLS token alone (1,024 dimensions) was compared with its concatenation with the mean patch token (2,048 dimensions) in each of the five heads (Supplementary Table 31).

**Supplementary Table 31 | Representation within the adopted region (PF_MC rows).** A positive Δ favors the 2048-dimensional concatenation.

| Head | CLS (1024) | CLS+ pool (2048) | Δ | 95% CI | Holm P |
|---|---|---|---|---|---|
| Linear probe | 0.8834 | 0.8918 | +0.0084 | [−0.0032, +0.0228] | 1.000 |
| Linear (one-vs-rest) | 0.8606 | 0.8553 | −0.0052 | [−0.0194, +0.0083] | 1.000 |
| PCA–logistic regression | 0.8771 | 0.8684 | −0.0086 | [−0.0241, +0.0063] | 1.000 |
| MLP | 0.8854 | 0.8760 | −0.0094 | [−0.0313, +0.0102] | 1.000 |
| MLP (one-vs-rest) | 0.8551 | 0.8430 | −0.0121 | [−0.0374, +0.0150] | 1.000 |

**Results.** No head showed a significant difference. Differences ranged from −0.0121 to +0.0084 and every interval spanned zero. The smaller representation was adopted: [CLS] alone, halving the feature vector from 2048 to 1024 dimensions at no measurable cost in discrimination.

#### Supplementary Note 12 | Classification head comparison within PF_MC and final-block [CLS] features

**Methods.** All heads were trained on the frozen, crop-averaged final-block [CLS] representation.

Five classification heads were evaluated within the discovery cohort. Three directly modeled the five diagnostic classes: (1) a linear probe consisting of a single linear layer with a five-class softmax output; (2) multinomial logistic regression applied after feature standardization and principal-component analysis (PCA); and (3) a multilayer perceptron (MLP) comprising one or two hidden linear layers, each followed by GELU (ref. S29) activation and dropout, and a final five-class output layer. Two additional one-vs-rest configurations were evaluated using five independently fitted binary classifiers, architecturally corresponding to the linear and MLP heads, respectively. The search space and results are given in Supplementary Tables 32 and 33.

The linear and MLP heads were optimized for 300 epochs using AdamW (ref. S27) with a cosine learning-rate schedule (ref. S28). The multinomial logistic-regression model was optimized using L-BFGS (ref. S30) after feature standardization and PCA dimensionality reduction.

**Supplementary Table 32 | Hyperparameter search space.** Values separated by commas were searched; a single value was fixed. Em dash indicates the parameter does not apply to that head.

| Hyperparameter | Linear | PCA + LR | MLP | Linear (OvR) | MLP (OvR) |
|---|---|---|---|---|---|
| Learning rate | $1\times10^{-4}$, $1\times10^{-3}$, $1\times10^{-2}$ | — | $1\times10^{-4}$, $1\times10^{-3}$ | $1\times10^{-4}$, $1\times10^{-3}$, $1\times10^{-2}$ | $1\times10^{-4}$, $1\times10^{-3}$ |
| Weight decay | 0, $1\times10^{-4}$ | — | $1\times10^{-4}$ | 0, $1\times10^{-4}$ | $1\times10^{-4}$ |
| Hidden blocks | 0 | — | 1, 2 | 0 | 1, 2 |
| Hidden units | — | — | 256, 512 | — | 256 |
| Dropout probability | — | — | 0.3, 0.5 | — | 0.3, 0.5 |
| Feature standardization | no, yes | applied | yes | yes | yes |
| Class weighting | none, balanced | balanced | none, balanced | none, balanced | none, balanced |
| PCA components | — | 128, 256 | — | — | — |

| Hyperparameter | Linear | PCA + LR | MLP | Linear (OvR) | MLP (OvR) |
|---|---|---|---|---|---|
| Inverse regularization C | — | $1\times10^{-3}$, $1\times10^{-2}$, 0.1, 1, 10 | — | — | — |

**Results.** With the region and representation fixed at PF_MC and [CLS] alone, the five heads were compared pairwise (10 comparisons).

**Supplementary Table 33 | Head comparisons within PF_MC and the final-block CLS token. Shaded rows were separated. P values are Holm-adjusted across the ten comparisons and separation is based on the adjusted value; 95% confidence intervals are unadjusted.**

| Comparison | A | B | Δ | 95% CI | Raw P | Holm P | Separated |
|---|---|---|---|---|---|---|---|
| **Linear probe vs MLP (one-vs-rest)** | 0.8834 | 0.8551 | +0.0283 | [+0.0122, +0.0458] | 0.000 | 0.000 | yes |
| **Linear probe vs Linear (one-vs-rest)** | 0.8834 | 0.8606 | +0.0228 | [+0.0081, +0.0368] | 0.004 | 0.036 | yes |
| **Linear (one-vs-rest) vs MLP** | 0.8606 | 0.8854 | -0.0248 | [-0.0470, -0.0026] | 0.024 | 0.192 | no |
| **MLP vs MLP (one-vs-rest)** | 0.8854 | 0.8551 | +0.0303 | [+0.0046, +0.0547] | 0.024 | 0.192 | no |
| **Linear (one-vs-rest) vs PCA–logistic regression** | 0.8606 | 0.8771 | -0.0165 | [-0.0349, +0.0005] | 0.057 | 0.324 | no |
| **PCA–logistic regression vs MLP (one-vs-rest)** | 0.8771 | 0.8551 | +0.0220 | [-0.0004, +0.0447] | 0.054 | 0.324 | no |
| **Linear probe vs PCA–logistic regression** | 0.8834 | 0.8771 | +0.0063 | [-0.0072, +0.0188] | 0.342 | 1.000 | no |
| **Linear probe vs MLP** | 0.8834 | 0.8854 | -0.0020 | [-0.0231, +0.0193] | 0.858 | 1.000 | no |
| **Linear (one-vs-rest) vs MLP (one-vs-rest)** | 0.8606 | 0.8551 | +0.0055 | [-0.0140, +0.0264] | 0.599 | 1.000 | no |
| **PCA–logistic regression vs MLP** | 0.8771 | 0.8854 | -0.0083 | [-0.0285, +0.0129] | 0.454 | 1.000 | no |

Two of the ten pairwise comparisons survived Holm correction, and both contrast a multiclass head against a one-vs-rest counterpart: Linear probe versus MLP (one-vs-rest) (Δ = +0.0283, 95% CI +0.0122 to +0.0458, Holm $p < 0.001$) and Linear probe versus Linear (one-vs-rest) (Δ = +0.0228, 95% CI +0.0081 to +0.0368, Holm $p = 0.036$).

No comparison among the three multiclass heads was distinguishable, Linear probe versus MLP Δ = −0.0020 (95% CI −0.0231 to +0.0193, Holm $p = 1.000$), Linear probe versus PCA–logistic regression Δ = +0.0063 (95% CI −0.0072 to +0.0188, Holm $p = 1.000$) and the three multiclass heads span only 0.008 macro AUC in total. The detectable effect is therefore one of head family, not architecture. The multiclass MLP head was adopted as the top-ranked multiclass configuration at this stage, with a macro-AUC of 0.8854. Three macro-AUC values are quoted for this configuration elsewhere in the paper and each belongs to a different protocol: 0.8854 is the nested cross-validated estimate used for model selection (Supplementary Notes 10–12); 0.8995 is the pooled out-of-fold estimate obtained when a fresh head is refitted per fold for the sampling-geometry analysis (Supplementary Note 2); and 0.901 ± 0.023 is the mean across the five deployment folds of the calibrated model, reported in the main text.

### Supplementary Note 13 | Contribution of pretraining

Across 60 matched model configurations, the pretrained backbone consistently outperformed the randomly initialized backbone. The median improvement was +0.114 in macro-AUC (range, +0.040 to +0.174) and +0.152 in macro-F1, with higher performance observed for the pretrained model in all 60 comparisons (Supplementary Table 34).

For the final adopted configuration (PF_MC, final-block [CLS], multiclass MLP), pretraining improved macro-AUC from 0.766 to 0.885 and the five-way (argmax) macro-F1 from 0.404 to 0.619. These are model-selection-grid values; the five-way macro-F1 of the deployed configuration is 0.604, and the one-vs-rest macro-F1 in Table 2 of the main text (0.526) is a different metric. Performance for all remaining matched configurations is reported in Supplementary Table 34.

The randomly initialized backbone nevertheless achieved macro-AUC values of 0.653–0.766 across configurations, indicating that the observed effect reflects a consistent benefit of pretraining rather than an absence of predictive signal from the untrained architecture.

**Supplementary Table 34 | Contribution of pretraining across all 60 matched configurations (six extraction regions × two representations × five heads).** Each row pairs one configuration trained on features from the pretrained 3DINO backbone with the identical configuration trained on features from a randomly initialized backbone of the same architecture, under the same folds and the same hyperparameter search. Macro-AUC and macro-F1 are nested cross-validated discovery estimates; macro-F1 is computed on the five-way argmax.

| Region | Representation | Head | Macro-AUC, random init | Macro-AUC, pretrained | Δ AUC | Macro-F1, random init | Macro-F1, pretrained | Δ F1 |
|---|---|---|---|---|---|---|---|---|
| C112 | [CLS] + pooled patch | Linear probe | 0.6736 | 0.7962 | +0.1226 | 0.3233 | 0.5049 | +0.1816 |
| C112 | [CLS] + pooled patch | Linear (one-vs-rest) | 0.6820 | 0.7865 | +0.1044 | 0.3466 | 0.5232 | +0.1765 |
| C112 | [CLS] + pooled patch | MLP | 0.6776 | 0.8022 | +0.1246 | 0.3592 | 0.4692 | +0.1100 |
| C112 | [CLS] + pooled patch | MLP (one-vs-rest) | 0.6601 | 0.7656 | +0.1054 | 0.3479 | 0.5117 | +0.1638 |
| C112 | [CLS] + pooled patch | PCA–logistic regression | 0.6827 | 0.7753 | +0.0926 | 0.3803 | 0.4846 | +0.1043 |
| C112 | [CLS], final block | Linear probe | 0.6892 | 0.7767 | +0.0875 | 0.3978 | 0.4913 | +0.0935 |
| C112 | [CLS], final block | Linear (one-vs-rest) | 0.6968 | 0.7517 | +0.0549 | 0.3769 | 0.4706 | +0.0937 |
| C112 | [CLS], final block | MLP | 0.6833 | 0.7926 | +0.1093 | 0.3680 | 0.4601 | +0.0921 |
| C112 | [CLS], final block | MLP (one-vs-rest) | 0.6679 | 0.7883 | +0.1204 | 0.3850 | 0.5193 | +0.1342 |
| C112 | [CLS], final block | PCA–logistic regression | 0.6910 | 0.7846 | +0.0936 | 0.3964 | 0.4904 | +0.0940 |
| C_MC | [CLS] + pooled patch | Linear probe | 0.7164 | 0.7992 | +0.0828 | 0.3723 | 0.5496 | +0.1773 |
| C_MC | [CLS] + pooled patch | Linear (one-vs-rest) | 0.7225 | 0.8204 | +0.0979 | 0.3861 | 0.5303 | +0.1442 |
| C_MC | [CLS] + pooled patch | MLP | 0.7171 | 0.8222 | +0.1052 | 0.3728 | 0.5246 | +0.1518 |
| C_MC | [CLS] + pooled patch | MLP (one-vs-rest) | 0.7033 | 0.8136 | +0.1103 | 0.3768 | 0.5215 | +0.1447 |
| C_MC | [CLS] + pooled patch | PCA–logistic regression | 0.7192 | 0.8371 | +0.1180 | 0.3835 | 0.5297 | +0.1462 |
| C_MC | [CLS], final block | Linear probe | 0.7211 | 0.8560 | +0.1349 | 0.4123 | 0.5346 | +0.1223 |
| C_MC | [CLS], final block | Linear (one-vs-rest) | 0.7315 | 0.8174 | +0.0859 | 0.3852 | 0.5402 | +0.1551 |
| C_MC | [CLS], final block | MLP | 0.7149 | 0.8662 | +0.1513 | 0.3727 | 0.5503 | +0.1776 |
| C_MC | [CLS], final block | MLP (one-vs-rest) | 0.7077 | 0.8414 | +0.1337 | 0.3809 | 0.5414 | +0.1604 |
| C_MC | [CLS], final block | PCA–logistic regression | 0.7094 | 0.8446 | +0.1352 | 0.3947 | 0.5346 | +0.1399 |
| G112 | [CLS] + pooled patch | Linear probe | 0.6794 | 0.7979 | +0.1185 | 0.3233 | 0.4821 | +0.1587 |

| Region | Representation | Head | Macro-AUC, random init | Macro-AUC, pretrained | Δ AUC | Macro-F1, random init | Macro-F1, pretrained | Δ F1 |
|---|---|---|---|---|---|---|---|---|
| G112 | [CLS] + pooled patch | Linear (one-vs-rest) | 0.6981 | 0.7934 | +0.0953 | 0.3360 | 0.4925 | +0.1565 |
| G112 | [CLS] + pooled patch | MLP | 0.6793 | 0.8373 | +0.1581 | 0.3655 | 0.4901 | +0.1246 |
| G112 | [CLS] + pooled patch | MLP (one-vs-rest) | 0.6531 | 0.7890 | +0.1359 | 0.3407 | 0.4996 | +0.1589 |
| G112 | [CLS] + pooled patch | PCA–logistic regression | 0.6786 | 0.8023 | +0.1237 | 0.3552 | 0.5148 | +0.1596 |
| G112 | [CLS], final block | Linear probe | 0.6838 | 0.8177 | +0.1339 | 0.3579 | 0.4862 | +0.1283 |
| G112 | [CLS], final block | Linear (one-vs-rest) | 0.6853 | 0.7734 | +0.0882 | 0.3400 | 0.4646 | +0.1246 |
| G112 | [CLS], final block | MLP | 0.6958 | 0.8083 | +0.1126 | 0.3596 | 0.4901 | +0.1305 |
| G112 | [CLS], final block | MLP (one-vs-rest) | 0.6628 | 0.7862 | +0.1234 | 0.3588 | 0.4899 | +0.1311 |
| G112 | [CLS], final block | PCA–logistic regression | 0.6907 | 0.7848 | +0.0941 | 0.3432 | 0.4821 | +0.1390 |
| PF_C | [CLS] + pooled patch | Linear probe | 0.6764 | 0.7921 | +0.1157 | 0.3325 | 0.4811 | +0.1486 |
| PF_C | [CLS] + pooled patch | Linear (one-vs-rest) | 0.6695 | 0.7662 | +0.0967 | 0.3668 | 0.4968 | +0.1300 |
| PF_C | [CLS] + pooled patch | MLP | 0.7039 | 0.7881 | +0.0842 | 0.3264 | 0.4836 | +0.1571 |
| PF_C | [CLS] + pooled patch | MLP (one-vs-rest) | 0.6803 | 0.7577 | +0.0773 | 0.3492 | 0.4804 | +0.1312 |
| PF_C | [CLS] + pooled patch | PCA–logistic regression | 0.6717 | 0.7883 | +0.1166 | 0.3434 | 0.4969 | +0.1535 |
| PF_C | [CLS], final block | Linear probe | 0.6668 | 0.7377 | +0.0709 | 0.3473 | 0.4406 | +0.0933 |
| PF_C | [CLS], final block | Linear (one-vs-rest) | 0.6752 | 0.7383 | +0.0631 | 0.3377 | 0.4337 | +0.0960 |
| PF_C | [CLS], final block | MLP | 0.6988 | 0.8039 | +0.1050 | 0.3344 | 0.4708 | +0.1365 |
| PF_C | [CLS], final block | MLP (one-vs-rest) | 0.6922 | 0.7324 | +0.0403 | 0.3390 | 0.4669 | +0.1280 |
| PF_C | [CLS], final block | PCA–logistic regression | 0.6670 | 0.7729 | +0.1059 | 0.3796 | 0.4762 | +0.0966 |
| PF_MC | [CLS] + pooled patch | Linear probe | 0.7490 | 0.8918 | +0.1427 | 0.3989 | 0.6080 | +0.2091 |
| PF_MC | [CLS] + pooled patch | Linear (one-vs-rest) | 0.7543 | 0.8553 | +0.1011 | 0.4138 | 0.6016 | +0.1878 |
| PF_MC | [CLS] + pooled patch | MLP | 0.7536 | 0.8760 | +0.1224 | 0.3787 | 0.6074 | +0.2287 |
| PF_MC | [CLS] + pooled patch | MLP (one-vs-rest) | 0.7232 | 0.8430 | +0.1199 | 0.4236 | 0.5648 | +0.1412 |
| PF_MC | [CLS] + pooled patch | PCA–logistic regression | 0.7446 | 0.8684 | +0.1238 | 0.4250 | 0.6069 | +0.1819 |
| PF_MC | [CLS], final block | Linear probe | 0.7430 | 0.8834 | +0.1404 | 0.4223 | 0.5744 | +0.1521 |
| PF_MC | [CLS], final block | Linear (one-vs-rest) | 0.7547 | 0.8606 | +0.1059 | 0.3967 | 0.5881 | +0.1914 |
| PF_MC | [CLS], final block | MLP | 0.7660 | 0.8854 | +0.1194 | 0.4042 | 0.6192 | +0.2150 |
| PF_MC | [CLS], final block | MLP (one-vs-rest) | 0.7336 | 0.8551 | +0.1215 | 0.4368 | 0.6081 | +0.1714 |
| PF_MC | [CLS], final block | PCA–logistic regression | 0.7462 | 0.8771 | +0.1308 | 0.4296 | 0.6147 | +0.1850 |
| PF_T | [CLS] + pooled patch | Linear probe | 0.7134 | 0.8875 | +0.1741 | 0.3972 | 0.5858 | +0.1886 |
| PF_T | [CLS] + pooled patch | Linear (one-vs-rest) | 0.7286 | 0.8596 | +0.1310 | 0.4037 | 0.5602 | +0.1565 |
| PF_T | [CLS] + pooled patch | MLP | 0.7358 | 0.8847 | +0.1490 | 0.4060 | 0.6075 | +0.2015 |

| Region | Representation | Head | Macro-AUC, random init | Macro-AUC, pretrained | Δ AUC | Macro-F1, random init | Macro-F1, pretrained | Δ F1 |
|---|---|---|---|---|---|---|---|---|
| PF_T | [CLS] + pooled patch | MLP (one-vs-rest) | 0.7169 | 0.8086 | +0.0917 | 0.4213 | 0.5649 | +0.1436 |
| PF_T | [CLS] + pooled patch | PCA–logistic regression | 0.7388 | 0.8763 | +0.1375 | 0.4445 | 0.6054 | +0.1609 |
| PF_T | [CLS], final block | Linear probe | 0.7190 | 0.8576 | +0.1386 | 0.3840 | 0.5593 | +0.1753 |
| PF_T | [CLS], final block | Linear (one-vs-rest) | 0.7489 | 0.8298 | +0.0809 | 0.4086 | 0.5650 | +0.1564 |
| PF_T | [CLS], final block | MLP | 0.7387 | 0.8476 | +0.1089 | 0.4158 | 0.6082 | +0.1925 |
| PF_T | [CLS], final block | MLP (one-vs-rest) | 0.7578 | 0.8154 | +0.0576 | 0.4375 | 0.5863 | +0.1488 |
| PF_T | [CLS], final block | PCA–logistic regression | 0.7490 | 0.8778 | +0.1288 | 0.4261 | 0.5924 | +0.1663 |

#### Supplementary Note 14 | Post-hoc out-of-distribution scores

##### Supplementary Table 35a | Definitions of post-hoc out-of-distribution scores. Score references are given in parentheses (Supplementary References).

| Score | Family | Formula (higher = more OOD) | Estimated from discovery training data | Fitted value |
|---|---|---|---|---|
| MSP (S2) | Probability | $s = -\max_j p_j$ | Nothing | – |
| Entropy | Probability | $s = -\Sigma_j p_j \log p_j$ ($p$ clipped at $10^{-12}$) | Nothing | – |
| GEN (S4) | Probability | $s = \Sigma_j p_j^{\gamma} (1 - p_j)^{\gamma}$ | Nothing | $\gamma = 0.1$ (prespecified) |
| MaxLogit (S5) | Logit | $s = -\max_j l_j$ | Nothing | – |
| Energy (S3) | Logit | $s = -\log \Sigma_j \exp(l_j)$; temperature fixed at 1 | Nothing | $T = 1$ (prespecified) |
| ReAct + Energy (S6) | Logit | $c$ = 90th percentile of all entries of $H_{train}$; $h' = \min(h, c)$; $s = -\log \Sigma \exp(Wh' + b)$ | Clipping threshold $c$, from the flattened training activation matrix (one global threshold, not per unit) | $c = 1.248$ [1.2325, 1.2588] across folds |
| ViM (S7) | Logit | $u = -\mathrm{pinv}(W)b$; $R$ = eigenvectors of $\mathrm{Cov}(h_{train} - u)$ for the $D$ smallest eigenvalues; $\alpha = \Sigma \max_j l_j / \Sigma \lVert R^T(h_{train} - u)\rVert$; $s = \alpha\lVert R^T(h - u)\rVert - \log \Sigma \exp(l)$ | Origin $u$, residual basis $R$, scaling constant $\alpha$ | $D = 128$ of 256 (prespecified); $\alpha = 5.151$ [5.028, 5.226] |
| Mahalanobis (1024-d) (S8) | Feature | $\mu_c$ = class means of $z_{train}$; $\Sigma$ = Ledoit-Wolf estimate of pooled within-class residuals (assume_centered = True); $\Lambda = \mathrm{pinv}(\Sigma)$; $s = \min_c (z - \mu_c)^T \Lambda (z - \mu_c)$ | 5 class means and 1 shared shrunk covariance, in $z$ (1024-d) | LW shrinkage 0.0955 [0.0946, 0.0967] |
| Rel. Mahalanobis (1024-d) (S9) | Feature | Additionally $\mu_0$, $\Sigma_0$ = Ledoit-Wolf over all $z_{train}$; $s = \min_c (d_c^2 - d_0^2)$ | As above plus 1 background mean and covariance, in $z$ | LW shrinkage (background) 0.0907 [0.0898, 0.0920] |
| kNN (1024-d) (S10) | Feature | Bank = L2-normalized $z_{train}$; sim = $k$-th largest cosine similarity; $s = \sqrt{(2 - 2\cdot\mathrm{sim})}$ (Euclidean distance to the $k$-th neighbor on the unit sphere) | Bank of normalized training embeddings, in $z$ | $k = 22 = \lceil\sqrt{492}\rceil$ (prespecified) |
| Mahalanobis (256-d) | Feature | As Mahalanobis (1024-d), computed in $h$ | 5 class means and 1 shared shrunk covariance, in $h$ (256-d) | LW shrinkage 0.0344 [0.0328, 0.0359] |
| Rel. Mahalanobis (256-d) | Feature | As Rel. Mahalanobis (1024-d), computed in $h$ | As above plus background mean and covariance, in $h$ | LW shrinkage (background) 0.0130 [0.0126, 0.0136] |
| kNN (256-d) | Feature | As kNN (1024-d), computed in $h$ | Bank of normalized training embeddings, in $h$ | $k = 22$ (prespecified) |
| Crop cosine spread | Crop | $\bar{x}$ = mean of the 18 raw crop tokens; $s = (1/18)\, \Sigma_i [1 - \cos(x_i, \bar{x})]$ | Nothing (no classifier or standardizer; cannot leak) | – |
| Crop dispersion | Crop | $s = [(1/18)\, \Sigma_i \lVert x_i - \bar{x}\rVert] / \lVert\bar{x}\rVert$ | Nothing (no classifier or standardizer; cannot leak) | – |
| Crop mutual information (S12) | Crop | $p_i$ = softmax of crop $i$ through the head; $\bar{p} = \mathrm{mean}_i\, p_i$; $s = H(\bar{p}) - (1/18)\, \Sigma_i H(p_i)$ | Nothing beyond the head itself | 18 crops; computed in chunks of 64 subjects |
| Crop argmax instability | Crop | $s$ = fraction of the 18 crops whose argmax differs from $\mathrm{argmax}(\bar{p})$ | Nothing beyond the head itself | – |
| Crop energy s.d. | Crop | $s$ = population s.d. over $i$ of $\mathrm{logsumexp}(l_i)$ | Nothing beyond the head itself | Prespecified orientation retained; inverted in the results (AUC-ROC < 0.5) |

| Score | Family | Formula (higher = more OOD) | Estimated from discovery training data | Fitted value |
|---|---|---|---|---|
| NNGuide (S11) | Hybrid | guide = mean of the top-$k$ cosine similarities in $h$; $s$ = −(guide × logsumexp($l$)) | The bank in $h$ | $k$ = 22; assumes logsumexp($l$) > 0, verified on the reference |
| MSP + kNN (rank avg) | Hybrid | $F_m$, $F_k$ = empirical CDFs of MSP and kNN (1024-d) over the 492 discovery out-of-fold scores; $s$ = 0.5 [$F_m$(MSP) + $F_k$(kNN)] | The two in-distribution empirical CDFs | Equal weights, constituents fixed in advance; no free parameters |

**Note.** $p$ denotes the five-class softmax probabilities, $l$ the corresponding logits, $h$ the 256-dimensional penultimate classification-head representation, and $z$ the 1024-dimensional standardized crop-averaged final-block [CLS] representation. All scores were oriented such that larger values indicated greater distributional shift; published scores with the opposite orientation were sign-inverted before analysis.

**Supplementary Table 35b | Fixed parameters used for post-hoc OOD detection.**

| Parameter | Value | Basis |
|---|---|---|
| kNN neighbors, k | 22 | √492, based on the discovery sample size |
| ReAct clipping percentile | 90th percentile | Prespecified from the original method |
| ViM residual dimension | 128 of 256 | Lower half of the eigenspectrum |
| GEN exponent, γ | 0.1 | Prespecified from the original method |
| Energy temperature | 1 | Fixed without tuning on OOD data |
| Covariance regularization | Ledoit-Wolf shrinkage | Closed-form covariance regularization |
| Bootstrap resamples | 2,000 | Prespecified |
| Random seed | 42 | Matched to the deployed-model analysis |

#### Supplementary Note 15 | Agentic framework configuration

The agentic framework is described in the main text (Methods), and the quality-control checks, their measurements and thresholds in Table 4 of the main text. Supplementary Table 36 gives the agent configuration, Supplementary Table 37 the findings with their numeric triggers and severity, and Supplementary Table 38 the disposition of each finding as specified to the quality-control agent. Tumor segmentation labels followed our convention (refs. 29 and 33 of the main text), adopted by Brain Tumor Segmentation in Pediatrics (BraTS-PEDs) (ref. S1).

Because the five calibrated probabilities sum to one, at most one class can exceed the threshold. A subject that was not abstained therefore falls necessarily in the moderate or high tier, and the absence of low-tier predictions carries no information about the cohort. For the same reason the number of classes above the threshold takes only the values zero and one and was not interpreted as a measure of ambiguity. The agent also reported the runner-up class, the margin between the two highest probabilities, and the proportion of the eighteen crops whose highest-probability class matched the reported one, the last as a measure of stability with respect to the sampling region.

**Supplementary Table 36 | Agent configuration.** Both agents used settings from a single version-controlled configuration file. Model documentation: ref. S31.

| Parameter | Quality-control agent | Prediction agent |
|---|---|---|
| Interface | Anthropic Messages API, function calling | Anthropic Messages API, function calling |
| Model identifier | claude-opus-5 | claude-opus-5 |
| Maximum output tokens | 8192 | 8192 |
| Reasoning effort | High | High |
| Extended reasoning | Adaptive | Adaptive |
| Sampling temperature | Not set; not accepted by the model | Not set; not accepted by the model |
| Maximum turns | 16 | 8 |
| Tools available | 8 | 3 |
| Image data received | None | None |

**Supplementary Table 37 | Findings, their numeric triggers and their severity.** Invalidating findings withhold the result, cautionary findings qualify it, and informational findings are recorded without altering the verdict.

| Finding | Trigger | Severity | Consequence for interpretation |
|---|---|---|---|
| volume_grossly_wrong | Brain mask volume < 250 mL | Invalidating | Mask volume is not anatomically plausible; the strip discarded most of the head |
| fragmented_mask | >1 connected component exceeding 5% of mask voxels | Invalidating | Brain mask is fragmented |
| lr_asymmetry_high | $\|L − R\| / (L + R) > 0.15$ | Invalidating | Brain mask is markedly asymmetric |
| tumor_outside_brain_mask | Fraction of tumor voxels outside the mask > 0.02 | Invalidating | Either the strip clipped tissue the tumor extends into, or the two are misregistered |
| seg_mask_shape_mismatch | Segmentation and mask on different sampling grids | Invalidating | Containment cannot be evaluated |
| empty_segmentation | No non-zero label voxels | Invalidating | No tumor to measure; the network ran and returned nothing |

| Finding | Trigger | Severity | Consequence for interpretation |
|---|---|---|---|
| no_brain_voxels | No voxel above 0.02 × the crop's 99.5th intensity percentile | Invalidating | No brain voxels above the background threshold |
| degenerate_intensity_range | p99.95 − p0.05 over brain voxels < $1 \times 10^{-6}$ | Invalidating | Brain voxels span no intensity range |
| normalization_range_mismatch | After clip and rescale, min or max deviates from [−1, 1] by >$1 \times 10^{-3}$ | Invalidating | The tensor handed to the network is not on the scale used in development |
| unexpected_voxel_size | Voxel size not 1.0 × 1.0 × 1.0 mm | Invalidating | The sampling region is in fixed atlas coordinates; wrong spacing samples the wrong anatomy |
| unexpected_orientation | Orientation code not LPS | Invalidating | As above, for orientation |
| unexpected_shape | Matrix not 240 × 240 × 155 | Invalidating | Preprocessing did not produce the geometry used in development |
| axes_reordered_vs_nifti | Tensor spatial axes differ from the NIfTI axis order | Invalidating | The convolutions receive reordered anatomy without raising an error |
| gpu_too_new_for_segmentation | Segmentation log reports no compiled kernel for the device | Cautionary | The GPU is newer than the segmentation image supports; every subject fails at that stage |
| segmentation_stage_failed | Segmentation container exited non-zero, or its input could not be staged | Cautionary | No tumor volume, label breakdown or containment; the prediction is unaffected |
| upstream_qc_inconclusive | Quality-control agent returned no verdict | Cautionary | Preprocessing is unverified, which is not the same as verified clean |
| non_axial_2d_acquisition | Two-dimensional T2 acquisition in a sagittal or coronal plane | Cautionary | Registration cannot be verified from its transform parameters alone |
| multifocal_segmentation | >1 component exceeding 10% of tumor voxels | Informational | May be genuinely multifocal disease |
| tumor_edge_outside_mask_minor | Fraction outside the mask >0.005 and ≤0.02 | Informational | Partial-volume effect at the mask boundary |
| label_breakdown_not_interpretable | Channels duplicated from a single bulk sequence | Informational | Per-label split withheld; whole-tumor volume remains valid |

**Supplementary Table 38 | Disposition of each finding, as specified to the quality-control agent.** The final group lists findings that the agent was instructed not to treat as failures.

| Finding | Disposition | Reasoning |
|---|---|---|
| Brain mask volume below 250 mL | ESCALATE | A mask that small cannot be a pediatric brain |
| Fragmented mask, or high left-right asymmetry | ESCALATE | Structural failure of the extraction |
| Clipped cerebellum or brainstem | ESCALATE | High consequence in the posterior fossa |
| Empty segmentation | ESCALATE | Nothing remains to measure |
| Edema with essentially no tumor | ESCALATE | Edema is peritumoral by definition, so the tumor was missed |
| Meaningful fraction of tumor outside the brain mask | ESCALATE | Either the strip clipped tissue or the two are misregistered |
| A check that did not run, covering a high-consequence signal | ESCALATE | Containment, clipping or fragmentation can no longer be excluded |
| Two-dimensional non-axial acquisition | FLAG | Registration cannot be verified from its transforms; clean values do not clear it |
| More than one large segmentation component | FLAG | May be genuinely multifocal |
| A check that did not run, covering a low-consequence signal | FLAG | Absent evidence, graded by what it covers |
| Multiple candidate brain masks on disk | FLAG | The measured volume is provisional |

| Finding | Disposition | Reasoning |
|---|---|---|
| Brain mask volume above 1500 mL, structurally intact | No finding | Head size or a generous boundary; there is no upper bound on volume |
| Line-search warning with transforms within bounds | No finding | Benign optimizer message |
| Absent enhancing component | No finding | Several pediatric posterior fossa histologies are characteristically non-enhancing |
| Absent edema or cystic component | No finding | Their presence is tumor-dependent |
| Stray single-voxel labels | No finding | Noise rather than structure |
| A few tumor voxels outside the mask | No finding | Partial-volume effect at the boundary |
| Label breakdown uninterpretable under channel duplication | No finding | A statement of scope; whole-tumor volume remains valid |

#### Supplementary Note 16 | Agent system prompts

Agent system prompts, reproduced verbatim as deployed. The statuses named in the quality-control prompt as absent evidence are the values a tool returns when it could not perform its check.

##### Quality-control agent

You are a QC reasoning agent for a pediatric brain-MRI pipeline
(preproc -> skull-strip -> tumor-segmentation -> normalization). You see only JSON
from tools, never images. Run the tools in order — inspect_inputs, acquisition_qc,
run_pipeline, reg_qc, strip_qc, seg_qc, containment_qc, norm_qc — calling each one
(never just describing it).

You are not a lookup table. Weigh ALL the evidence together and reason about it:

- A single soft flag does not automatically mean escalate. Integrate every signal.

- Brain-mask volume on its own is weak evidence. A volume above the soft band on a
structurally clean mask — one connected component, low left-right asymmetry, sane
registration — is a big head or a mildly generous boundary, not a failed strip.
Do not FLAG on volume alone. strip_qc reports it as volume_note precisely because
it is informational, and there is no upper bou.nd on volume at all: a large mask is
a large head until something structural says otherwise. What indicates a bad strip
is structure: fragmentation, high asymmetry, or a volume below the 250 mL floor,
which is the one volume reading that fails on its own because a mask that small
cannot be a brain.

- Weigh signals by clinical consequence, not by count:
* HIGH stakes (escalate hard): a clipped cerebellum/brainstem or fragmented mask —
subtle but dangerous in posterior fossa, can cause downstream misdiagnosis.
* LOW stakes (do not flag): a mildly over-inclusive mask — slightly high volume,
symmetric, single component — is cosmetic and carries no downstream risk.

- A line-search warning with sane transforms is benign; do not treat it as a failure.

- Acquisition: a 2D T2 must be axial. A 2D sagittal or coronal T2 does not by itself
mean the case failed, but registration cannot be trusted on its transforms alone —
FLAG it so a human checks registration, even when reg_qc looks clean. A 3D
acquisition is acceptable in any orientation. Note that clean reg_qc numbers do NOT
clear this flag: the transform can be sane and the alignment still wrong.

- Modality availability changes what the segmentation MEANS. inspect_inputs reports
seg_mode. When only one bulk sequence (T2w or FLAIR) is real, the other channels are
duplicates of it — the network never saw the contrast differences the label
definitions rest on, so the enhancing/nonenhancing/cystic split is an artifact of
the bypass, not a finding. In that mode seg_qc reports whole_tumor_ml only and
raises label_breakdown_not_interpretable; treat that flag as a statement of scope,
NOT as a QC failure — the whole-tumor segmentation is still valid and is what the
downstream posterior-fossa histology model needs. Do not escalate on it, and do not
reason about which label dominates in that mode.
Missing T2w entirely IS a hard problem: the prediction step reads T2w specifically.

- Segmentation labels are BraTS-PEDs: 1 = enhancing tumor, 2 = nonenhancing tumor,
3 = cystic component (all three are tumor proper), 4 = peritumoral edema, which
surrounds the tumor rather than being part of it.
An empty segmentation is a hard ESCALATE — there is nothing to measure downstream.
So is edema with essentially no tumor: edema is peritumoral by definition, so
finding it with nothing to surround means the tumor itself was missed.
Do NOT treat an absent enhancing component as a failure. Many pediatric tumors,
low-grade glioma especially, are characteristically non-enhancing; a segmentation
that is entirely label 2 with no label 1 is an expected presentation, not an error.
The same goes for absent edema or absent cyst — their presence is tumor-dependent.
Stray single-voxel labels are noise rather than structure; weigh them as such.
Multiple large components may be genuinely multifocal disease, so treat that as a
FLAG for a human glance rather than an automatic failure.

- Containment: tumor voxels are expected to lie inside the brain mask. A meaningful
fraction outside is HIGH stakes — ESCALATE — because it usually means either the
strip clipped tissue the tumor extends into, or the segmentation and brain mask are

misregistered relative to each other. A handful of edge voxels outside the mask is partial-volume noise at the boundary; weigh that as minor.

- A check that did not run is not a check that passed. When a tool returns status tool_error, no_log, no_mask, no_seg_output, no_norm_output, no_t2 or missing_inputs, that signal is absent rather than clean, and no other tool supplies it. Do not return PROCEED while any check is in that state. Identify the check that did not run and grade on consequence: FLAG where the missing signal is low-consequence, ESCALATE where it covers a high-consequence signal — containment, a clipped cerebellum or brainstem, mask fragmentation — that you are consequently unable to exclude. This is distinct from a check that ran and returned an adverse result: empty_seg and shape_mismatch are findings, and retain the handling described above.

- The segmentation stage crashing is not the same as the segmentation being empty, and the two must not be graded alike. An empty mask is a FINDING: the network ran, looked, and saw nothing, which is a hard ESCALATE per the rule above. A stage that exits non-zero produced no mask at all, so seg_qc and containment_qc return no_seg_output and the run_pipeline result carries segmentation_stage_failed. That is absent evidence ABOUT THE TUMOR, and it is evidence about nothing else. The classifier reads the normalized T2, which does not pass through the segmentation stage, so what bears on the prediction is registration, skull strip and normalization geometry — all of which still ran and are still yours to weigh. Do not ESCALATE on the absent containment check alone in this case: containment asks whether the tumor mask lies inside the brain mask, and here neither the tumor mask nor the prediction depends on the answer. FLAG it, say that no tumor measurement exists for this subject, and grade the checks that did run on their own merits. If those are themselves adverse, escalate on them.

- If strip_qc reports multiple_mask_candidates, the measured volume is provisional: more than one brain mask is on disk and the tools may not be describing the same object. Say so in the rationale rather than treating the number as settled.

Decide one of:
PROCEED — all signals consistent with a good result.
FLAG — usable, but one borderline signal a human should glance at.
ESCALATE — a genuine failure, or a high-consequence signal you cannot rule out.

QC CONFIDENCE qualifies the DECISION, not the quality of the study. It is your own assessment of the evidence, and is a different quantity from the prediction agent's PREDICTION CONFIDENCE, which is computed from the model's probabilities and is not a judgement at all. The two appear side by side in the case report, so they are named apart and must not be reasoned about as though they were the same scale. An unambiguous failure warrants ESCALATE at high confidence; do not reduce the rating because the case itself is poor.
high — every check ran and the signals are concordant. The decision would not change if any single measurement shifted modestly.
moderate — every check ran, but the decision rests on one borderline value, or on signals that are partially discordant.
low — a check did not run, a measurement is provisional (for example multiple_mask_candidates), or the signals are genuinely in conflict.
A decision reached with any check unrun is low confidence, not moderate.

Output, after norm_qc:
1. DECISION: PROCEED / FLAG / ESCALATE
2. QC CONFIDENCE: high / moderate / low
3. RATIONALE: one or two sentences citing the actual numbers and how you weighed them.
4. RESOLUTION CRITERIA: the specific evidence that would raise the confidence or change the decision — the check to re-run, the measurement to confirm, or the image a reader should inspect. State "none; the evidence is complete" only when every check ran and the signals are concordant.

#### Prediction agent

You are an agent for prospective pediatric posterior fossa histology prediction. Call the tools in order: inspect_image, qc_context, predict. Call each one; never just describe it.

The prediction itself is deterministic and is NOT yours to revise. Never override the gate, re-rank the classes, or substitute your own reading of the probabilities for the reported one. Your job is to say what the result can and cannot support.

What the numbers mean:

- The gate is `any calibrated q_ >= 0.5`. If nothing clears it the system abstains and there is no prediction to defend. Report the abstention plainly, name which class came closest, and stop; do not speculate about which histology it "would" have been.

- PREDICTION CONFIDENCE is computed, not judged. predict() returns both the value and the tier — report them, never a figure of your own. It is unrelated to the QC agent's QC CONFIDENCE, which is that agent's certainty in its own verdict; both appear in the case report, so do not carry a reading across from one to the other.

- DISTRIBUTIONAL CERTAINTY is a second, separate quantity: 1 minus the normalized entropy of the calibrated distribution, where 1 is all mass on one class and 0 is uniform across five. It answers a different question from PREDICTION CONFIDENCE — how concentrated the whole distribution is, rather than how much mass the winner holds. They can disagree, and when they do, say so rather than reporting only one.

- Two predictors can serve a request, and predict() reports which in `model_kind`. Read it before interpreting the numbers, because the two differ in what the gate means:

* `3dino` (default) — one multiclass softmax over the five classes with a single global temperature. The probabilities sum to 1, so AT MOST ONE class can clear the 0.50 gate; n_above_gate is 0 or 1 and carries no information beyond whether the subject was abstained. PREDICTION CONFIDENCE is the calibrated probability of the predicted class. Because the gate and the tiers are the same quantity, a subject that was not abstained is always Moderate or High — a Low tier cannot occur, and its absence says nothing. A single global temperature rescales every class by the same exponent and therefore cannot change which class wins.

* `resnet` — five independent one-vs-rest heads with per-class temperatures, gated on calibrated q_. Several classes clearing the gate at once is the normal case rather than a conflict; say how many cleared it and name the runner-up. The two confidence quantities mean the same things they do under 3dino — PREDICTION CONFIDENCE is the probability of the predicted class, DISTRIBUTIONAL CERTAINTY is 1 minus the normalized entropy — but here they are read off the coupled softmax while the gate is applied to calibrated q_. Those are different distributions, so unlike 3dino a subject can clear the gate with a PREDICTION CONFIDENCE below 0.50: a Low tier is reachable, and it means the coupled distribution is flat even though one head was confident.
Ependymoma carries a temperature of 20.3 that compresses its calibrated q_ into roughly [0.34, 0.57], so it clears the gate in most subjects regardless of the image: when it is the only class above the gate that is weak evidence, and when it is the argmax its calibration is near-degenerate and the reading is less trustworthy than for the other four. None of this applies under `3dino`.

- Under `3dino`, predict() also reports view_disagreement and view_argmax_agreement: the spread across the 18 jittered crops the backbone reads, and how many of them voted for the reported class. High disagreement means the crops did not see the same thing, which points at the input rather than at the histology. Report it when it is high; do not convert it into a confidence of your own.

- A narrow top-2 margin with a confident-looking top class is a near-tie. Say which two classes are competing rather than reporting only the winner.

Upstream context changes what the prediction is worth:

- If qc_context reports channels duplicated from a single sequence, preprocessing differed from the multiparametric path the model was developed on. Note it as a caveat on transferability. Do not treat it as invalidating the prediction.
- If upstream QC was not clean, or the image shape or intensity range is unexpected, say that the input may not match training conditions.
- The flag upstream_qc_inconclusive means the QC agent ran but never reached a verdict, so preprocessing is unverified rather than verified-clean. Report it as absent evidence, on the same footing as a check that did not run: it is not a mild version of PROCEED.
- If there is no upstream context at all, say that you cannot confirm how the subject was prepared, rather than assuming it was done correctly.

- If predict returns any status other than ok — no_input, models_missing, or a tool error — there is no result to interpret. Report that the prediction could not be produced, and why. Do not supply a class, and do not describe it as an abstention: abstention is a specific finding the gate reaches from real probabilities, not a synonym for a computation that did not complete.

State plainly that this model was validated on treatment-naive, posterior-fossa-confined tumors whose histology is one of the five classes, and that nothing in the output can tell you whether this subject meets those conditions.

Output:
1. PREDICTION: the class, or ABSTAIN
2. PREDICTION CONFIDENCE: tier and value, exactly as predict() reported them
3. RATIONALE: two or three sentences citing the actual numbers
4. LIMITATIONS: what this result cannot establish — the conditions it assumes, the caveats carried from upstream QC, and the differentials it does not exclude.

#### Supplementary Note 17 | Reporting checklist (TRIPOD+AI)

The study is reported according to TRIPOD+AI (ref. S32). Supplementary Table 39 gives the location of each item in the main text (MT) or the Supplementary Information (SI); highlighted entries are not yet reported and need text before submission.

Supplementary Table 39 | TRIPOD+AI checklist. Item numbers follow ref. S32.

| Item | Checklist topic | Where reported |
|---|---|---|
| 1 | Title | PedDx: An End-to-End Deep Learning Framework for Pediatric Posterior Fossa Tumor Diagnosis with Agentic AI–Enabled Independent External Evaluation |
| 2 | Abstract | Abstract to be written |
| 3a | Healthcare context and rationale; existing models | MT Introduction, paragraphs 1–2 (refs. 1–14) |
| 3b | Target population, intended purpose and users | MT Introduction, paragraphs 3 and 5 (treatment-naïve posterior fossa tumors; assistive use with deferral); |
| 3c | Known health inequalities | Not reported |
| 4 | Objectives | MT Introduction, final paragraph |
| 5a | Data sources and representativeness | MT Methods, Study design and cohorts; Table 1 |
| 5b | Dates of data collection | Not reported |
| 6a | Setting, number and location of centers | MT Methods, Study design and cohorts; Table 1 (contributing institutions) |
| 6b | Eligibility criteria | MT Methods, Study design and cohorts; SI Note 1 (27 CHOA non-medulloblastoma cases) |
| 6c | Treatments and how they were handled | MT Methods, Study design and cohorts (treatment-naïve in-distribution data; post-treatment cases analyzed as clinical OOD) |
| 7 | Data preparation and quality checking | MT Methods, Image preprocessing and model architecture; Quality-control measurements and states; SI Notes 10 and 15 |
| 8a | Outcome definition and assessment | MT Methods, Study design and cohorts (five histologic classes) |
| 8b | Qualifications of outcome assessors | MT Methods, Study design and cohorts (histopathologic confirmation through CBTN records or patient charts) |
| 8c | Blinding of outcome assessment | Not reported |
| 9a | Choice of predictors | MT Introduction, paragraph 5; Methods, Image preprocessing and model architecture (axial T2-weighted MRI only) |
| 9b | Definition and measurement of predictors | MT Methods, Image preprocessing and model architecture; SI Notes 10–11 |
| 9c | Subjective predictor assessment | Not applicable: features are computed automatically |
| 10 | Sample size | MT Methods: Study design and cohorts; SI Note 1 |
| 11 | Missing data | MT Methods, Study design and cohorts (170 exclusions); Quality-control measurements and states (missing sequences) |

| Item | Checklist topic | Where reported |
|---|---|---|
| 12a | Use and partitioning of the data | MT Methods, Study design and cohorts (approximately 80/20 split stratified by diagnosis); Model selection and internal validation (nested cross-validation) |
| 12b | Handling of predictors | MT Methods, Image preprocessing and model architecture (crop normalization, feature standardization) |
| 12c | Model type, building steps, tuning, internal validation | MT Methods, Model selection and internal validation; SI Notes 10–13 |
| 12d | Heterogeneity across clusters | MT Results, Independent evaluation of the complete workflow (performance by cohort and site, Fig. 6d) |
| 12e | Performance measures and model comparison | MT Methods, Statistical analysis; Calibration, uncertainty, and selective prediction; SI Note 3 |
| 12f | Model updating | MT Methods, Calibration, uncertainty, and selective prediction (weights, temperature and threshold fixed; no refitting on external data) |
| 12g | How predictions were calculated | MT Methods, Image preprocessing and model architecture; Calibration, uncertainty, and selective prediction; MT Code availability |
| 13 | Class imbalance | MT Methods, Study design and cohorts (stratified split); SI Supplementary Table 32 (class weighting searched as a hyperparameter) |
| 14 | Fairness | Not reported |
| 15 | Model output and thresholds | MT Methods, Calibration, uncertainty, and selective prediction (calibrated probabilities, most probable class, abstention below 0.50, confidence tiers) |
| 16 | Differences between development and evaluation data | MT Results, External classifier evaluation and class-specific limitations; Independent evaluation of the complete workflow; Methods, Independent external evaluation |
| 17 | Ethical approval and consent | MT Methods, Study design and cohorts (IRB approval for CBTN/CHOP and CHOA; data transfer agreement); IRB approval for WCH data |
| 18a | Funding and role of funders | MT Acknowledgements; |
| 18b | Conflicts of interest | MT Competing interests |
| 18c | Protocol | Not Applicable |
| 18d | Registration | The study was not registered |
| 18e | Data sharing | MT Data availability |
| 18f | Code sharing | MT Code availability |
| 19 | Patient and public involvement | No involvement |
| 20a | Flow of participants | SI Supplementary Fig. 1; MT Results, Study overview |
| 20b | Participant characteristics by data source | MT Table 1; SI Supplementary Tables 1a,b; |
| 20c | Evaluation versus development data | MT Table 1 (diagnoses by cohort); SI Supplementary Table 1b (acquisition characteristics) |
| 21 | Participants and events in each analysis | MT Tables 1–3; Results (n per cohort and class); SI Notes 2–9 |
| 22 | Full model specification and access | MT Methods, Image preprocessing and model architecture; Agentic workflow and application (Fig. 8); MT Code availability |
| 23a | Performance with confidence intervals, key subgroups | MT Results; Tables 2–3; SI Notes 3–9 (by histology; no sociodemographic subgroups) |
| 23b | Heterogeneity across clusters | MT Results (replication, CHOA and Macquarie; Fig. 6d) |
| 24 | Model updating results | Not applicable: the model was not updated |
| 25 | Interpretation | MT Discussion |
| 26 | Limitations | MT Discussion, limitations paragraph |
| 27a | Handling of poor-quality or unavailable input | MT Methods, Quality-control measurements and states; Processing verdict and reporting; Results (abstention) |
| 27b | User interaction and required expertise | MT Methods, Agentic workflow and application; Processing verdict and reporting (Fig. 8); |
| 27c | Next steps | MT Discussion, final paragraph |
